# Effects of collaborative clinical visit agenda-setting interventions: A systematic review and meta-analysis

**DOI:** 10.64898/2026.08.30.26361729

**Authors:** Ailyn Sierpe, Renata W. Yen, Annika Milliman, Elizabeth Cady, Boyoung Ahn, Anne E. Dade, Anna Marie Devito, Bradley A. Eckert, Vismaya V. Gopalan, Stephanie C. Krasinski, Meredith A. MacMartin, Sophia G. Musacchio, Jingyi Zhang, Catherine H. Saunders

**Affiliations:** Dartmouth Health, 1 Medical Center Dr, Lebanon, NH 03756, United States; IQ Health Science Department, Radboud University Medical Center, Geert Grooteplein Zuid 10, 6525 GA Nijmegen, Netherlands; The Dartmouth Institute for Health Policy and Clinical Practice, Geisel School of Medicine at Dartmouth College, 1 Medical Center Dr, Lebanon, NH 03756, United States; Center for Technology and Behavioral Health, Geisel School of Medicine at Dartmouth College, 46 Centerra Pkwy, Lebanon, NH 03766, United States; The Johns Hopkins University School of Medicine, 733 N Broadway, Baltimore, MD 21205, United States; Hartford HealthCare Cancer Institute, Hartford HealthCare, 195 Retreat Ave, Hartford, CT 06103, United States; University of Pennsylvania Perelman School of Medicine, 3400 Civic Center Blvd, Philadelphia, PA 19104, United States

**Keywords:** Clinical agenda-setting, pre-visit planning, patient-clinician communication, visit communication, patient participation, patient-centered care, systematic review, meta-analysis

## Abstract

**Background:** Agenda-setting is a fundamental patient-centered communication practice in which a clinician works with a patient to elicit, propose, and organize topics for discussion during a clinical encounter. Various agenda-setting interventions have been developed, including patient-facing tools and clinician training, but their effects have not been systematically evaluated. We aimed to determine the effects of these interventions on encounter, patient, care partner, and clinician outcomes.

**Methods:** We searched grey literature and seven databases, including PubMed, from inception through July 2025 for randomized and non-randomized comparative studies of interventions designed to promote or improve clinical visit agenda-setting. Two reviewers independently screened articles and extracted data, with a third reviewer resolving conflicts. We assessed risk of bias using RoB 2 for randomized studies and ROBINS-I for non-randomized studies. We conducted random effects meta-analyses when outcomes were sufficiently comparable, assessed heterogeneity using I^2^, and rated certainty of evidence using GRADE. Post hoc exploratory subgroup analyses examined study design, adjustment status, and intervention structure.

**Results:** Twenty-nine articles describing 22 unique studies met the inclusion criteria, including 13 randomized and nine non-randomized studies. Agenda-setting interventions increased the occurrence of agenda-setting (risk ratio 5.43, 95% confidence interval (CI) 2.06 to 14.28, I^2^=34.6%) and favored the intervention for concerns addressed when measured as a continuous outcome (standardized mean difference (SMD) 0.37, 95% CI 0.16 to 0.57, I^2^=65.3%) and overall clinician satisfaction (SMD 0.50, 95% CI 0.23 to 0.78, I^2^=0.0%). There were no clear differences in the number of concerns raised (mean difference (MD) 0.21, 95% CI −0.19 to 0.61, I^2^=59.6%), visit duration (MD 0.64 minutes, 95% CI −0.83 to 2.12, I^2^=51.4%), or overall patient satisfaction (SMD 0.05, 95% CI −0.05 to 0.15, I^2^=47.0%). Potentially important heterogeneity was present for four of these six outcomes. Post hoc exploratory subgroup analyses did not provide clear evidence that effects varied by study design, adjustment status, or intervention structure. Risk of bias was often high, serious, or critical, and certainty of evidence was low or very low for all pooled outcomes.

**Conclusions:** To our knowledge, this is the first comprehensive synthesis of clinical visit agenda-setting interventions. Such interventions may increase the occurrence of agenda-setting and the extent to which patient concerns are addressed without increasing visit length. However, the certainty of evidence was low or very low, and the available evidence does not establish a superior intervention structure.

## Introduction

Collaborative clinical visit agenda-setting is a foundational communication practice in which clinicians and patients work together before or at the start of a visit to identify topics for discussion.^1–4^ More technically, agenda-setting is a three-step process: visit agenda items are elicited, proposed, and then organized.^1,5,6^ Agenda-setting has been associated with greater patient satisfaction and improved clinician understanding of patient concerns, while patient-centered communication may improve treatment adherence and health outcomes.^1,5,7,8^ Critically, agenda-setting may reduce the number of concerns left unaddressed by the end of the visit, including concerns raised late in the encounter.^1,9^ Despite these benefits, some clinicians may hesitate to use agenda-setting due to concerns that it would add time to an already constrained visit. However, trials suggest that agenda-setting can improve communication without significantly lengthening visits.^4,10^

Agenda-setting is often taught in medical education and is widely acknowledged as a best practice.^3,5,11,12^ However, many clinicians have only a limited understanding of or do not use agenda-setting fully, with observational studies finding considerable variation in the occurrence and skill level of agenda-setting.^1,5,6,11–13^ Assessing agenda-setting presents a related challenge, as studies have defined and operationalized it differently, with no single universally adopted measure.^6,14,15^

Researchers have developed a broad range of interventions to promote or strengthen agenda-setting. Patient-facing interventions have used checklists or pre-visit questionnaires to help patients and their care partners identify and prioritize topics before the visit.^4,16–23^ Clinician-facing interventions have generally involved training in agenda-setting skills, including topic elicitation workshops.^5,10,24–28^ Some studies have combined both patient-facing and clinician-facing approaches.^29^ The outcomes used to evaluate these interventions have varied just as widely, which has made determining overall efficacy challenging. Outcomes have included measures such as visit length, the number of topics discussed, or ‘surprise’ topics raised only at the end of the visit.^1,4,29,30^ Others have focused on patient-reported measures of clinical communication or general satisfaction.^5,29^

Despite its importance as a patient-centered communication practice, collaborative agenda-setting has remained understudied. We do not yet know how effective agenda-setting interventions are overall, whether some approaches work better than others, and which patient populations are most likely to benefit. To address this gap, we conducted what is, to our knowledge, the first systematic review and meta-analysis of clinical visit agenda-setting interventions.^31^ Our primary aim was to assess the effects of agenda-setting interventions. We also characterized the interventions and their delivery, and examined how agenda-setting was operationalized and measured.

## Methods

### Study design

We conducted a systematic review and meta-analysis of collaborative agenda-setting interventions. Our protocol was published elsewhere^31^ and registered in the International Prospective Register of Systematic Reviews (PROSPERO, CRD42023468045).^32^ We conducted the review with guidance from the *Cochrane Handbook for Systematic Reviews of Interventions*^33^ and report it in accordance with the Preferred Reporting Items for Systematic Reviews and Meta-Analyses (PRISMA) checklist (see **Appendix 1**).^34^

The review addressed three prespecified research questions:

1. What are the effects of clinical visit agenda-setting interventions on outcomes relating to the clinical encounter itself, patients, care partners, and clinicians, as well as any other study-specified outcomes?
2. What are the characteristics and delivery attributes of clinical visit agenda-setting interventions?
3. How has clinical visit agenda-setting been operationalized and measured?

### Inclusion criteria

We included randomized and non-randomized comparative studies evaluating interventions designed to promote or improve clinical visit agenda-setting. No intervention, usual care, or another agenda-setting intervention were eligible comparators. We included non-randomized studies if they used a pre-post or other quasi-experimental comparative design. We included pilot and feasibility studies and excluded studies without a comparison and those reporting only qualitative findings. Mixed methods studies were included when they reported an otherwise eligible quantitative comparison.

We did not exclude studies based on clinical setting or health condition. Participants could include patients, care partners, clinicians, and anyone else involved in an agenda-setting intervention. Included interventions could be patient-facing, clinician-facing, or both. We defined agenda-setting as a collaborative process in which a clinician elicited, and when applicable, proposed or organized topics for discussion together with the patient.^1,5,6^ The intervention had to be designed to initiate this process before or at the start of the clinical encounter.^1,2,8,9,13,31,35,36^ While eliciting patient concerns to exhaustion is best practice, it is not generally operationalized as a requirement for agenda-setting to have occurred, so we did not include this in our definition.^2,6,14,15,31,37^ We excluded interventions in which agenda-setting was only incidental to the intervention’s primary purpose, such as interventions primarily focused on goal-setting or addressing patient-clinician communication more broadly.

We included all study-specified outcomes, such as visit process measures, patient- or care partner-reported outcomes, clinician-reported outcomes, and observer-assessed outcomes. We did not restrict eligibility by publication language or status. Further details on our inclusion and exclusion criteria are reported in our protocol.^31^

### Search strategy

With assistance from two biomedical research librarians, we developed and piloted a search strategy accounting for the varied terminology used to describe agenda-setting. Search terms included agenda-setting, agenda-mapping, topic elicitation or solicitation, patient agendas and priorities, opening statements, and visit structure. As there is no Medical Subject Headings (MeSH) term for agenda-setting, we adapted keywords to each database. Our complete list of searches for every data source, including exact queries used, is provided in **Appendix 2**.

We searched APA PsycInfo, the Cochrane Library, Cumulative Index to Nursing and Allied Health Literature (CINAHL), MEDLINE via PubMed, ProQuest, Scopus, and Web of Science. Each source was searched from inception through July 2025, with no date, language, or other search restrictions. We supplemented the database searches by conducting backward and forward citation searches, searching ClinicalTrials.gov, screening the first 25 pages of Google Scholar results in default relevance order, and reviewing references suggested by colleagues with expertise in agenda-setting.

### Data screening

We imported our database search results into Rayyan and removed duplicate items.^38^ Before beginning formal screening, the reviewers piloted the inclusion and exclusion criteria on a sample of 50 abstracts and resolved differences in their interpretation of the criteria. Each title and abstract was then screened independently by two reviewers, with a third independent reviewer resolving disagreements. Articles retained after title and abstract screening were advanced to full-text review by two independent reviewers, with a third reviewer resolving disagreements. We did not use automated tools to make screening decisions.

When multiple articles described the same intervention and participant sample, we linked the articles and treated them as one study. We designated the article containing the most complete comparative intervention data as the primary article, or when articles were similarly complete, chose the most recent article. Companion articles were used to supplement information about intervention development, delivery, implementation, and outcomes. We did not count companion articles as separate studies or include the same participants more than once in our analysis.

### Data extraction

We developed and piloted a standardized data extraction form with items adapted from the Template for Intervention Description and Replication (TIDieR) checklist.^39^ One reviewer extracted the data from the included studies, while a second reviewer independently verified all extracted data, and a third reviewer resolved any conflicts. We did not use automated tools for extraction.

For each study, we extracted study aims and design, dates, location, setting, participant eligibility criteria, recruitment strategy, and sampling procedure. Participant characteristics were extracted separately for patients, care partners, and clinicians. Patient and care partner characteristics included age, sex or gender, race, ethnicity, health literacy, education, and socioeconomic status. We also extracted patients’ health conditions. Clinician characteristics included age, gender, race, ethnicity, clinical role, and years in practice. For each group, we recorded baseline enrollment by study arm, attrition, and reasons for loss to follow-up. For each intervention or comparator, we extracted its name and rationale, materials, procedures, who delivered it, mode and location of delivery, the intervention period, duration, frequency or repetition, length of follow-up, tailoring, modifications, and planned and observed fidelity.

We extracted all study-specified outcomes, extracting each outcome’s definition, scale limits, method of measurement, and measurement period. We sought all measures, time points, and analyses reported for each outcome. We extracted results separately for the intervention and comparison groups. For dichotomous outcomes, we extracted numerators, denominators, and any reported risk or odds ratios, confidence intervals, and p values. For continuous outcomes, we extracted the analytic sample size, mean, standard deviation, standard error, interquartile range, confidence interval, and p value. For each outcome, we recorded whether the estimate was adjusted and, when reported, the variables included in the adjustment. We also captured qualitative descriptions of results when reported.

When information was missing or unclear, we attempted to contact study authors. If the requested information remained unavailable, we derived missing values from other reported statistics where appropriate, following guidance in the *Cochrane Handbook for Systematic Reviews of Interventions*.^33^

### Risk of bias assessment

To assess risk of bias, we used the revised Cochrane risk-of-bias (RoB 2) tool^40^ for randomized trials and the RoB 2 extension for cluster-randomized trials when clinicians or practices were allocated.^41^ We used the Risk of Bias in Non-Randomized Studies of Interventions (ROBINS-I) tool^42^ for non-randomized studies. Companion articles describing the same intervention and participants were considered together in assessing the relevant study. One reviewer assessed each study, with a second reviewer confirming the assessment and a third reviewer resolving any disagreements. Randomized trial results were assessed as having a low risk of bias, some concerns, or high risk of bias. Non-randomized studies were assessed as having low, moderate, serious, or critical risk of bias. We considered the domain level and overall judgment of these assessments. Risk of bias assessments did not impact study inclusion.

### Data synthesis

Before synthesis, we grouped outcomes according to their underlying construct and measurement format. Within each group, we assessed whether measures were sufficiently comparable for quantitative synthesis. When an article reported more than one measure of the same construct, we selected the measure that most closely represented the relevant construct and that was most comparable with the measures reported by the other contributing studies. This prevented the same participants from contributing overlapping data.

We conducted a meta-analysis when at least two studies reported sufficiently comparable data for the same outcome. For dichotomous outcomes, we calculated risk ratios (RRs). For continuous outcomes, we calculated mean differences (MDs) when studies used the same measurement scale and standardized mean differences (SMDs) when they used different scales to assess the same construct. We used random effects models as we expected intervention effects to vary across clinical settings and intervention approaches. We reported pooled estimates with 95% confidence intervals (CIs) and set statistical significance at p<0.05. Analyses were conducted using the meta^43^ and metafor packages^44^ in R (v4.6.1) via RStudio (v2026.08.1+195).^45^

For each meta-analysis, we assessed heterogeneity using Cochran’s Q (χ^2^) test and the I^2^ statistic. In cases where there was sufficient heterogeneity (p<0.10 and I^2^>40%),^46,47^ we examined differences in study design, participants, setting, and intervention characteristics as possible explanations and considered their influence on the pooled estimate. For outcomes that could not be pooled, we assessed the individual study findings for variation in the direction and magnitude of effects and for notable outliers.

In a change from the protocol,^31^ for meta-analyses containing at least three studies, we generated funnel plots of effect estimates against their standard errors and visually assessed them for asymmetry and possible small-study effects. We considered graphical assessments containing only two studies uninformative. As visual interpretation is limited when few studies are available, we treated these assessments as descriptive and did not assume that asymmetry represented publication bias.^33^ We retained the protocol-specified threshold of at least 10 studies for Egger’s regression, with p<0.05 indicating statistically significant asymmetry. For meta-analyses containing fewer than three studies and outcomes not included in a meta-analysis, we assessed publication bias qualitatively.

All included studies also contributed to a narrative synthesis. We developed evidence tables summarizing study design, setting, participant characteristics, enrollment, intervention and comparison characteristics, and reported outcomes. Following the Synthesis Without Meta-analysis reporting guideline, we grouped findings by outcome and summarized the direction and magnitude of intervention effects.^48^ We also narratively synthesized intervention characteristics and delivery attributes, along with how agenda-setting was operationalized and measured.

Where data allowed, we conducted post hoc exploratory subgroup analyses. We examined differences by study design (randomized versus non-randomized studies), adjustment status (adjusted versus unadjusted estimates), and intervention structure (structured versus unstructured agenda-setting interventions).

### Certainty of evidence assessment

We used the Grading of Recommendations Assessment, Development and Evaluation (GRADE) approach^49^ to assess certainty of evidence for each pooled outcome. We considered risk of bias, inconsistency, indirectness, imprecision, and publication bias, as well as criteria that can increase certainty in non-randomized evidence. We classified certainty as high, moderate, low, or very low.

## Results

### Study selection

Our searches identified 6,663 articles, with 5,164 from databases and 1,499 from other sources (**Figure 1**). After removing 5,103 duplicates, we screened 1,560 articles at the title and abstract level and excluded 1,518. We retrieved and screened 42 full-text articles. Thirteen of these articles were excluded. We included the remaining 29 articles in the review, representing 22 unique studies of clinical visit agenda-setting interventions. Articles excluded in full-text review are provided in **Appendix 3**.

**Figure 1.**
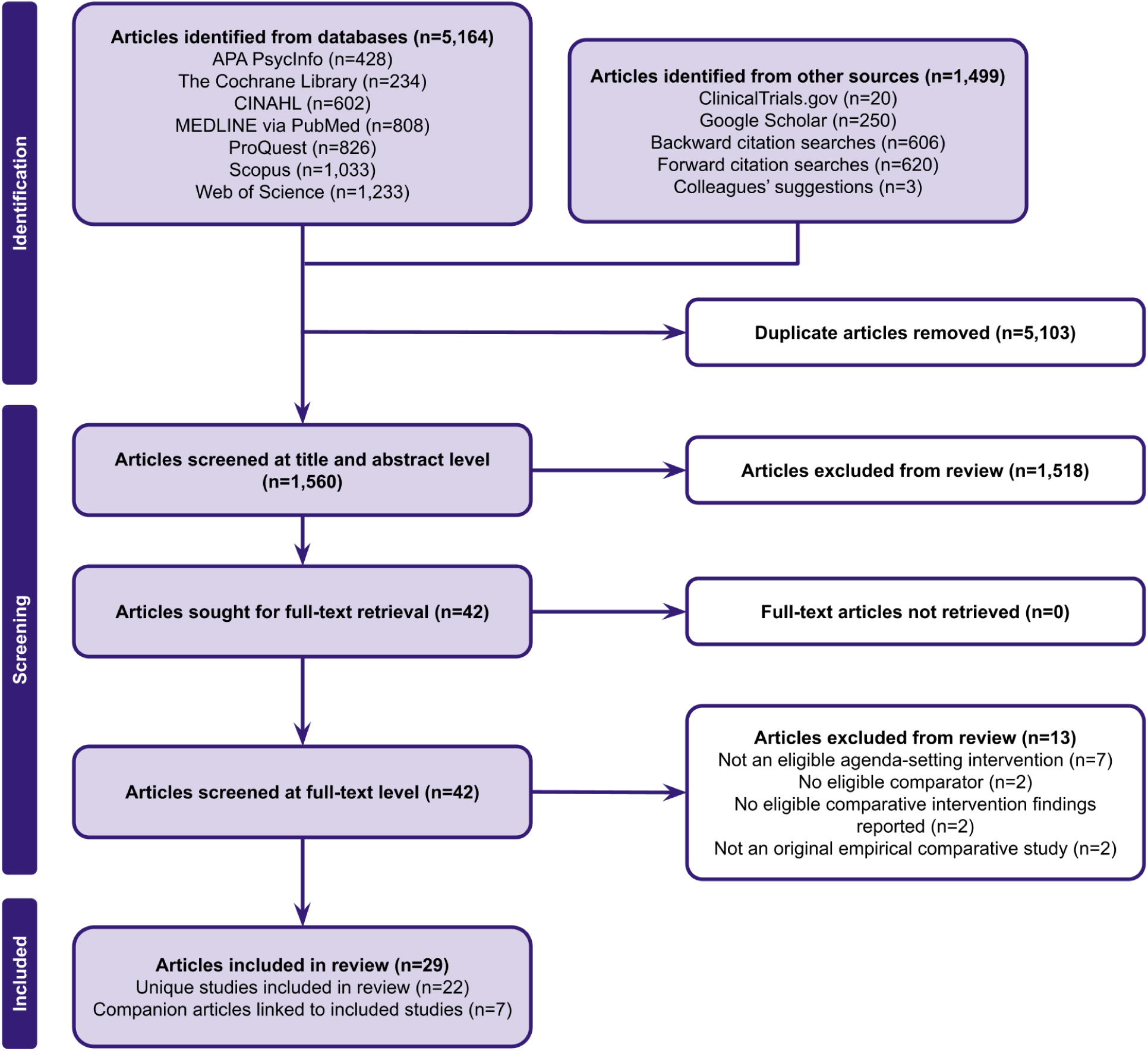
PRISMA flow diagram^34^

Seven articles were treated as companion articles rather than separate studies because they described the same intervention and participant sample as a primary article. Everden et al. (2014)^30^ was linked to Early et al. (2015),^16^ Gregory and Robling (2010)^50^ and Gregory et al. (2011)^51^ were linked to Robling et al. (2012),^22^ Middleton (1998)^52^ and Middleton and McKinley (2000)^53^ were linked to Middleton et al. (2006),^29^ O’Malley et al. (2012)^54^ was linked to O’Malley et al. (2022),^21^ and Stuart et al. (2019)^55^ was linked to Leydon et al. (2018).^27^

### Study and participant characteristics

The primary articles (**Table 1**) were published between 2001 and 2024,^4,5,10,16–29,56–60^ while the companion articles dated back to 1998.^30,50–55^ Thirteen studies (59.1%) were conducted in the United States,^4,5,10,18,21,23,25,26,28,56–58,60^ six (27.3%) in the United Kingdom,^16,17,22,27,29,59^ and one each (4.5%) in Australia,^24^ Malaysia,^19^ and Denmark.^20^ Thirteen studies (59.1%) used randomized designs.^4,10,16,17,19,21–23,27,29,57–59^ Nine (40.9%) used non-randomized comparative designs, including pre-post studies, quasi-experimental comparisons, or sequential intervention phases.^5,18,20,24–26,28,56,60^ Most studies evaluated agenda-setting during adult outpatient encounters.^4,5,10,16–21,23,25–29,56–60^ Robling et al. (2012) included children with type 1 diabetes and their care partners.^22^ Two studies evaluated agenda-setting by patient-care partner dyads, one involving older adults with cognitive impairment^4^ and the other involving patients receiving treatment for breast cancer.^23^ One study evaluated psychiatry trainees using standardized patient encounters rather than observed clinic visits.^24^ Among studies enrolling patients, sample sizes ranged from 64 in Pritt (2020)^28^ to 3,124 patients in Hamilton et al. (2007).^17^ Several studies enrolled relatively few clinicians but evaluated multiple encounters per clinician, making the clinician, clinic, or practice the relevant allocation unit.^10,19,22,27,29,58^

**Table 1a.** Study and participant characteristics.

| Study | Location and clinical setting | Clinical population or visit type | Study period | Design and allocation unit | Participant eligibility | Patient, care partner, or encounter sample | Clinician sample |
| --- | --- | --- | --- | --- | --- | --- | --- |
| Brock et al. (2011)<sup>10</sup> | Puget Sound region, Washington, United States; 12 community primary care clinics | Established adult primary care patients | Physician recruitment July 2003 to October 2004, training and patient recruitment through March 2005 | Post-intervention only cluster RCT, physicians allocated to training or control | Adults who had seen the physician at least twice in two years, were English-speaking, acted as their own guardian, and had no serious cognitive impairment | 1,460 enrolled, including 796 intervention and 664 control patients | 28 intervention and 25 control physicians after losses |
| Deuter et al. (2023)<sup>60</sup> | South Texas, United States; outpatient psychiatric practice-based research network | Established adult psychiatric patients | N/R | Paired single-group pre-post study | English-speaking established psychiatric patients aged 18 to 75 years; severe cognitive impairment excluded | 141 enrolled, 138 included in some analyses after three exclusions | Treating psychiatrists; number N/R |
| Ditton-Phare et al. (2016)<sup>24</sup> | Newcastle, New South Wales, and Melbourne, Victoria, Australia; psychiatry vocational training program | Simulated encounters about schizophrenia diagnosis or prognosis | Training delivered during 2013-2014 | Pre-post standardized patient evaluation | Psychiatry trainees participating in ComPsych training | No clinical patients, standardized patient encounters | 30 psychiatry trainees; 25 contributed paired outcome data |
| Early et al. (2015)<sup>16</sup> | Cambridge, United Kingdom; respiratory outpatient clinic | Adults attending respiratory consultations | March to May 2014 | Individually randomized RCT preceded by a pilot | Adults on designated respiratory clinic lists who were literate and able to consent | 84 intervention and 81 control; 83 and 80 analyzed | Participating respiratory clinicians; number N/R |
| Haas et al. (2003)<sup>25</sup> | Utah, United States; university family medicine clinic | Adult English-speaking outpatients | N/R | One-group pre-post pilot | Adult English-speaking outpatients willing to have visits recorded | 36 pre-workshop and 29 post-workshop audiotaped encounters | Three family physicians |
| Haas et al. (2006)<sup>26</sup> | Utah, United States; seven community primary care clinics | Adult English-speaking primary care patients | N/R | Pre-post comparison using independent patient samples | Adult English-speaking patients of participating primary care clinicians | 301 pre-workshop and 322 post-workshop patients | 75 clinicians, including physicians and advanced practice clinicians |
| Hamilton et al. (2007)<sup>17</sup> | Devon and Dorset, United Kingdom; ten general practices | All patients attending during routine hours | June 2004 to June 2005 | Individually randomized RCT | All patients attending participating practices during routine hours; care partners could help complete the form | 3,124 randomized; prescribing or satisfaction outcomes available for 1,783, including 896 intervention and 887 control patients | 55 general practitioners |
| Kuhle et al. (2013)<sup>18</sup> | Mayo Clinic, Rochester, Minnesota, United States; occupational and executive health clinic in a tertiary center | Employees and employer-sponsored executive health patients | May 2011 | Quasi-experimental quality improvement comparison | Employees of the medical center or employer-sponsored executive health patients | 77 intervention and 36 control patients | N/R |
| Lee et al. (2022)<sup>19</sup> | University of Malaya Medical Centre, Kuala Lumpur, Malaysia; university-based primary care clinic | Follow-up visits for chronic disease | June to October 2019 | Unblinded cluster RCT, physicians were clusters | Chronic disease follow-up patients able to use the online tool; participating physicians were family medicine trainees | 110 intervention and 138 control; 101 and 131 included in principal adjusted analyses | 12 intervention and 13 control physicians |
| Leydon et al. (2018)<sup>27</sup> | Dorset, Hampshire, and Wiltshire, United Kingdom; general practices | Adult general practice consultations | Principal recordings August 2013 to March 2014 | Three-arm cluster-randomized feasibility trial, general practitioners allocated | Adults attending participating general practices; non-English-speaking patients excluded because the intervention depended on wording | 320 patients overall; between-arm comparison included 87 Some, 81 Any, and 110 usual care encounters | Seven general practitioners per arm, 21 total |
| Mauksch et al. (2001)<sup>58</sup> | United States; family medicine residency clinic at an urban academic medical center | Established adult primary care patients | July 1996 to July 1997 | Pilot comparative study with physician-level random assignment | Adult English-speaking patients who had previously seen the study physician | 82 intervention and 80 control encounters | Eight intervention and nine control physicians |
| McLean and Armstrong (2004)<sup>59</sup> | Southeast England, United Kingdom; four semi-rural training practices | Patients with minor self-limiting illness | N/R | Open individually randomized RCT | Patients with self-limiting illness; patients requiring referral, non-symptomatic prescriptions, or already expressing a clear concern were excluded | 56 intervention and 54 control patients | Participating general practitioners; number N/R |
| Middleton et al. (2006)<sup>29</sup> | Leicestershire and Nottinghamshire, United Kingdom; general practices | General practice consultations | N/R | Factorial study combining physician education with an embedded patient-level randomized agenda form trial | No patient exclusion criteria; participating physicians volunteered for the educational trial | 976 patients enrolled; complete physician-patient data available for 857 | 46 general practitioners; data available for 45 |
| Munch et al. (2024)<sup>20</sup> | Greater Copenhagen, Denmark; outpatient type 2 diabetes clinic | Annual 45-60 minute nurse-led status visits | April to August 2022 | Non-randomized concurrent comparison | Adults attending the type 2 diabetes clinic for at least six months who could read Danish | 52 intervention and 55 control patients; 20 intervention patients interviewed | Eight intervention and nine control diabetes nurses |
| O'Malley et al. (2022)<sup>21</sup> | Bethesda, Maryland, United States; adult primary care clinic | Established patients with hypertension and at least two additional chronic conditions | October 2011 to May 2012 | Pilot RCT | Adults with an established clinician relationship, hypertension, and at least two other chronic conditions; cognitive impairment and inability to complete English surveys excluded | 51 intervention and 69 control patients randomized; 106 completed the initial visit | 11 general internists |
| Pritt (2020)<sup>28</sup> | Rural Appalachian Pennsylvania; hospital-based family medicine and community primary care clinics | Established adult chronic-care visits | October 2019 to March 2020 | Non-randomized before-after comparison of usual care and intervention phases, physicians randomized | Adults attending continuing care for a chronic condition, able to complete the visit in English, and living in an Appalachian community | 50 control and 14 intervention patients; usable audio was available for 43 control encounters and one intervention encounter | Nine physicians |
| Robling et al. (2012)<sup>22</sup> | United Kingdom, 26 secondary and tertiary pediatric diabetes services | Children aged 4-15 years with type 1 diabetes and their care partners | N/R | Pragmatic cluster-randomized trial, pediatric diabetes centers were clusters | Children aged 4 to 15 years with type 1 diabetes for at least 12 months and a participating parent or care partner | 359 intervention and 334 control children; 288 and 287 parent or care partner respondents at baseline | 79 practitioners across 13 intervention teams; 13 control teams |
| Rodriguez et al. (2008)<sup>5</sup> | California, United States; primary care, orthopedics, and ophthalmology practices | Commercially insured and Medicare patients | Workshop October 2005, follow-up surveys through 2007 | Non-randomized controlled comparison of trained physicians with matched untrained physicians using repeated patient surveys | Lower-performing practices selected from a multispecialty organization and matched by survey performance, geography, specialty, and size | 894 intervention-practice and 1,187 comparison-practice survey respondents | 10 intervention and 11 matched comparison physicians |
| Sepucha et al. (2002)<sup>56</sup> | United States; surgical and medical oncology clinics | Patients with breast cancer consulting about treatment | October 1996 to March 1997 | Non-randomized sequential controlled trial | English-speaking patients with breast cancer consulting a surgeon or oncologist about treatment | 99 enrolled and 94 completed; 42 intervention and 52 comparison patients | Surgeons and oncologists at the participating centers |
| Talen et al. (2011)<sup>57</sup> | Chicago, Illinois, United States; outpatient family medicine | Adult primary care consultations | N/R | Individually randomized RCT | Adults aged 21 to 80 years able to read English at approximately sixth-grade level | 87 intervention and 121 control patients | 12 family medicine physicians |
| Wolff et al. (2018)<sup>4</sup> | Baltimore, Maryland, United States; two general primary care clinics and one geriatrics clinic | Adults aged 65 years or older with cognitive impairment attending with a care partner | N/R | Two-group pilot RCT | Adults aged 65 years or older with cognitive impairment attending with an unpaid companion | 49 intervention and 44 control patient-care partner dyads | 14 clinicians |
| Wolff et al. (2019)<sup>23</sup> | Baltimore, Maryland, United States; breast oncology clinics | Patients receiving active breast cancer treatment who regularly attended with a care partner | N/R | Two-arm single-blind pilot RCT | Patients receiving active breast cancer therapy and attending visits with an unpaid care partner | 69 intervention and 63 control patient-care partner dyads | Oncology clinicians; number N/R |

N/R indicates not reported. RCT indicates randomized controlled trial. Participant counts refer to the most relevant enrolled or analyzed sample and may vary across outcomes.

**Table 1b.** Study and participant characteristics.

| Study | Age | Sex or gender | Race, ethnicity, education, socioeconomic status, or health literacy | Attrition or missing data | Follow-up | Main outcome domains and measures |
| --- | --- | --- | --- | --- | --- | --- |
| Brock et al. (2011)<sup>10</sup> | Patients: 52.0 intervention, 53.6 control; physicians: 47.5 and 46.1 years | Patients: 54.5% and 59.0% female; physicians: 38.5% and 45.5% female | Patients: 81.4% and 85.5% White; other race and ethnicity categories reported; education and health literacy N/R | Thirteen encounters under three minutes excluded; five physicians left participating clinics | Immediate post-visit | Observed agenda-setting behaviors and number of concerns raised (coded audio recordings); visit duration (recorded visit times); overall patient satisfaction, participatory decision-making, autonomy support, trust, and health status (Mauksch satisfaction scale, Medical Outcomes Study Participatory Decision-Making Scale, Health Care Climate Questionnaire, Primary Care Assessment Survey trust subscale, 8-item Short Form Health Survey, and Patient Health Questionnaire measures); overall clinician satisfaction and relationship difficulty (post-visit questionnaire and Difficult Doctor-Patient Relationship Questionnaire) |
| Deuter et al. (2023)<sup>60</sup> | Mean 45 years | 58% female | 66% White non-Hispanic; 18% Hispanic; education, income, and health literacy N/R | Three participants excluded for missing information; some item-level denominators unclear | Immediate post-visit | Concerns addressed, overall satisfaction, ease, and helpfulness (study-specific patient post-visit Likert survey); psychiatrist-rated usefulness and satisfaction (study-specific post-visit survey) |
| Ditton-Phare et al. (2016)<sup>24</sup> | Median 32 years | 50% female | Race, ethnicity, and prior communication training incompletely reported | Five trainees did not contribute paired data | Same day to four weeks after training, depending on cohort | Agenda-setting, questioning, checking, empathic communication, and information organization (ComSkill coding of standardized patient interviews) |
| Early et al. (2015)<sup>16</sup> | 62.1 intervention, 59.9 control | 54.2% and 43.8% female | 96.0% and 97.5% White; education and health literacy N/R | One participant per arm excluded because one did not see a physician and one withdrew | Immediate post-consultation | Important issues raised and discussed and desired visit outcome (study-specific post-visit items); visit duration (stopwatch); confidence in managing the condition (study-specific item scored from 0 to 10) |
| Haas et al. (2003)<sup>25</sup> | Patient age N/R; physicians averaged five years after residency | Patient gender N/R; two of three physicians female | N/R | Recording failures and incomplete questionnaires; one physician withdrew and one left the clinic | Immediate post-visit | Agenda elicitation and negotiation, late or unresolved concerns, and visit duration (coded audiotapes); concerns addressed and patient satisfaction (study-specific six-item patient survey); clinician satisfaction and visit efficiency (study-specific five-item clinician evaluation) |
| Haas et al. (2006)<sup>26</sup> | 34 years pre-workshop, 33 post-workshop | 57.8% and 54.3% female, with gender missing for some participants | N/R | Independent pre-workshop and post-workshop patient samples; item-level missingness not fully described | Immediate post-visit | Patient-reported visit structuring, concerns addressed, satisfaction, and adequacy of visit time (study-specific 30-item post-visit questionnaire) |
| Hamilton et al. (2007)<sup>17</sup> | Mean 53 years | 38.7% male | N/R | Outcomes available for 1,783 of 3,124 randomized patients; 29 of 53 eligible physicians completed telephone interviews | Two and 12 weeks for adherence; one week for reattendance | Patient satisfaction (Medical Interview Satisfaction Scale); decision satisfaction (Satisfaction with Decision Questionnaire); prescribing and reattendance (practice records); adherence (structured telephone interviews) |
| Kuhle et al. (2013)<sup>18</sup> | N/R | N/R | N/R | Only returned questionnaires were analyzed | Immediate post-visit | Important concerns addressed (study-specific five-point Likert item); form helpfulness and intention to reuse it (study-specific post-visit items) |
| Lee et al. (2022)<sup>19</sup> | Patients: 55.6 intervention, 63.6 control; physicians: 33.4 and 33.2 years | Patients: 52.8% and 51.8% female; physicians: 33% and 24% female | Tertiary education: 64.2% and 50.7%; mobile-device proficiency measured; race and ethnicity N/R | One patient per arm did not complete the post-consultation questionnaire | Immediate post-visit | Agenda items discussed and visit duration (clinician report); patient-reported communication and overall patient satisfaction (30-item Healthcare Professional-Patient Communication questionnaire) |
| Leydon et al. (2018)<sup>27</sup> | 57.5 Some, 57.2 Any, 56.0 control | 55.2%, 61.7%, and 60.0% female | White ethnicity: 67.8%, 76.5%, and 73.6%; education and health literacy N/R | One withdrawal; outcome-specific missingness; unresolved denominator discrepancy for unvoiced concerns | Immediate post-consultation | Elicitation of additional concerns and intervention fidelity (coded video recordings); planned, discussed, and unvoiced concerns (pre- and post-visit questionnaires); visit duration (coded video recordings); overall patient satisfaction (21-item Medical Interview Satisfaction Scale and a single-item rating); patient enablement (Patient Enablement Instrument) |
| Mauksch et al. (2001)<sup>58</sup> | Patients: mean 42.2 years | Patients: 66.9% female; physicians: 50% and 66% female | N/R | One physician left the practice | Immediate post-visit and one-month reattendance | Elicitation, prioritization, concerns addressed, problems documented; patient and clinician satisfaction (study-specific immediate post-visit surveys); return visits (clinical records); visit duration (timer) |
| McLean and Armstrong (2004)<sup>59</sup> | N/R | N/R | N/R | No material attrition reported | Immediate post-visit | Visit duration (clinician-recorded time); overall patient satisfaction, satisfaction with clinician, and adequacy of visit time (Consultation Satisfaction Questionnaire and its subscales); patient enablement (Patient Enablement Instrument); anxiety (six-item short-form State-Trait Anxiety Inventory) |
| Middleton et al. (2006)<sup>29</sup> | Patient age N/R; most clinicians aged 40 to 44 years | Clinicians: 74% male in education group and 67% in comparison group | N/R | Complete data for 857 patients and 45 physicians | Immediate post-consultation | Problems identified and late-arising problems (clinician encounter form); consultation duration and time per problem (stopwatch); satisfaction, professional care, depth of relationship, and adequacy of visit time (Consultation Satisfaction Questionnaire subscales) |
| Munch et al. (2024)<sup>20</sup> | 67.6 intervention, 67.8 control | 60% and 75% male | Education reported by arm; others N/R | Figures implied several missing item responses; analytic denominators N/R by arm | Immediate survey; interviews one to five days later | Nurse effort to identify topics, topics discussed, and supportiveness and use of the Conversation Cards (study-specific 10-item post-visit survey); patient experience (semi-structured interviews) |
| O'Malley et al. (2022)<sup>21</sup> | 65.6 intervention, 66.5 control | 52.8% and 54.1% female | 57% Black and 36% White; health literacy measured; education and socioeconomic variables collected | Two participants per arm lost after the baseline encounter; 106 completed the initial visit | Immediate, four weeks and 12 weeks | Overall patient satisfaction (five-item Likert measure); trust (Primary Care Assessment Survey); physicians' perceived encounter difficulty (study-specific post-visit rating); observed patient activation (Roter Interaction Analysis System-based composite); shared decision-making (20-point patient rating); medication adherence (pill count and Morisky Medication Adherence Scale); functional status (Medical Outcomes Study six-item Short Form); blood pressure |
| Pritt (2020)<sup>28</sup> | 51.4 intervention, 54.5 control | 50% and 62% female | Intervention: 57.1% White, 35.7% Black; control: 88% White, 4% Black; education and income reported; health literacy N/R | Recordings were not obtained or were unusable for 21 visits; only one intervention recording was usable; two patient questionnaires were missing; recruitment ended early because of COVID-19 | Immediate post-visit | Agenda-setting and unmet concerns (coded audio recordings); patient-centeredness and relationship quality (Consultation Care Measure) |
| Robling et al. (2012)<sup>22</sup> | 10.4 intervention, 10.7 control | 52% and 46% male | Approximately 91% White British in both arms; socioeconomic status reported for parents; health literacy N/R | Five intervention and 11 control children withdrew; parent survey attrition occurred in both arms | 12 months | Occurrence of agenda-setting and consultation skills (coded audio recordings); glycated hemoglobin (HbA1c; capillary blood sample); quality of life (Pediatric Quality of Life Inventory Generic Core Scales and Type 1 Diabetes Module); patient enablement (Patient Enablement Instrument); healthcare climate, diabetes distress, continuity, and self-management (Health Care Climate Questionnaire, Problem Areas in Diabetes, Diabetes Continuity of Care Scale, and Diabetes Management Scale) |
| Rodriguez et al. (2008)<sup>5</sup> | Patients: 49.8 intervention, 48.5 comparison | 39% and 44% male | Intervention patients: 61% White, 11% Black, 17% Asian; comparison: 77% White, 2% Black, 4% Asian; education reported | Outcome-specific survey response and sample-composition differences; analyses adjusted for patient characteristics | Surveys before and after training, including longer-term follow-up | Physician-patient interaction quality, overall physician rating, willingness to recommend, organizational access, care coordination, and office staff interactions (abbreviated Ambulatory Care Experiences Survey) |
| Sepucha et al. (2002)<sup>56</sup> | 53 intervention, 52 comparison | N/R, participants were patients with breast cancer | 79% and 81% White non-Hispanic; 54% and 69% college graduates; median household income \$50,000 to \$70,000 | Five of 99 participants did not complete because the clinician was ready before the preparation session ended | Immediate post-consultation | Patient satisfaction (Satisfaction With Interview Scale and Patient-Doctor Interaction Scale); communication barriers (Community Breast Health Project scale); clinician satisfaction (Physician Satisfaction Scale) |
| Talen et al. (2011)<sup>57</sup> | Mean 48 years | 76% and 75% female | Intervention arm: 68% White, 28% Black, 3% Latino; comparison-arm race incompletely reported | N/R | Immediate post-visit | Patient and clinician ratings of patient knowledge, communication and organizational skills, and ownership and partnership attitudes (study-specific eight-item patient survey and parallel clinician survey) |
| Wolff et al. (2018)<sup>4</sup> | 80.9 intervention, 78.8 control | 49.0% and 54.5% female | Non-White race or Hispanic ethnicity: 36.7% and 47.7%; education beyond high school: 51.0% and 45.5% | No dyads lost from the immediate assessment | Immediate and two weeks | Patient-centered communication, verbal activity, information disclosure, and memory discussion (Roter Interaction Analysis System coding of audio recordings); visit duration (audio recordings); prioritized topics and care partner roles (SAME Page checklist); intervention acceptability (study-specific post-visit items) |
| Wolff et al. (2019)<sup>23</sup> | 55.1 intervention, 52.9 control | 98.6% and 100% female | Non-White race or Hispanic ethnicity: 24.6% and 33.3%; education beyond high school: 92.8% and 77.8% | One intervention patient died; portal outcomes had outcome-specific denominators | Six weeks, with longer follow-up planned | Patient-care partner agenda agreement (Sharing in Care checklist); visit satisfaction and intervention acceptability (study-specific post-visit items); care partner roles; patient-portal registration and use (time-stamped electronic health record audit trails) |

N/R indicates not reported. Participant counts refer to the most relevant enrolled or analyzed sample and may vary across outcomes.

### Intervention and delivery characteristics

The interventions varied considerably in their intended recipients, delivery, and level of structure. Seven studies used structured agenda-setting interventions that presented specific topics or topic categories for patients to select.^4,16,20–23,60^ Eight studies used unstructured interventions that elicited agenda items using a general prompt without proposing specific topics.^10,17–19,29,56,57,59^ The remaining seven studies evaluated only clinician training-based interventions without a patient-facing component.^5,24–28,58^

Patient-facing interventions were most often completed immediately before the clinical encounter. Delivery approaches included paper forms completed in the waiting room,^16–18,29,57^ digital tools completed before the visit,^19,60^ and checklists or cards completed by patients alone or with care partners or clinicians.^4,20,21,23^ Sepucha et al. (2002) used a 20-minute facilitated conversation in which a researcher helped the patient identify and organize questions before the visit.^56^ Clinician-facing interventions ranged from brief videos to longer workshops with practice or follow-up coaching. Pritt (2020) and Leydon et al. (2018) each used a five-minute video to train clinicians to elicit additional concerns using two different prompts: “do you have *any* other concerns?” versus “*some* other concerns?”^27,28^ Rodriguez et al. (2008) combined a three-hour workshop with two follow-up teleconferences,^5^ while Brock et al. (2011) used a two-hour workshop with individual coaching.^10^

Most interventions were evaluated against usual care or pre-intervention practice. Comparative designs included participant- or dyad-randomized parallel-group trials,^4,16,17,21,23,57,59^ cluster-randomized trials in which clinicians or clinical sites were allocated,^10,19,22,27,58^ one factorial trial,^29^ before-after or sequential phase studies,^24–26,28,56,60^ and other controlled comparisons.^5,18,20^ **Table 2** summarizes intervention and delivery characteristics, comparators, and fidelity or implementation assessment.

**Table 2.**
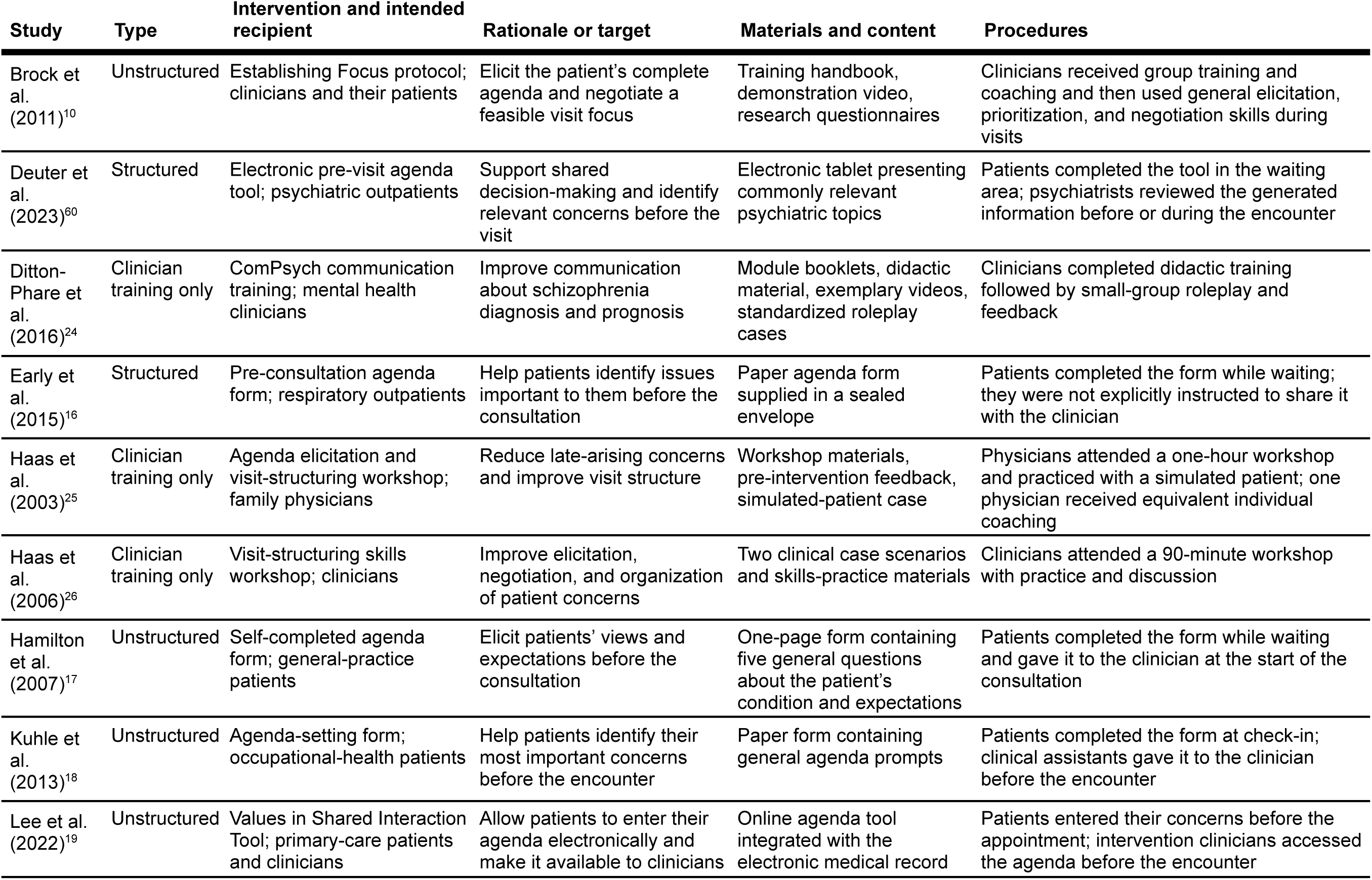
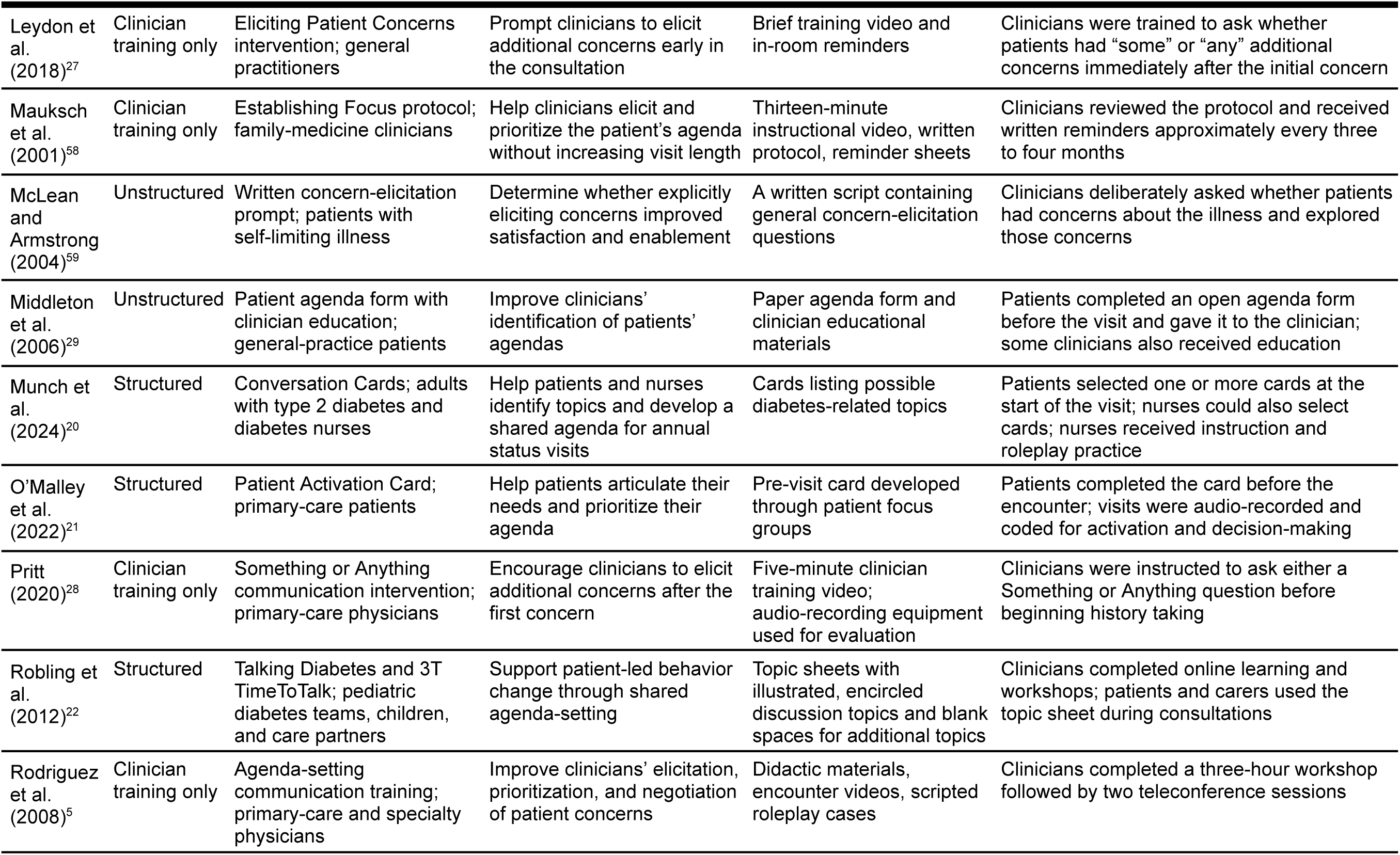
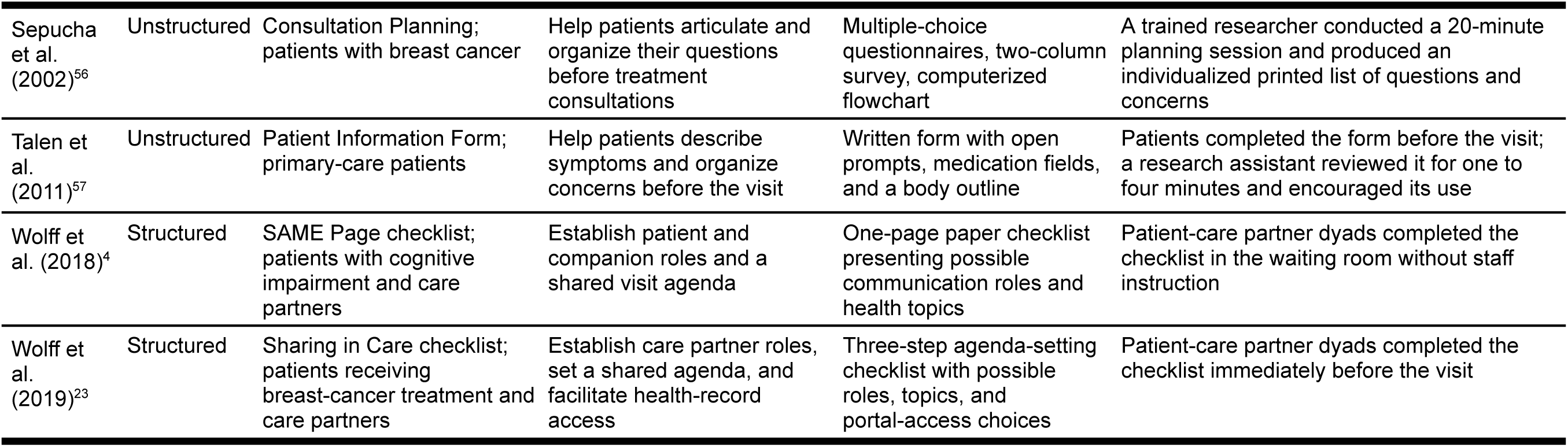
Intervention and delivery characteristics.

**Table 2b.** Intervention and delivery characteristics.

| Study | Deliverer | Mode and location | Timing and dose | Comparator | Tailoring or modification | Fidelity or implementation assessment |
| --- | --- | --- | --- | --- | --- | --- |
| Brock et al. (2011)<sup>10</sup> | Trainer and behavioral science coaches | In-person clinician training and workplace coaching | Two-hour group session followed by individual coaching | Usual care clinicians | Clinicians applied the protocol flexibly to individual encounters | Visits were audio-recorded; complete protocol adherence was not consistently reported |
| Deuter et al. (2023)<sup>60</sup> | Patient self-administered; psychiatrist reviewed results | Electronic tablet in the waiting area | Once immediately before the visit | Pre-intervention practice | Generated results reflected topics selected by each patient | Formal fidelity assessment N/R |
| Ditton-Phare et al. (2016)<sup>24</sup> | Trained facilitator | In-person didactic and small-group training | Approximately 30-45 minutes of didactic teaching followed by about two hours of roleplay | Pre-training performance | Roleplay adapted to diagnosis or prognosis scenarios | Communication was assessed using standardized encounters and coding |
| Early et al. (2015)<sup>16</sup> | Patient self-administered | Paper form in the waiting area | Once before the consultation | Sealed envelope with no paper form | Responses were individualized; patients decided whether to share the form | Completion was encouraged, but sharing with clinicians was not required |
| Haas et al. (2003)<sup>25</sup> | Study investigators | In-person workshop and individual coaching when needed | Approximately one hour | Pre-intervention encounters | One clinician unable to attend received individual instruction | Audiotaped encounters were coded by trained reviewers |
| Haas et al. (2006)<sup>26</sup> | Study investigators | In-person workshop | 90-minute session | Pre-intervention encounters | Practice used two standardized clinical cases | Formal fidelity assessment N/R |
| Hamilton et al. (2007)<sup>17</sup> | Patient self-administered | Paper form in general-practice waiting rooms | Once before the consultation | No agenda form | Patients or care partners could complete the form; clinicians could use it as they considered appropriate | Forms were not retained, preventing direct assessment of clinician use |
| Kuhle et al. (2013)<sup>18</sup> | Patient self-administered; clinical assistant transferred form | Paper form completed at check-in | Once before the encounter | Usual care | Patient supplied their own concerns | Form distribution and completion were reported; clinician use was not directly measured |
| Lee et al. (2022)<sup>19</sup> | Patient self-administered; clinician accessed results | Online tool linked to the electronic medical record | Once before the follow-up visit | Usual care | Free-text patient entries were individualized | Tool use could be identified electronically; detailed clinician use N/R |
| Leydon et al. (2018)<sup>27</sup> | Training video viewed by general practitioners | Video training with in-room reminders | Brief training before intervention consultations | Usual care and pre-training consultations | Clinicians were assigned to Some or Any wording | Correct delivery occurred in 75/87 Some and 70/80 Any consultations with fidelity data |
| Mauksch et al. (2001)<sup>58</sup> | Instructional video and research-team reminders | Video-based training in a family-medicine clinic | Thirteen-minute video with reminder sheets every three to four months | Control clinicians | Clinicians applied the protocol to individual visits | Formal adherence results N/R |
| McLean and Armstrong (2004)<sup>59</sup> | Treating clinician | Spoken prompt during the clinical encounter | Once after history and examination | Usual care | Follow-up questions depended on the concern expressed | Use of the assigned prompt was not separately quantified |
| Middleton et al. (2006)<sup>29</sup> | Patient self-administered; clinician education delivered by study team | Paper form in waiting area and one-day clinician workshop | Form once per study encounter; education once | No form, no clinician education, form only, or education plus form, according to factorial assignment | Patient entered their own concerns | Missing patient or clinician data varied across factorial groups |
| Munch et al. (2024)<sup>20</sup> | Patient and diabetes nurse | Physical cards used at the start of an annual status visit | Once per intervention visit; nurses also received instruction and monthly support meetings | Usual care | Patients and nurses selected cards relevant to the visit | Cards were used in 49/52 intervention visits; patients and nurses both selected cards in 43% of card-use visits |
| O'Malley et al. (2022)<sup>21</sup> | Patient self-administered | Paper card immediately before primary-care visit | One pre-visit exposure | Usual care | Patients selected and prioritized their own needs | Encounters were audio-recorded and rated in duplicate; tool-completion fidelity was not otherwise reported |
| Pritt (2020)<sup>28</sup> | Training video viewed by physicians | Video training in two primary-care practices | Approximately five minutes before the intervention phase | Pre-intervention usual care phase | Physicians were assigned to Something or Anything wording | Only one of 14 intervention encounters had usable audio |
| Robling et al. (2012)<sup>22</sup> | Trained facilitators and participating diabetes teams | Online learning and in-person workshops | Training delivered across approximately 12 months | Usual care teams | Clinicians could deploy a flexible menu of skills; sheets included blank topics | Process evaluation assessed training delivery and agenda-setting skills |
| Rodriguez et al. (2008)<sup>5</sup> | Two study investigators | In-person workshop and telephone follow-up | Three-hour workshop plus two 45-minute calls at 3 and 7 weeks | Matched comparison practices | Roleplay and discussion reflected clinicians' practice settings | Formal fidelity results N/R |
| Sepucha et al. (2002)<sup>56</sup> | Two trained researchers | Individual in-person session before the consultation | Approximately 20 minutes | Productive Listening | The printed flowchart reflected each patient's questions, priorities, and preferred order | Five of 99 enrolled patients did not complete the session because the clinician was ready |
| Talen et al. (2011)<sup>57</sup> | Research assistant | Paper form completed in the waiting room or examination area | One pre-visit form plus a one- to four-minute review | Usual care | Body outline and open fields were individualized | Formal fidelity assessment N/R |
| Wolff et al. (2018)<sup>4</sup> | Patient-care partner dyad | Paper checklist in the waiting room | Once immediately before the visit | Usual care | Dyads selected their preferred companion roles and topics | Formal fidelity assessment N/R |
| Wolff et al. (2019)<sup>23</sup> | Patient-care partner dyad | Paper checklist immediately before oncology visit | One pre-visit exposure | Usual care | Dyads selected roles, topics, and portal-access preferences | Checklist completion was documented; formal fidelity assessment N/R |

N/R indicates not reported.

Intervention fidelity and adherence were reported inconsistently. Munch et al. (2024) documented high use of Conversation Cards, or physical cards listing diabetes-related topics that patients and nurses could choose to develop a shared visit agenda.^20^ Leydon et al. (2018) reported high clinician adherence to the assigned concern elicitation prompt wording.^27^ Most other studies reported only whether intervention materials were delivered or completed,^16,18,19,21,60^ or did not report a formal fidelity assessment. Completion of an intervention form or tool did not necessarily indicate that the resulting agenda was communicated to the clinician or was used during the encounter.

### Operationalization and measurement of agenda-setting

There was substantial variation in how the studies defined agenda-setting and determined whether it had occurred. Some measured occurrence of agenda-setting as a single observed behavior.^22,25^ Others separately coded initial elicitation of concerns, elicitation of additional concerns, prioritization or negotiation of concerns, and visit structuring.^10,25,26^ Leydon et al. (2018) and Pritt (2020) focused more narrowly on whether the clinician used a specific prompt to solicit additional concerns after the patient raised their first concern.^27,28^ Agenda-setting was measured through direct observation of audio- or video-recorded encounters,^4,10,21,22,24,25,27,28^ patient-reported assessments of agenda-setting, concerns addressed, or visit communication,^16,18–20,26,60^ and clinician-reported assessments of agenda items discussed or visit processes.^19,25,58^ **Table 1** identifies the measure used for each construct in every study.

Patient-reported satisfaction with clinician was assessed using the Consultation Satisfaction Questionnaire professional-care and depth-of-relationship subscales, a study-specific clinician-capability item, the Primary Care Assessment Survey trust measure, or abbreviated Ambulatory Care Experiences Survey measures.^5,21,26,29,59^ Adequacy of visit time was assessed using the Consultation Satisfaction Questionnaire perceived-time subscale or a study-specific item.^26,29,59^ Patient activation or enablement was assessed using the Patient Enablement Instrument,^22,27,59^ confidence in managing one’s condition,^16^ patient knowledge and communication measures,^57^ or observer-coded activation.^21^ Other patient-reported outcomes included trust,^10,21^ anxiety,^59^ communication barriers,^56^ and intervention acceptability.^4,18,20,23,60^

Most communication, satisfaction, and encounter-related outcomes were measured during or immediately after the visit.^4,10,16,18–21,24–29,56–60^ Fewer studies assessed later clinical or behavioral outcomes, including reattendance,^17,58^ medication adherence,^17,21^ blood pressure,^21^ glycemic control or self-management,^22^ and patient-portal use.^23^ Care partner outcomes included patient-care partner agenda agreement, participation roles, and communication during the encounter.^4,23^ Clinician outcomes included overall clinician satisfaction,^10,21,25,56,58,60^ perceived visit efficiency,^25^ and relationship or encounter difficulty.^10,21^

### Effects of agenda-setting interventions

We conducted 13 quantitative syntheses (**Table 3**). Twelve included at least two studies with nonzero analytic weight. Our meta-analysis of prioritization of concerns included two studies but was effectively determined by one^26^ as the second study had zero variance.^10^ **Figure 2** presents forest plots for six selected outcomes: occurrence of agenda-setting, number of concerns raised, concerns addressed measured continuously, visit duration, overall patient satisfaction, and overall clinician satisfaction. **Appendix 4** presents forest plots for the remaining pooled outcomes. **Appendix 5** presents post hoc exploratory subgroup analyses by study design, adjustment status, and intervention structure.

**Figure 2.**
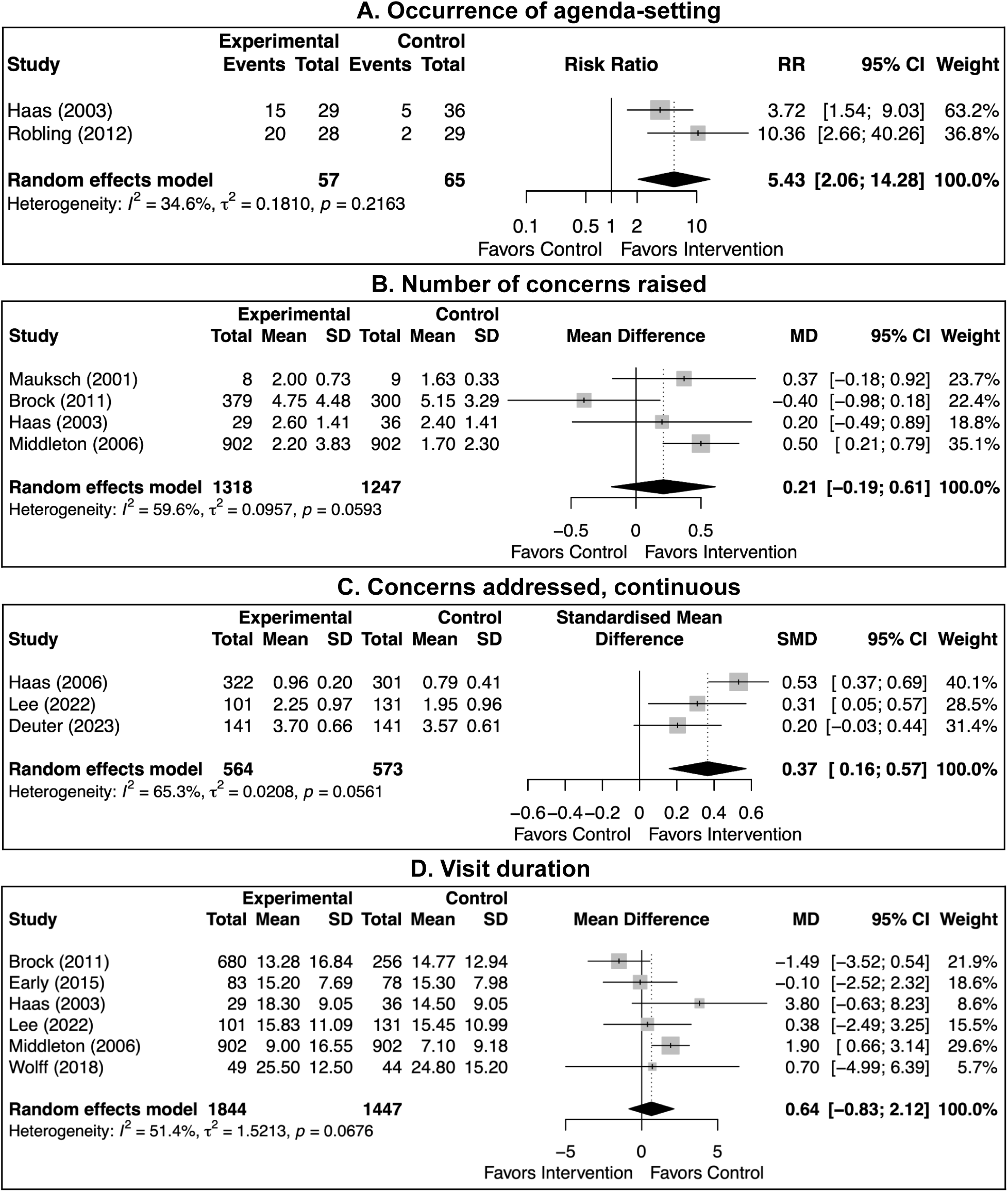
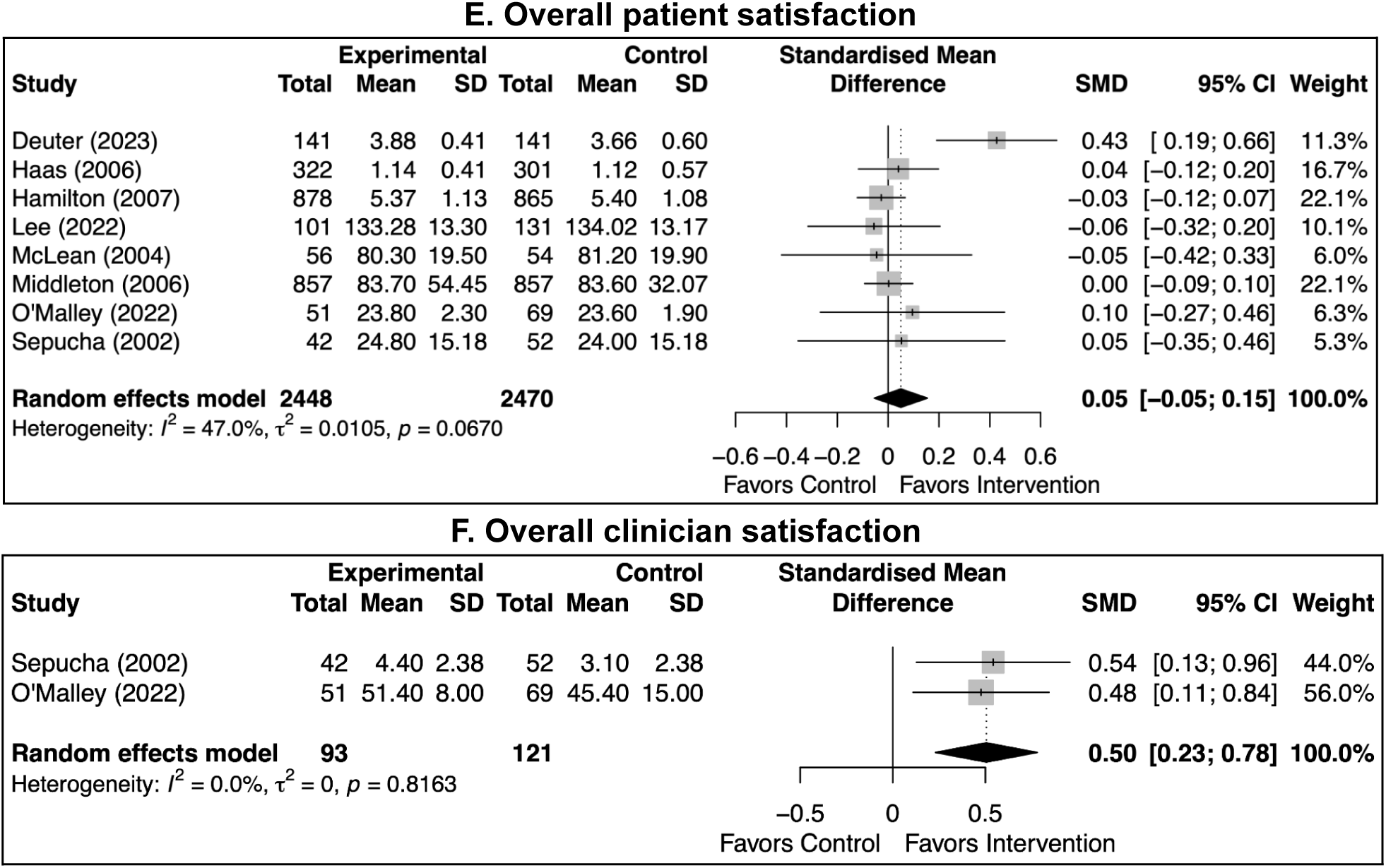
Effects of agenda-setting interventions on selected outcomes

**Table 3.**
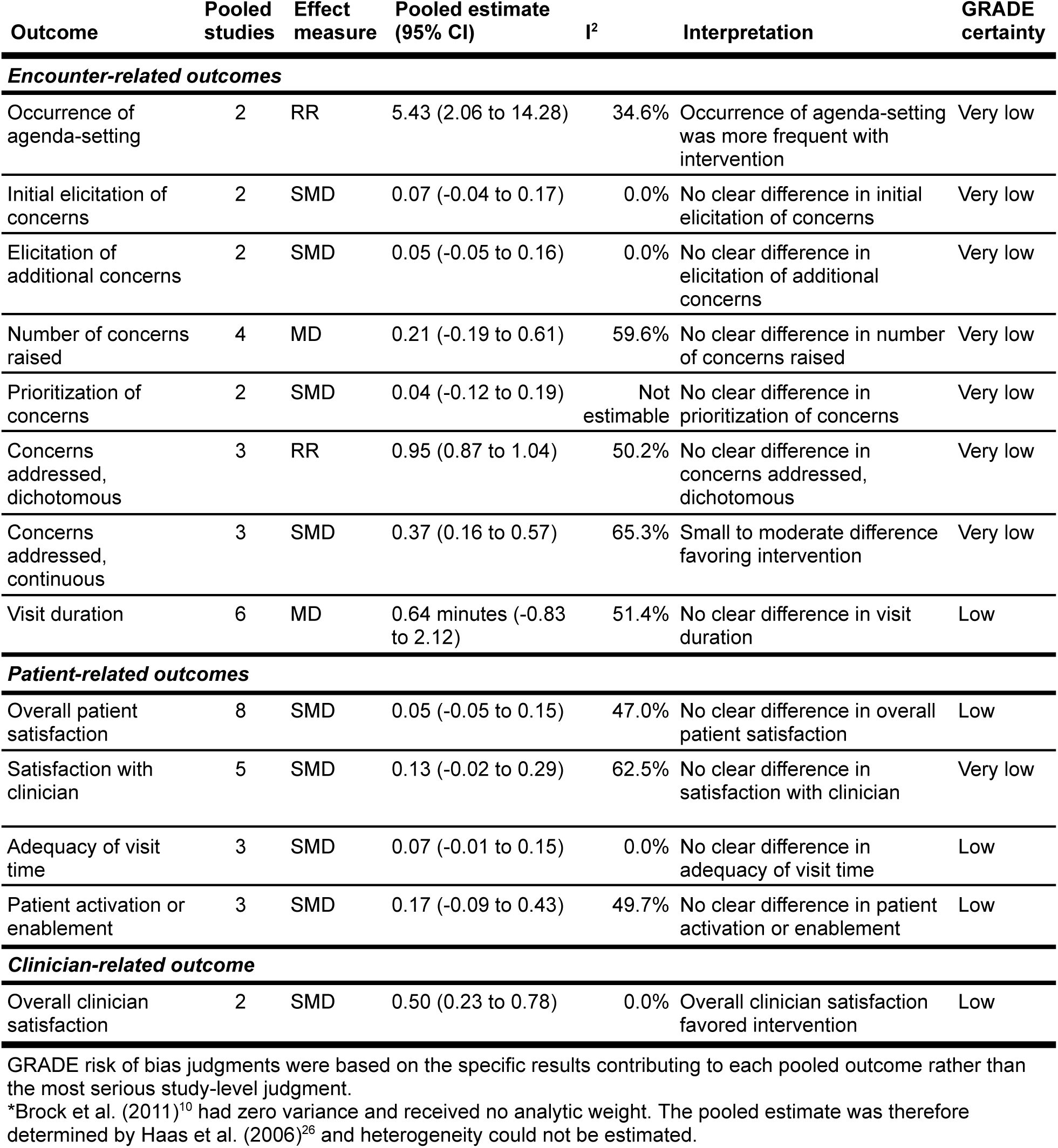
Meta-analysis estimates with GRADE certainty of evidence.

| Outcome | Pooled studies | Effect measure | Pooled estimate (95% CI) | I<sup>2</sup> | Interpretation | GRADE certainty |
| --- | --- | --- | --- | --- | --- | --- |
| **Encounter-related outcomes** |  |  |  |  |  |  |
| Occurrence of agenda-setting | 2 | RR | 5.43 (2.06 to 14.28) | 34.6% | Occurrence of agenda-setting was more frequent with intervention | Very low |
| Initial elicitation of concerns | 2 | SMD | 0.07 (-0.04 to 0.17) | 0.0% | No clear difference in initial elicitation of concerns | Very low |
| Elicitation of additional concerns | 2 | SMD | 0.05 (-0.05 to 0.16) | 0.0% | No clear difference in elicitation of additional concerns | Very low |
| Number of concerns raised | 4 | MD | 0.21 (-0.19 to 0.61) | 59.6% | No clear difference in number of concerns raised | Very low |
| Prioritization of concerns | 2 | SMD | 0.04 (-0.12 to 0.19) | Not estimable | No clear difference in prioritization of concerns | Very low |
| Concerns addressed, dichotomous | 3 | RR | 0.95 (0.87 to 1.04) | 50.2% | No clear difference in concerns addressed, dichotomous | Very low |
| Concerns addressed, continuous | 3 | SMD | 0.37 (0.16 to 0.57) | 65.3% | Small to moderate difference favoring intervention | Very low |
| Visit duration | 6 | MD | 0.64 minutes (-0.83 to 2.12) | 51.4% | No clear difference in visit duration | Low |
| **Patient-related outcomes** |  |  |  |  |  |  |
| Overall patient satisfaction | 8 | SMD | 0.05 (-0.05 to 0.15) | 47.0% | No clear difference in overall patient satisfaction | Low |
| Satisfaction with clinician | 5 | SMD | 0.13 (-0.02 to 0.29) | 62.5% | No clear difference in satisfaction with clinician | Very low |
| Adequacy of visit time | 3 | SMD | 0.07 (-0.01 to 0.15) | 0.0% | No clear difference in adequacy of visit time | Low |
| Patient activation or enablement | 3 | SMD | 0.17 (-0.09 to 0.43) | 49.7% | No clear difference in patient activation or enablement | Low |
| **Clinician-related outcome** |  |  |  |  |  |  |
| Overall clinician satisfaction | 2 | SMD | 0.50 (0.23 to 0.78) | 0.0% | Overall clinician satisfaction favored intervention | Low |

GRADE risk of bias judgments were based on the specific results contributing to each pooled outcome rather than the most serious study-level judgment.
\*Brock et al. (2011)<sup>10</sup> had zero variance and received no analytic weight. The pooled estimate was therefore determined by Haas et al. (2006)<sup>26</sup> and heterogeneity could not be estimated.

Agenda-setting interventions increased the **occurrence of agenda-setting**. Across Haas et al. (2003) and Robling et al. (2012), participants receiving an intervention were more than five times as likely to experience observed agenda-setting as those receiving comparison care, although the confidence interval was wide (RR 5.43, 95% CI 2.06 to 14.28, I^2^=34.6%).^22,25^ However, the pooled estimate did not show a clear difference in the **initial elicitation of concerns** (SMD 0.07, 95% CI −0.04 to 0.17, I^2^=0.0%) or the **elicitation of additional concerns** (SMD 0.05, 95% CI −0.05 to 0.16, I^2^=0.0%).^10,26^

The interventions did not clearly affect the **number of concerns raised** (MD 0.21 concerns, 95% CI −0.19 to 0.61, I^2^=59.6%)^10,25,29,58^ or the **prioritization of concerns** (SMD 0.04, 95% CI −0.12 to 0.19).^10,26^ Two studies were included in the latter synthesis, but Brock et al. (2011) had zero variance and supplied no analytic weight.^10^ The estimate was therefore determined by Haas et al. (2006) and heterogeneity could not be estimated.^26^

When **concerns addressed** was defined **dichotomously** as whether all or important concerns were addressed, the pooled estimate did not show a clear difference between intervention and comparison groups (RR 0.95, 95% CI 0.87 to 1.04, I^2^=50.2%).^16,18,25^ When **concerns addressed** were measured **continuously**, agenda-setting interventions were favored, with a small to moderate difference between groups (SMD 0.37, 95% CI 0.16 to 0.57, I^2^=65.3%).^19,26,60^ The heterogeneity suggests that the magnitude of the difference varied across studies or measurement approaches.

Agenda-setting interventions did not clearly affect **visit duration**. Across six studies, intervention visits were an estimated 0.64 minutes longer than comparison visits, with a confidence interval compatible with visits being shorter or longer (MD 0.64 minutes, 95% CI −0.83 to 2.12, I^2^=51.4%).^4,10,16,19,25,29^ In our exploratory subgroup analysis by intervention structure, the pooled MD was 0.38 minutes (95% CI −1.75 to 2.52) for unstructured interventions and 0.02 minutes (95% CI −2.21 to 2.25) for structured interventions. The interaction test did not provide evidence of a difference between intervention structures (p=0.82).

The interventions did not clearly affect **overall patient satisfaction** (SMD 0.05, 95% CI −0.05 to 0.15, I^2^=47.0%) across eight studies.^17,19,21,26,29,56,59,60^ In our subgroup analysis by intervention structure, the pooled SMD was −0.01 (95% CI −0.08 to 0.05, I^2^=0.0%) for unstructured interventions and 0.29 (95% CI −0.03 to 0.61, I^2^=55.6%) for structured interventions. The interaction test did not provide clear evidence of a difference between intervention structures (p=0.07). There was also no clear difference in patient-reported **satisfaction with clinician** (SMD 0.13, 95% CI −0.02 to 0.29, I^2^=62.5%).^5,21,26,29,59^ In our subgroup analysis by intervention structure, the pooled SMD was 0.24 (95% CI −0.23 to 0.71) for unstructured interventions and 0.39 (95% CI 0.02 to 0.76) for structured interventions. There was no evidence of a difference between intervention structures (p=0.61).

There were no clear differences in patient-reported **adequacy of visit time** (SMD 0.07, 95% CI −0.01 to 0.15, I^2^=0.0%)^26,29,59^ or **patient activation or enablement** (SMD 0.17, 95% CI −0.09 to 0.43, I^2^=49.7%).^16,57,59^ Subgroup analysis of activation or enablement by intervention structure found a pooled SMD of 0.26 (95% CI −0.06 to 0.58) for unstructured interventions and 0.00 (95% CI −0.31 to 0.31) for the structured intervention, with an interaction test indicating no evidence of a difference between structures (p=0.25).

**Overall clinician satisfaction** favored agenda-setting interventions (SMD 0.50, 95% CI 0.23 to 0.78, I^2^=0.0%).^21,56^ Our subgroup analysis by intervention structure produced an SMD of 0.54 (95% CI 0.13 to 0.96) for the unstructured intervention and 0.48 (95% CI 0.11 to 0.84) for the structured intervention. The interaction test did not provide evidence of a difference between intervention structures (p=0.82).

### Narrative synthesis of outcomes not pooled

Several additional encounter-related outcomes could not be pooled. Mauksch et al. (2001) reported greater collaborative prioritization in the intervention group than in the comparison (mean percentage answering yes, 93.75% versus 84.92%, standardized effect size 0.92) and more documented follow-up requests (mean percentage of encounters, 25.9% versus 8.9%; standardized effect size, 1.04).^58^ These results were reported as physician-level percentages and study-specific standardized effect sizes that were not compatible with the corresponding pooled estimates.^58^ Munch et al. (2024) found no statistically significant differences between Conversation Cards and usual care in patients’ ratings of nurses’ efforts to identify relevant topics or whether patients discussed the topics they wanted to discuss.^20^

Leydon et al. (2018) was not included in the meta-analyses because it reported descriptive results from a cluster-randomized three-arm feasibility trial without cluster-adjusted pairwise effect estimates.^27^ Concern outcomes were reported using categorical or study-specific comparisons. The mean number of concerns discussed did not clearly differ between groups.^27^ Mean visit duration was 10.2 minutes with usual care, 11.6 minutes with “Some,” and 10.2 minutes with “Any,” but a compatible measure of dispersion was unavailable.^27^

Mauksch et al. (2001) reported higher overall patient satisfaction in the intervention group than in the comparison group (mean 9.5 versus 9.0; standardized effect size, 0.85), but the result was reported as a physician-level study-specific standardized effect that was not compatible with the pooled patient-level measures.^58^ O’Malley et al. (2022) reported greater observed patient activation during intervention encounters than during usual care encounters (mean 4.4 versus 3.8 on a 1 to 10 scale, p=0.047), but no measure of dispersion was reported for inclusion in the patient activation or enablement meta-analysis.^21^ O’Malley et al. did not identify clear differences in shared decision-making or trust.^21^

Leydon et al. (2018) reported mean Medical Interview Satisfaction Scale scores of 102.52 with usual care, 102.78 with “Some,” and 101.22 with “Any.”^27^ Mean Patient Enablement Instrument scores were 4.07, 5.08, and 3.70, respectively. These patient-reported outcomes were not pooled because compatible pairwise effect estimates and variances were unavailable.^27^ Robling et al. (2012) did not identify a clear improvement in HbA1c.^22^ The adjusted between-group coefficient for log-transformed HbA1c was 0.01 (95% CI −0.02 to 0.04).^22^ O’Malley et al. (2022) found no clear effect on medication adherence or blood pressure^21^ and Hamilton et al. (2007) found no clear differences in prescribing, medication cost, or medication adherence.^17^

For care-partner-related outcomes, Wolff et al. (2018) found that SAME Page visits contained a higher proportion of patient-centered communication than control visits (mean ratio 0.86 versus 0.68; p=0.046).^4^ We did not quantitatively synthesize other care-partner outcomes because their constructs and measures were not sufficiently comparable. For additional clinician-related outcomes, Mauksch et al. (2001) found no clear differences in clinician satisfaction, the proportion of scheduled appointment time used, or patient return within one month.^58^

### Risk of bias

**Figures 3** and **4** present the overall and domain-level risk of bias summaries for the included studies. Of the 13 randomized studies assessed with RoB 2, only one had low overall risk of bias.^4^ Seven studies raised some concerns overall,^10,16,21,23,29,58,59^ and five had high overall risk of bias based on their most serious assessed result.^17,19,22,27,57^ For Robling et al. (2012), the high risk assessment applied to the agenda-setting process result, while its other patient and clinical outcomes only raised some concerns.^22^ Common concerns included incomplete reporting of sequence generation or allocation concealment, recruitment after cluster assignment, missing outcome data, and subjective outcomes reported by participants who knew which intervention they received.

**Figure 3.**
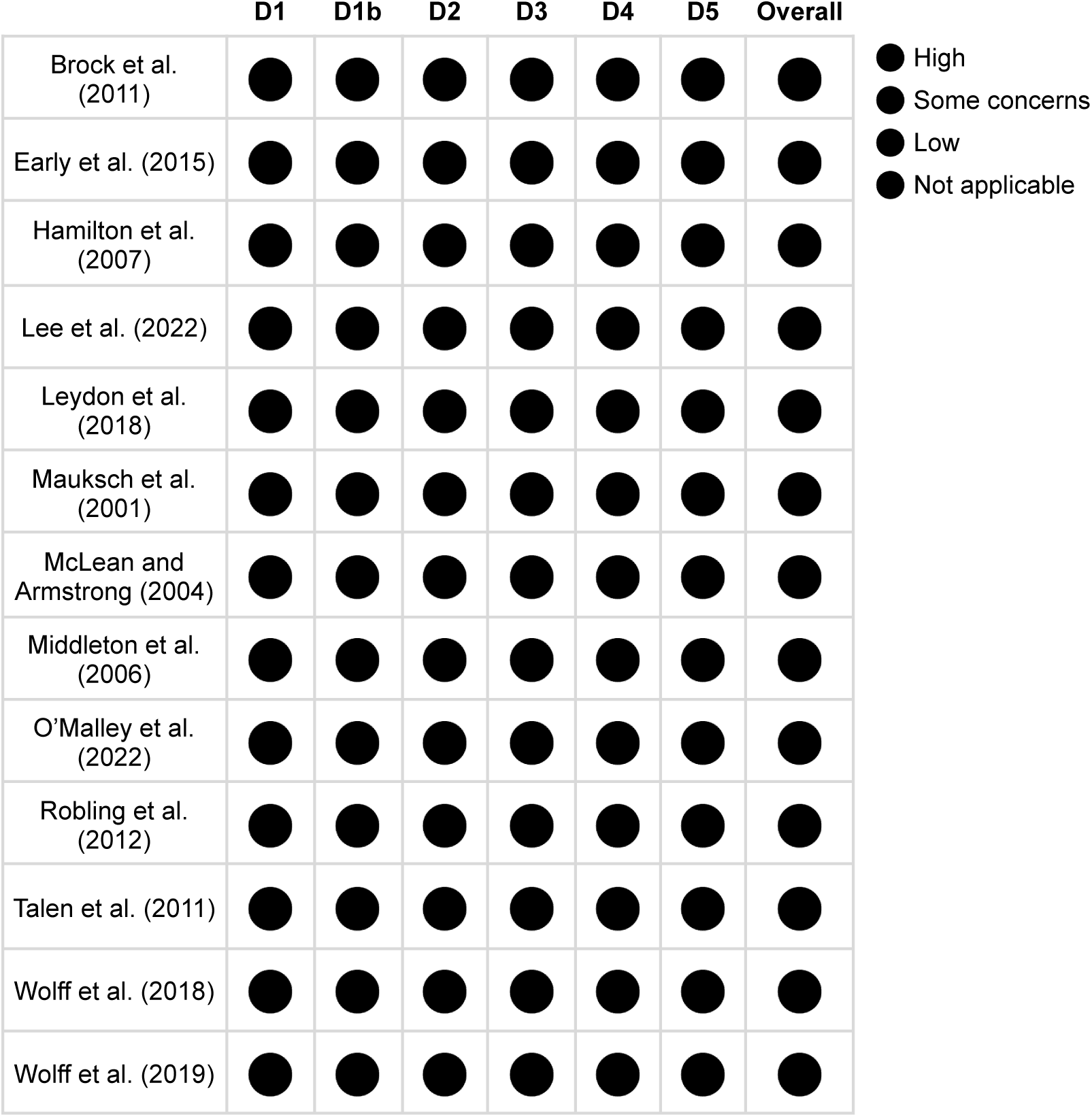
Traffic light plot of RoB 2 assessments of randomized studies

**Figure 4.**
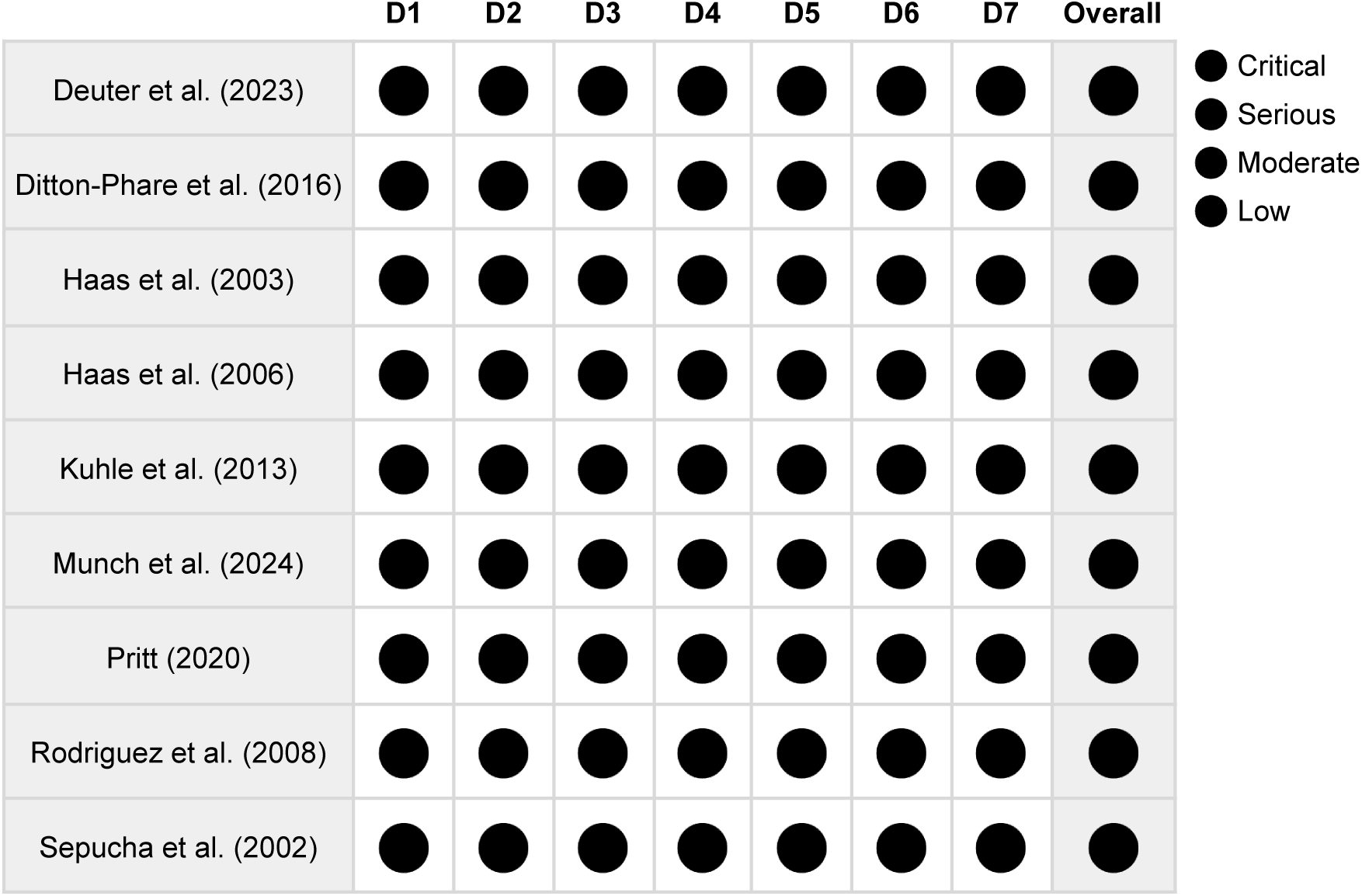
Traffic light plot of ROBINS-I assessments of non-randomized studies

Among the nine non-randomized studies assessed with ROBINS-I, five had serious overall risk of bias^5,18,20,24,56^ and three had critical risk of bias.^25,26,60^ Pritt (2020) had critical risk of bias for its audio-coded agenda-setting outcomes and serious risk for its patient-centered communication questionnaire outcomes.^28^ Frequent concerns included uncontrolled confounding, non-randomized selection into intervention groups, outcome measurement by unblinded participants or assessors, and incomplete or selectively reported results.

### Reporting bias and small-study effects

We generated funnel plots for eight outcomes with at least three contributing studies: number of concerns raised, concerns addressed (dichotomous), concerns addressed (continuous), visit duration, overall patient satisfaction, satisfaction with clinician, adequacy of visit time, and patient activation or enablement (**Appendix 6**). Visual inspection did not identify a consistent pattern of small-study asymmetry. The assessments for visit duration, overall patient satisfaction, and satisfaction with clinician did not show clear directional asymmetry, although estimates were dispersed. The remaining assessments included fewer studies and were too sparse for reliable interpretation. The assessments for the number of concerns raised and concerns addressed (continuous) appeared uneven, with less precise estimates tending to show smaller or less favorable intervention effects. However, chance and between-study heterogeneity were plausible explanations. These assessments did not provide reliable conclusions about small-study effects or publication bias. As no meta-analysis included at least 10 studies, we did not conduct Egger’s regression.

Several articles described pilot or feasibility studies or otherwise enrolled small samples.^4,21,23–25,27,58^ Pritt (2020) also stopped recruitment early because of the COVID-19 pandemic.^28^ Together with incomplete outcome reporting and the absence of prespecified analysis plans in several articles, this increased concern about selective non-reporting.

### Certainty of evidence

The certainty of evidence assessed using GRADE was low or very low for every pooled outcome (**Table 3**). No outcomes were supported by high- or moderate-certainty evidence. Certainty was reduced most frequently because of risk of bias in the contributing results, inconsistency across studies, imprecision, and differences in how outcome constructs were operationalized.

Evidence for occurrence of agenda-setting was of very low certainty as the two contributing results had critical or high risk of bias, and the pooled estimate had a wide confidence interval.^22,25^ Evidence for concerns addressed (continuous) was also of very low certainty as the contributing results had critical or high risk of bias, the synthesis had substantial heterogeneity, and outcome measurement varied across studies.^19,26,60^ Evidence for visit duration was of low certainty because the pooled estimate was imprecise and included results with methodological limitations.^4,10,16,19,25,29^ Evidence for overall patient satisfaction was of low certainty because the pooled estimate was imprecise and included results with methodological limitations.^17,19,21,26,29,56,59,60^ Evidence for overall clinician satisfaction was of low certainty because only two studies using different designs and measures contributed; one raised some concerns and the other had serious risk of bias.^21,56^

### Exploratory subgroup and sensitivity analyses

Our post hoc exploratory subgroup analyses examined study design, adjustment status, and intervention structure. We excluded interventions only using clinician training from analyses of intervention structure. Interaction tests were used as the primary evidence of subgroup differences. We present detailed pooled estimates and interaction tests in **Appendix 5**.

Interaction tests did not provide evidence that effects differed between randomized and non-randomized studies for any outcome with an estimable comparison. The interaction for prioritization of concerns could not be estimated as Brock et al. (2011) supplied no analytic weight.^10^ Interaction tests also did not provide evidence that effects differed according to adjustment status. The intervention structure analyses, reported alongside the corresponding pooled outcomes above, did not provide evidence that structured and unstructured interventions differed in effectiveness. However, these analyses included few studies and were not designed to establish equivalence.

We did not conduct sensitivity analyses excluding results at high, serious, or critical risk of bias because few studies contributed to each outcome and removing these results would have left too little evidence for informative comparisons.

## Discussion

To our knowledge, this is the first systematic review and meta-analysis focused specifically on interventions for collaborative clinical visit agenda-setting. Across 29 articles describing 22 unique studies, interventions included structured topic menus, unstructured prompts, and clinician training only, reflecting longstanding variation in how agenda-setting has been defined and operationalized.^1,6,14^ Interventions increased occurrence of agenda-setting,^22,25^ concerns addressed measured continuously,^19,26,60^ and overall clinician satisfaction.^21,56^ No clear differences were found for the initial elicitation of concerns,^10,26^ elicitation of additional concerns,^10,26^ number of concerns raised,^10,25,29,58^ prioritization of concerns,^10,26^ concerns addressed measured dichotomously,^16,18,25^ visit duration,^4,10,16,19,25,29^ overall patient satisfaction,^17,19,21,26,29,56,59,60^ satisfaction with clinician,^5,21,26,29,59^ patient-reported adequacy of visit time,^26,29,59^ or patient activation or enablement.^16,57,59^ The review was strengthened by a published protocol, a broad search without language or date restrictions, duplicate screening, linkage of companion articles, detailed extraction, analyses, and assessment of risk of bias and certainty of evidence. Even so, few studies contributed to most outcomes, several studies were small or non-randomized, and many results had substantial risk of bias. Evidence for all pooled outcomes was of low or very low certainty.

Our findings on the effects of agenda-setting interventions are consistent with agenda-setting first acting on the organization and completeness of an encounter, while behavior, satisfaction, and clinical outcomes are downstream of communication and care processes.^7,61^ The favorable finding for the overall occurrence of agenda-setting and the absence of clear differences in narrowly coded elicitation behaviors are not necessarily contradictory. Broad agenda-setting may incorporate elicitation, organization, prioritization, and negotiation, while narrower measures may only capture a specifically-phrased question.^2,6,62^ Similarly, continuous measures of concerns addressed may detect incremental improvement that is obscured when measured dichotomously according to whether all or important concerns were addressed. Interpretation of patient-reported outcomes is further limited by measurement heterogeneity. Overall patient satisfaction, satisfaction with clinician, adequacy of visit time, and patient activation or enablement were assessed using different scales and time points. Global satisfaction measures may also be particularly insensitive to changes in one component of communication.^63,64^

Other studies of communication interventions indicate a similar tendency for such interventions to influence proximal processes more consistently than more distal ones. Reviews of question prompt lists generally report increases in patient question or information exchange, but inconsistent effects on patient satisfaction and consultation length.^65–70^ Their effectiveness may depend on whether clinicians endorse and directly incorporate the tool during the consultation, rather than it being used before the encounter.^71,72^ A Cochrane review of 209 patient decision aid studies found improvements in knowledge, risk perception, values-choice congruence, and participation, while effects on consultation length depended on whether the decision aid was used before or during the visit.^73^ These related findings may support focusing evaluation on the mechanisms most directly targeted by each intervention, and in the case of agenda-setting, outcomes explicitly focused on communication.

While agenda-setting identifies which topics require attention, shared decision-making additionally requires deliberation about reasonable options and integration of evidence with patient preferences.^74–76^ Though agenda-setting may provide an upstream foundation for using patient-centered communication in shared decision-making by identifying what matters to patients, the present evidence does not establish that it improves subsequent deliberation.

O’Malley et al. (2022) illustrated this distinction in observing improved patient activation without a clear improvement in shared decision-making.^21^ Future research should also assess whether patients felt heard and understood during the encounter using measures specifically designed for these experiences rather than relying on just general satisfaction.^77–80^

Our exploratory subgroup analyses did not provide evidence that effects varied by study design, adjustment status, or intervention structure. These analyses included few studies and the absence of a statistically significant interaction should not be interpreted as evidence of equivalence.^81,82^ Structured agenda-setting menus may help patients raise concerns they might not otherwise think to or feel comfortable mentioning, while unstructured prompts may allow patients to define concerns on their own terms. Both approaches still depend on reliable delivery, clinician receipt, and use during the encounter, with the included studies reporting substantial variation in adherence.^83,84^ A completed agenda-setting tool therefore cannot be treated as the equivalent of collaborative agenda-setting having necessarily occurred.

Most evidence came from adult outpatient care in the United States and United Kingdom, limiting generalizability to other health systems, pediatric care, multilingual encounters, and serious illness. Serious illness visits often include particularly complex medical, emotional, practical, and family concerns, making agenda-setting especially relevant.^85–87^ Emerging tools such as Serious Illness Topics illustrate how agenda-setting can be integrated with discussion of patient priorities and values.^85,88^ Future research should assess the use of agenda-setting interventions across additional clinical contexts.

While we have not established that agenda-setting has no time cost in all settings, our findings support it as a reasonably low-burden communication practice that does not increase already limited visit time. Agenda-setting training should emphasize open elicitation of concerns, allowing patients to communicate their initial concerns, asking about additional concerns, and collaboratively prioritizing which to address.^2,37,62,89,90^ Evidence would be strengthened by adequately powered randomized studies that directly compare intervention formats and evaluate their implementation across diverse clinical settings.

## Conclusion

Clinical visit agenda-setting interventions may increase the occurrence of agenda-setting and the extent to which concerns are addressed, while pooled estimates did not show clear differences in visit duration or patient-reported outcomes. The apparent benefit for overall clinician satisfaction was based on only two studies, and certainty was low or very low for all pooled outcomes. Agenda-setting therefore appears to be a potentially low-burden communication practice, but stronger trials are needed to determine how it should be implemented across clinical settings.

## Data Availability

All data produced in the present study are available upon reasonable request to the authors.

## Endnotes

## Acknowledgements

We thank Heather Blunt and Elaina Vitale for their assistance in developing the search strategy.

## Funding

This review received no specific grant from any funding agency in the public, commercial, or not-for-profit sectors. Meredith A. MacMartin’s time was supported by the National Center for Advancing Translational Sciences (NCATS; K12TR004987). Catherine H. Saunders’ time was supported by the National Institute of Diabetes and Digestive and Kidney Diseases (NIDDK; 1K01DK139400). The funders had no role in the review design, conduct, analysis, interpretation, manuscript preparation, or decision to submit the manuscript.

## Competing interests

Ailyn Sierpe, Anne E. Dade, and Catherine H. Saunders have developed or studied clinical visit agenda-setting interventions. All other authors declare no competing interests.

## Appendix 1. PRISMA checklist

**Appendix 1 Table 1.** Preferred Reporting Items for Systematic Reviews and Meta-Analyses 2020 checklist^1^.

| Section and topic | No. | Checklist item |
| --- | --- | --- |
| **Title** |  |  |
| Title | 1 | Identify the report as a systematic review. |
| **Abstract** |  |  |
| Abstract | 2 | See the PRISMA 2020 for Abstracts checklist. |
| **Introduction** |  |  |
| Rationale | 3 | Describe the rationale for the review in the context of existing knowledge. |
| Objectives | 4 | Provide an explicit statement of the objective(s) or question(s) the review addresses. |
| **Methods** |  |  |
| Eligibility criteria | 5 | Specify the inclusion and exclusion criteria for the review and how studies were grouped for the syntheses. |
| Information sources | 6 | Specify all databases, registers, websites, organisations, reference lists and other sources searched or consulted to identify studies. Specify the date when each source was last searched or consulted. |
| Search strategy | 7 | Present the full search strategies for all databases, registers and websites, including any filters and limits used. |
| Selection process | 8 | Specify the methods used to decide whether a study met the inclusion criteria of the review, including how many reviewers screened each record and each report retrieved, whether they worked independently, and if applicable, details of automation tools used in the process. |
| Data collection process | 9 | Specify the methods used to collect data from reports, including how many reviewers collected data from each report, whether they worked independently, any processes for obtaining or confirming data from study investigators, and if applicable, details of automation tools used in the process. |
| Data items | 10a | List and define all outcomes for which data were sought. Specify whether all results that were compatible with each outcome domain in each study were sought (e.g. for all measures, time points, analyses), and if not, the methods used to decide which results to collect. |
|  | 10b | List and define all other variables for which data were sought (e.g. participant and intervention characteristics, funding sources). Describe any assumptions made about any missing or unclear information. |
| Study risk of bias assessment | 11 | Specify the methods used to assess risk of bias in the included studies, including details of the tool(s) used, how many reviewers assessed each study and whether they worked independently, and if applicable, details of automation tools used in the process. |
| Effect measures | 12 | Specify for each outcome the effect measure(s) (e.g. risk ratio, mean difference) used in the synthesis or presentation of results. |
| Synthesis methods | 13a | Describe the processes used to decide which studies were eligible for each synthesis (e.g. tabulating the study intervention characteristics and comparing against the planned groups for each synthesis (item #5)). |
|  | 13b | Describe any methods required to prepare the data for presentation or synthesis, such as handling of missing summary statistics, or data conversions. |
|  | 13c | Describe any methods used to tabulate or visually display results of individual studies and syntheses. |
|  | 13d | Describe any methods used to synthesize results and provide a rationale for the choice(s). If meta-analysis was performed, describe the model(s), method(s) to identify the presence and extent of statistical heterogeneity, and software package(s) used. |
|  | 13e | Describe any methods used to explore possible causes of heterogeneity among study results (e.g. subgroup analysis, meta-regression). |
|  | 13f | Describe any sensitivity analyses conducted to assess robustness of the synthesized results. |
| Reporting bias assessment | 14 | Describe any methods used to assess risk of bias due to missing results in a synthesis (arising from reporting biases). |
| Certainty assessment | 15 | Describe any methods used to assess certainty (or confidence) in the body of evidence for an outcome. |
| **Results** |  |  |
| Study selection | 16a | Describe the results of the search and selection process, from the number of records identified in the search to the number of studies included in the review, ideally using a flow diagram. |
|  | 16b | Cite studies that might appear to meet the inclusion criteria, but which were excluded, and explain why they were excluded. |
| Study characteristics | 17 | Cite each included study and present its characteristics. |
| Risk of bias in studies | 18 | Present assessments of risk of bias for each included study. |
| Results of individual studies | 19 | For all outcomes, present, for each study: (a) summary statistics for each group (where appropriate) and (b) an effect estimate and its precision (e.g. confidence/credible interval), ideally using structured tables or plots. |
| Results of syntheses | 20a | For each synthesis, briefly summarise the characteristics and risk of bias among contributing studies. |
|  | 20b | Present results of all statistical syntheses conducted. If meta-analysis was done, present for each the summary estimate and its precision (e.g. confidence/credible interval) and measures of statistical heterogeneity. If comparing groups, describe the direction of the effect. |
|  | 20c | Present results of all investigations of possible causes of heterogeneity among study results. |
|  | 20d | Present results of all sensitivity analyses conducted to assess the robustness of the synthesized results. |
| Reporting biases | 21 | Present assessments of risk of bias due to missing results (arising from reporting biases) for each synthesis assessed. |
| Certainty of evidence | 22 | Present assessments of certainty (or confidence) in the body of evidence for each outcome assessed. |
| **Discussion** |  |  |

| **Section and topic** | **No.** | **Checklist item** |
| --- | --- | --- |
| Discussion | 23a | Provide a general interpretation of the results in the context of other evidence. |
|  | 23b | Discuss any limitations of the evidence included in the review. |
|  | 23c | Discuss any limitations of the review processes used. |
|  | 23d | Discuss implications of the results for practice, policy, and future research. |
| **Other information** |  |  |
| Registration and protocol | 24a | Provide registration information for the review, including register name and registration number, or state that the review was not registered. |
|  | 24b | Indicate where the review protocol can be accessed, or state that a protocol was not prepared. |
|  | 24c | Describe and explain any amendments to information provided at registration or in the protocol. |
| Support | 25 | Describe sources of financial or non-financial support for the review, and the role of the funders or sponsors in the review. |
| Competing interests | 26 | Declare any competing interests of review authors. |
| Availability of data, code and other materials | 27 | Report which of the following are publicly available and where they can be found: template data collection forms; data extracted from included studies; data used for all analyses; analytic code; any other materials used in the review. |

## Appendix 2. Search strategy

**Keywords**

**Appendix 2 Table 1.** Search strategy themes and keywords.

| Theme | Keywords |
| --- | --- |
| Clinical context | Patient, clinician, clinical, clinical visit, clinical encounter |
| Agenda-setting | Agenda setting, agenda mapping, agenda elicitation, agenda solicitation, topic setting, topic mapping, topic elicitation, topic solicitation, concern setting, concern mapping, concern elicitation, concern solicitation, priority elicitation, priority solicitation, patient agenda, opening statement, visit opening, visit structure, encounter opening, encounter structure |
| Interventions | Approach, brochure, checklist, coaching, conversation analysis, conversation approach, dashboard, education, electronic health record, electronic medical record, form, framework, guide, handout, information, instrument, intervention, issue card, material, model, pamphlet, portal, prompt, protocol, question asking, questionnaire, seminar, session, strategy, survey, technique, template, tool, training, webinar, workflow, worksheet, workshop |

**Database searches**

**Database:** APA PsycInfo

**Dates covered:** 1806 to present

**Date last searched:** July 2025

**Limits:** None used.

**Search terms / results:** We searched abstracts in PsycInfo with the queries below. The search returned 428 results.

**Appendix 2 Table 2.**
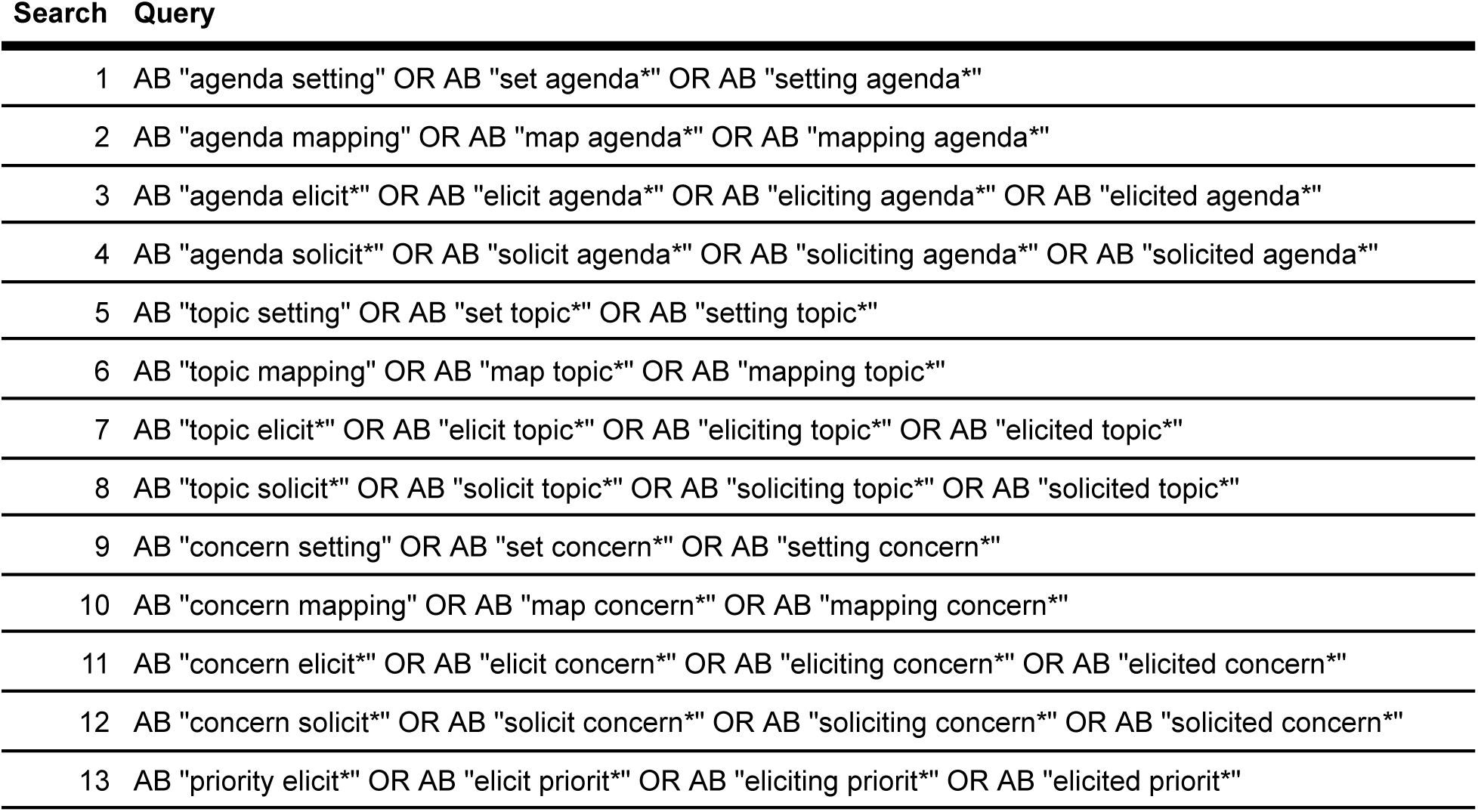
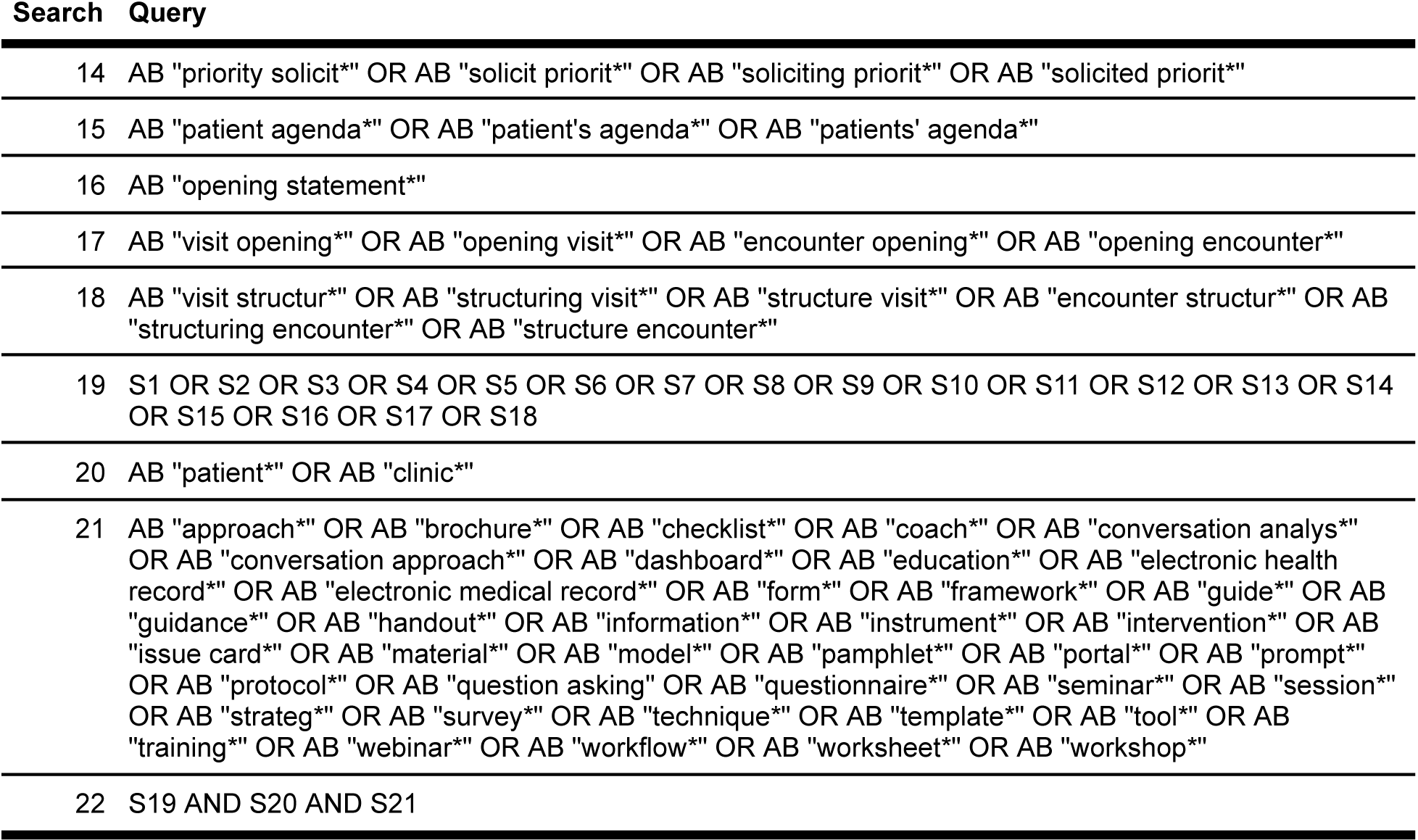
APA PsycInfo search queries.

**Database:** The Cochrane Library, including the Cochrane Database of Systematic Reviews (CDSR) and Cochrane Central Register of Controlled Trials (CENTRAL)

**Dates covered:** CDSR, 1996 to present; CENTRAL, 1989 to present

**Date last searched:** July 2025

**Limits:** None used.

**Search terms / results:** We searched titles, abstracts, and keywords in the Cochrane Library with the queries below. The search returned 234 results.

**Appendix 2 Table 3.**
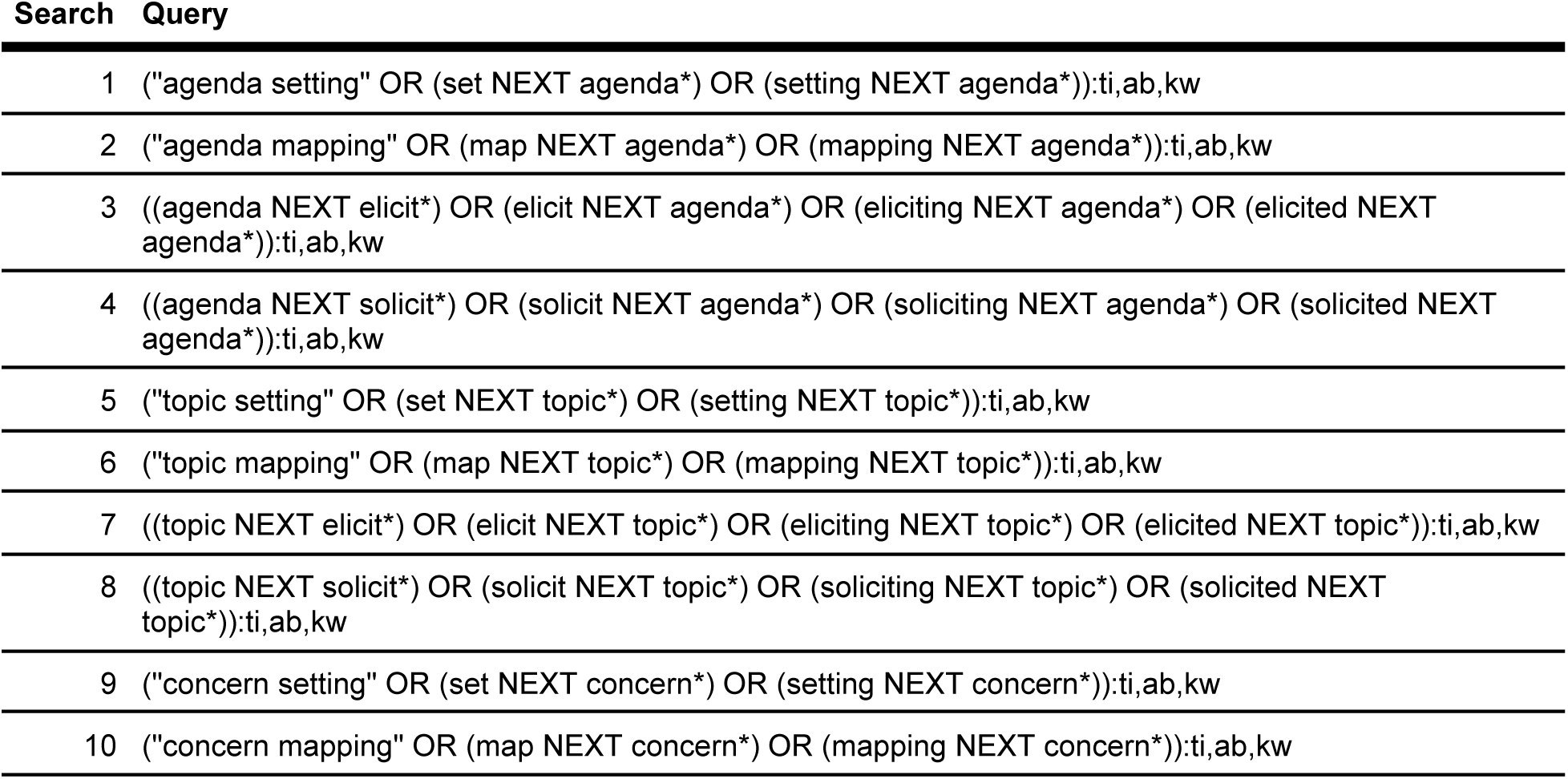
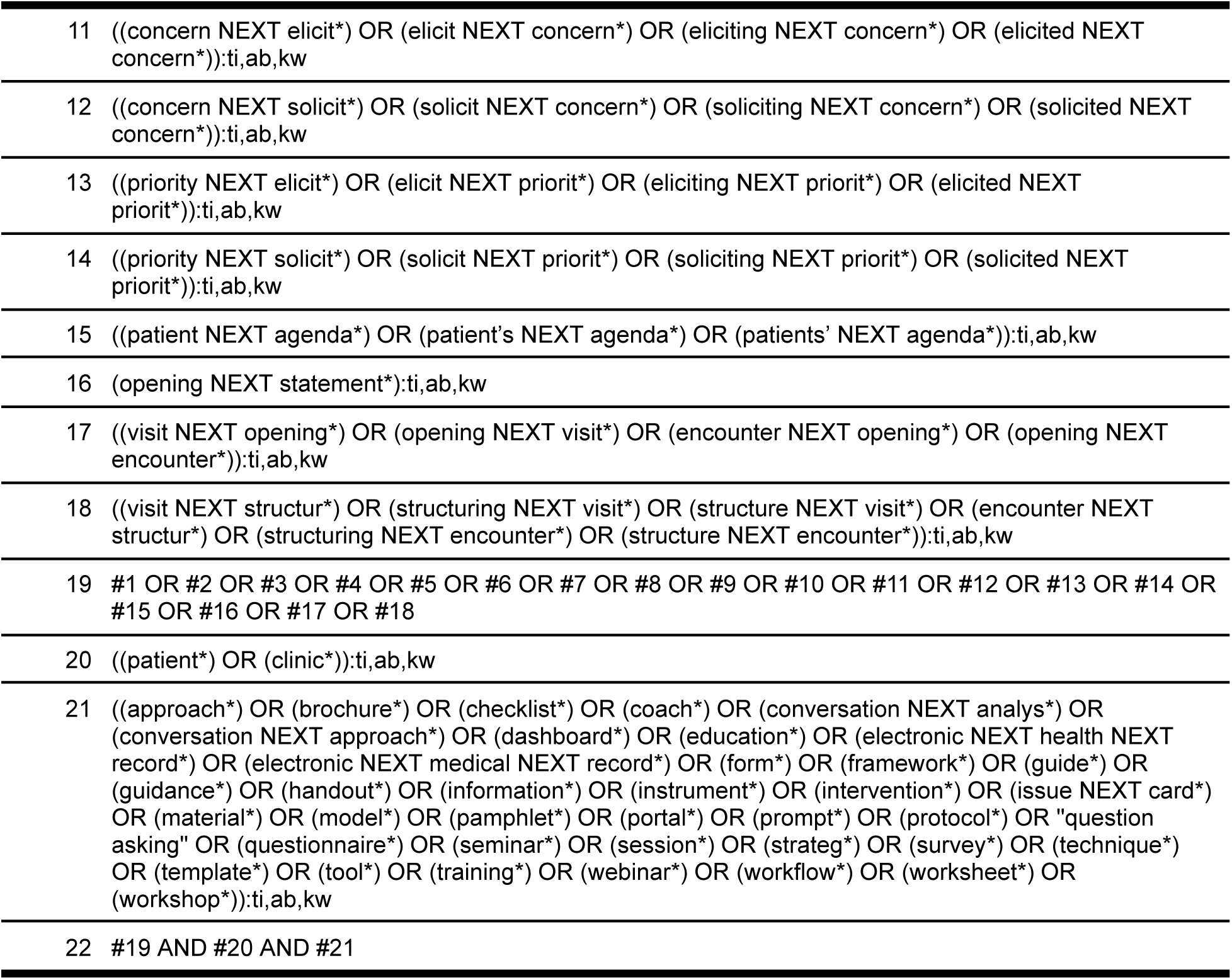
The Cochrane Library search queries.

**Database:** Cumulative Index to Nursing and Allied Health Literature (CINAHL)

**Dates covered:** 1981 to present

**Date last searched:** July 2025

**Limits:** None used.

**Search terms / results:** We searched abstracts in CINAHL with the queries below. We were unable to identify any CINAHL Subject Headings for clinical visit agenda-setting. The search returned 602 results.

**Appendix 2 Table 4.**
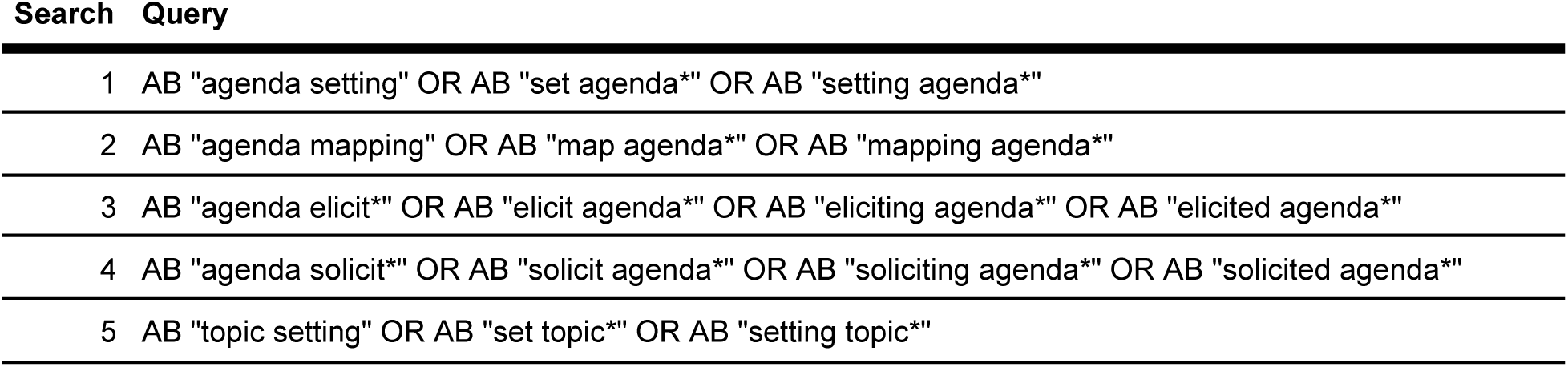
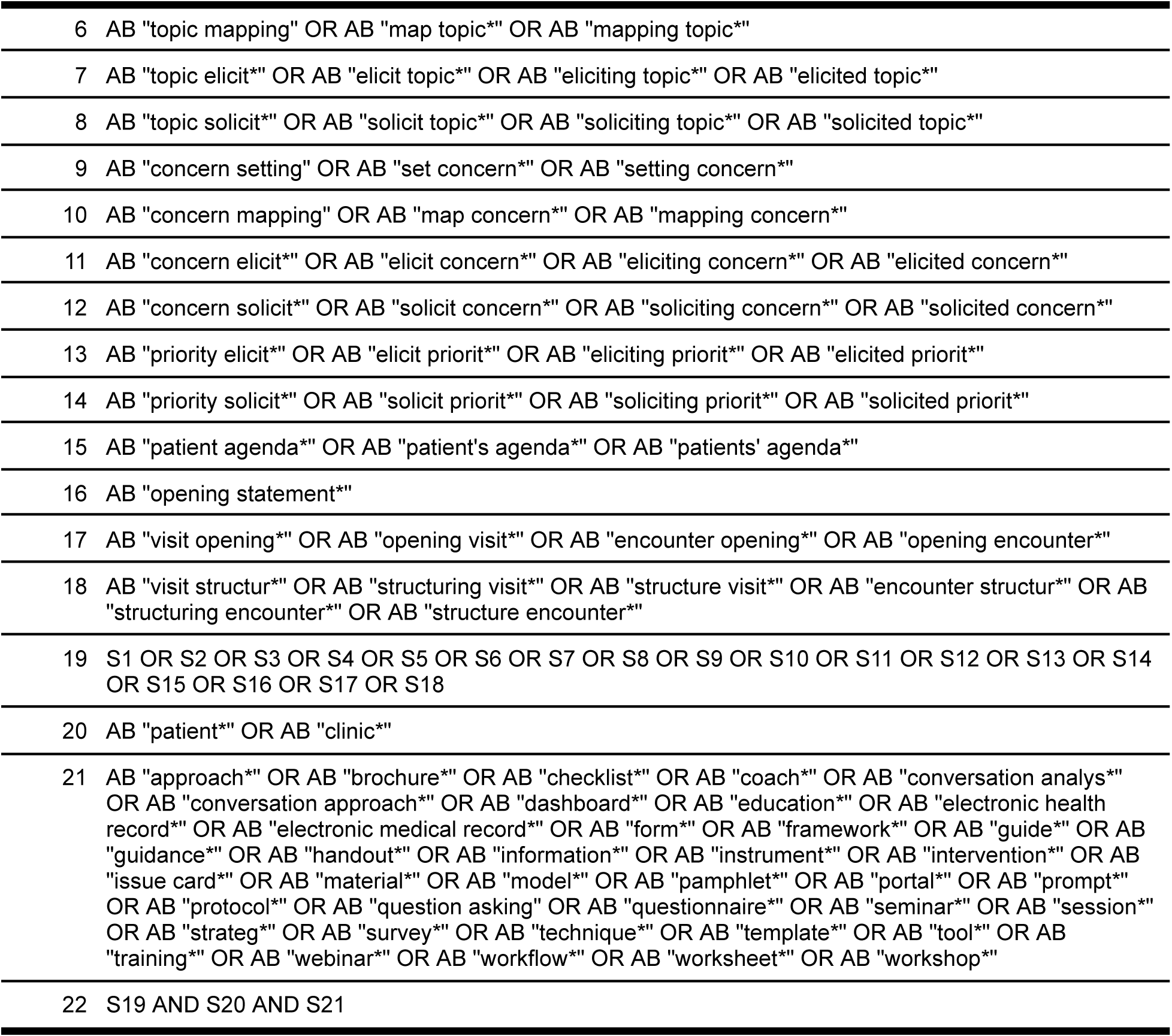
CINAHL search queries.

Search Query
|  |  |
| --- | --- |
| 1 | AB "agenda setting" OR AB "set agenda*" OR AB "setting agenda" |
| 2 | AB "agenda mapping" OR AB "map agenda*" OR AB "mapping agenda" |
| 3 | AB "agenda elicit*" OR AB "elicit agenda*" OR AB "eliciting agenda*" OR AB "elicited agenda" |
| 4 | AB "agenda solicit*" OR AB "solicit agenda*" OR AB "soliciting agenda*" OR AB "solicited agenda" |
| 5 | AB "topic setting" OR AB "set topic*" OR AB "setting topic" |
| Search Query |  |
| 6 | AB "topic mapping" OR AB "map topic*" OR AB "mapping topic" |
| 7 | AB "topic elicit*" OR AB "elicit topic*" OR AB "eliciting topic*" OR AB "elicited topic" |
| 8 | AB "topic solicit*" OR AB "solicit topic*" OR AB "soliciting topic*" OR AB "solicited topic" |
| 9 | AB "concern setting" OR AB "set concern*" OR AB "setting concern" |
| 10 | AB "concern mapping" OR AB "map concern*" OR AB "mapping concern" |
| 11 | AB "concern elicit*" OR AB "elicit concern*" OR AB "eliciting concern*" OR AB "elicited concern" |
| 12 | AB "concern solicit*" OR AB "solicit concern*" OR AB "soliciting concern*" OR AB "solicited concern" |
| 13 | AB "priority elicit*" OR AB "elicit priorit*" OR AB "eliciting priorit*" OR AB "elicited priorit" |
| 14 | AB "priority solicit*" OR AB "solicit priorit*" OR AB "soliciting priorit*" OR AB "solicited priorit" |
| 15 | AB "patient agenda*" OR AB "patient's agenda*" OR AB "patients' agenda" |
| 16 | AB "opening statement" |
| 17 | AB "visit opening*" OR AB "opening visit*" OR AB "encounter opening*" OR AB "opening encounter" |
| 18 | AB "visit structur*" OR AB "structuring visit*" OR AB "structure visit*" OR AB "encounter structur*" OR AB "structuring encounter*" OR AB "structure encounter" |
| 19 | S1 OR S2 OR S3 OR S4 OR S5 OR S6 OR S7 OR S8 OR S9 OR S10 OR S11 OR S12 OR S13 OR S14 OR S15 OR S16 OR S17 OR S18 |
| 20 | AB "patient*" OR AB "clinic" |
| 21 | AB "approach*" OR AB "brochure*" OR AB "checklist*" OR AB "coach*" OR AB "conversation analys*" OR AB "conversation approach*" OR AB "dashboard*" OR AB "education*" OR AB "electronic health record*" OR AB "electronic medical record*" OR AB "form*" OR AB "framework*" OR AB "guide*" OR AB "guidance*" OR AB "handout*" OR AB "information*" OR AB "instrument*" OR AB "intervention*" OR AB "issue card*" OR AB "material*" OR AB "model*" OR AB "pamphlet*" OR AB "portal*" OR AB "prompt*" OR AB "protocol*" OR AB "question asking" OR AB "questionnaire*" OR AB "seminar*" OR AB "session*" OR AB "strateg*" OR AB "survey*" OR AB "technique*" OR AB "template*" OR AB "tool*" OR AB "training*" OR AB "webinar*" OR AB "workflow*" OR AB "worksheet*" OR AB "workshop" |
| 22 | S19 AND S20 AND S21 |

**Database:** MEDLINE via PubMed

**Dates covered:** 1946 to present

**Date last searched:** July 2025

**Limits:** None used.

**Search terms / results:** We searched titles and abstracts in PubMed with the queries below. We were unable to identify any MeSH terms for clinical visit agenda-setting. The search returned 808 results.

**Appendix 2 Table 5.**
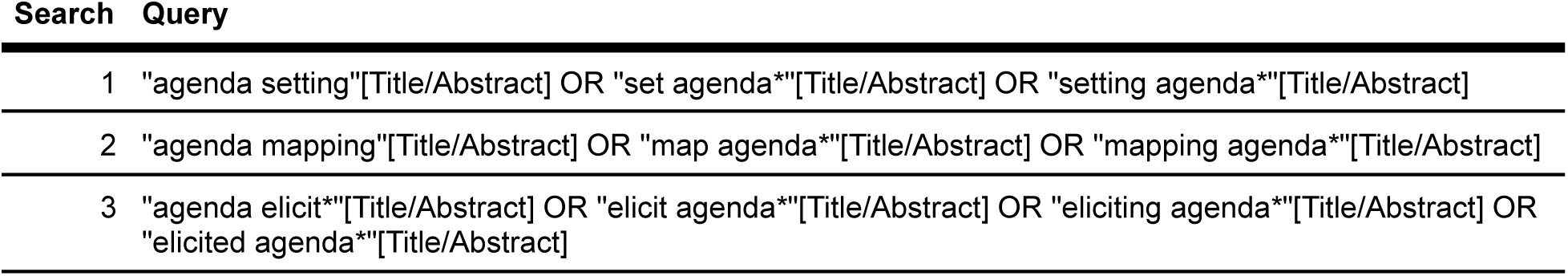
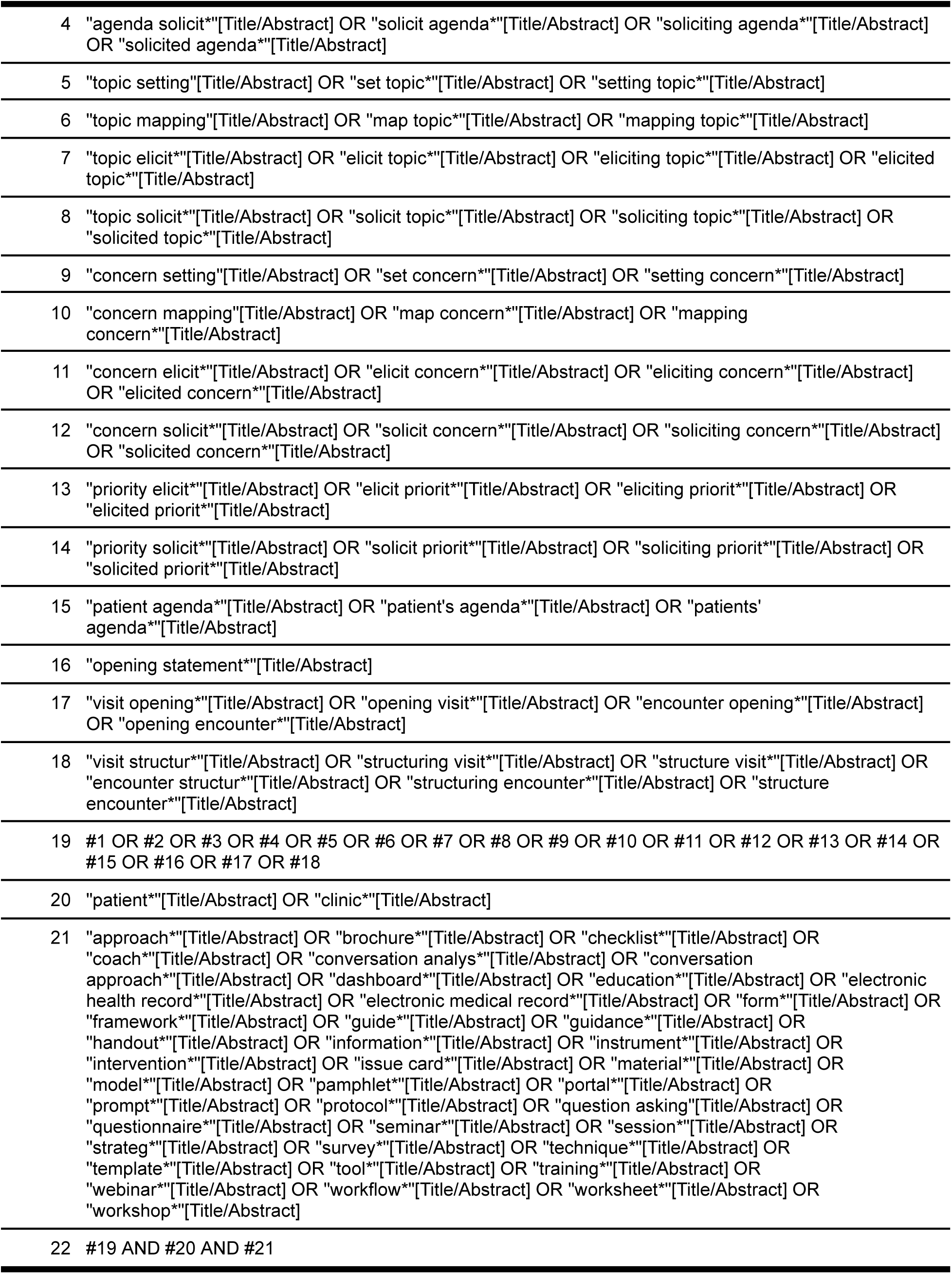
MEDLINE via PubMed search queries.

| Search | Query |
| --- | --- |
| 1 | "agenda setting"[Title/Abstract] OR "set agenda*" [Title/Abstract] OR "setting agenda*" [Title/Abstract] |
| 2 | "agenda mapping"[Title/Abstract] OR "map agenda*" [Title/Abstract] OR "mapping agenda*" [Title/Abstract] |
| 3 | "agenda elicit*" [Title/Abstract] OR "elicit agenda*" [Title/Abstract] OR "eliciting agenda*" [Title/Abstract] OR "elicited agenda*" [Title/Abstract] |
| Search Query |  |
| 4 | "agenda solicit"[Title/Abstract] OR "solicit agenda"[Title/Abstract] OR "soliciting agenda"[Title/Abstract] OR "solicited agenda"[Title/Abstract] |
| 5 | "topic setting"[Title/Abstract] OR "set topic"[Title/Abstract] OR "setting topic"[Title/Abstract] |
| 6 | "topic mapping"[Title/Abstract] OR "map topic"[Title/Abstract] OR "mapping topic"[Title/Abstract] |
| 7 | "topic elicit"[Title/Abstract] OR "elicit topic"[Title/Abstract] OR "eliciting topic"[Title/Abstract] OR "elicited topic"[Title/Abstract] |
| 8 | "topic solicit"[Title/Abstract] OR "solicit topic"[Title/Abstract] OR "soliciting topic"[Title/Abstract] OR "solicited topic"[Title/Abstract] |
| 9 | "concern setting"[Title/Abstract] OR "set concern"[Title/Abstract] OR "setting concern"[Title/Abstract] |
| 10 | "concern mapping"[Title/Abstract] OR "map concern"[Title/Abstract] OR "mapping concern"[Title/Abstract] |
| 11 | "concern elicit"[Title/Abstract] OR "elicit concern"[Title/Abstract] OR "eliciting concern"[Title/Abstract] OR "elicited concern"[Title/Abstract] |
| 12 | "concern solicit"[Title/Abstract] OR "solicit concern"[Title/Abstract] OR "soliciting concern"[Title/Abstract] OR "solicited concern"[Title/Abstract] |
| 13 | "priority elicit"[Title/Abstract] OR "elicit priorit"[Title/Abstract] OR "eliciting priorit"[Title/Abstract] OR "elicited priorit"[Title/Abstract] |
| 14 | "priority solicit"[Title/Abstract] OR "solicit priorit"[Title/Abstract] OR "soliciting priorit"[Title/Abstract] OR "solicited priorit"[Title/Abstract] |
| 15 | "patient agenda"[Title/Abstract] OR "patient's agenda"[Title/Abstract] OR "patients' agenda"[Title/Abstract] |
| 16 | "opening statement"[Title/Abstract] |
| 17 | "visit opening"[Title/Abstract] OR "opening visit"[Title/Abstract] OR "encounter opening"[Title/Abstract] OR "opening encounter"[Title/Abstract] |
| 18 | "visit structur"[Title/Abstract] OR "structuring visit"[Title/Abstract] OR "structure visit"[Title/Abstract] OR "encounter structur"[Title/Abstract] OR "structuring encounter"[Title/Abstract] OR "structure encounter"[Title/Abstract] |
| 19 | #1 OR #2 OR #3 OR #4 OR #5 OR #6 OR #7 OR #8 OR #9 OR #10 OR #11 OR #12 OR #13 OR #14 OR #15 OR #16 OR #17 OR #18 |
| 20 | "patient"[Title/Abstract] OR "clinic"[Title/Abstract] |
| 21 | "approach"[Title/Abstract] OR "brochure"[Title/Abstract] OR "checklist"[Title/Abstract] OR "coach"[Title/Abstract] OR "conversation analys"[Title/Abstract] OR "conversation approach"[Title/Abstract] OR "dashboard"[Title/Abstract] OR "education"[Title/Abstract] OR "electronic health record"[Title/Abstract] OR "electronic medical record"[Title/Abstract] OR "form"[Title/Abstract] OR "framework"[Title/Abstract] OR "guide"[Title/Abstract] OR "guidance"[Title/Abstract] OR "handout"[Title/Abstract] OR "information"[Title/Abstract] OR "instrument"[Title/Abstract] OR "intervention"[Title/Abstract] OR "issue card"[Title/Abstract] OR "material"[Title/Abstract] OR "model"[Title/Abstract] OR "pamphlet"[Title/Abstract] OR "portal"[Title/Abstract] OR "prompt"[Title/Abstract] OR "protocol"[Title/Abstract] OR "question asking"[Title/Abstract] OR "questionnaire"[Title/Abstract] OR "seminar"[Title/Abstract] OR "session"[Title/Abstract] OR "strateg"[Title/Abstract] OR "survey"[Title/Abstract] OR "technique"[Title/Abstract] OR "template"[Title/Abstract] OR "tool"[Title/Abstract] OR "training"[Title/Abstract] OR "webinar"[Title/Abstract] OR "workflow"[Title/Abstract] OR "worksheet"[Title/Abstract] OR "workshop"[Title/Abstract] |
| 22 | #19 AND #20 AND #21 |

**Database:** ProQuest

**Dates covered:** 1861 to present

**Date last searched:** July 2025

**Limits:** None used.

**Search terms / results:** We searched abstracts and summaries in ProQuest with the queries below. The search returned 826 results.

**Appendix 2 Table 6.**
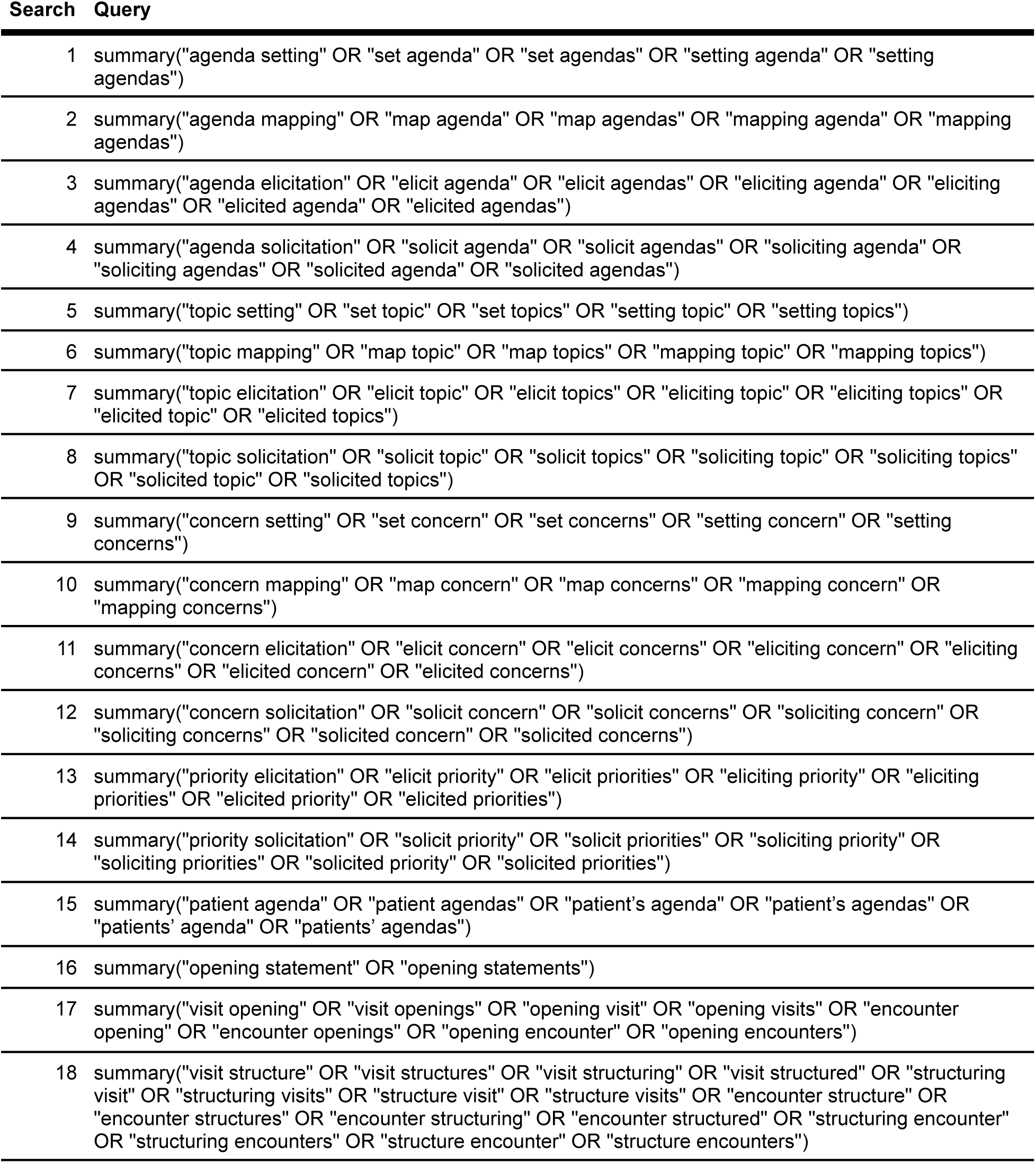
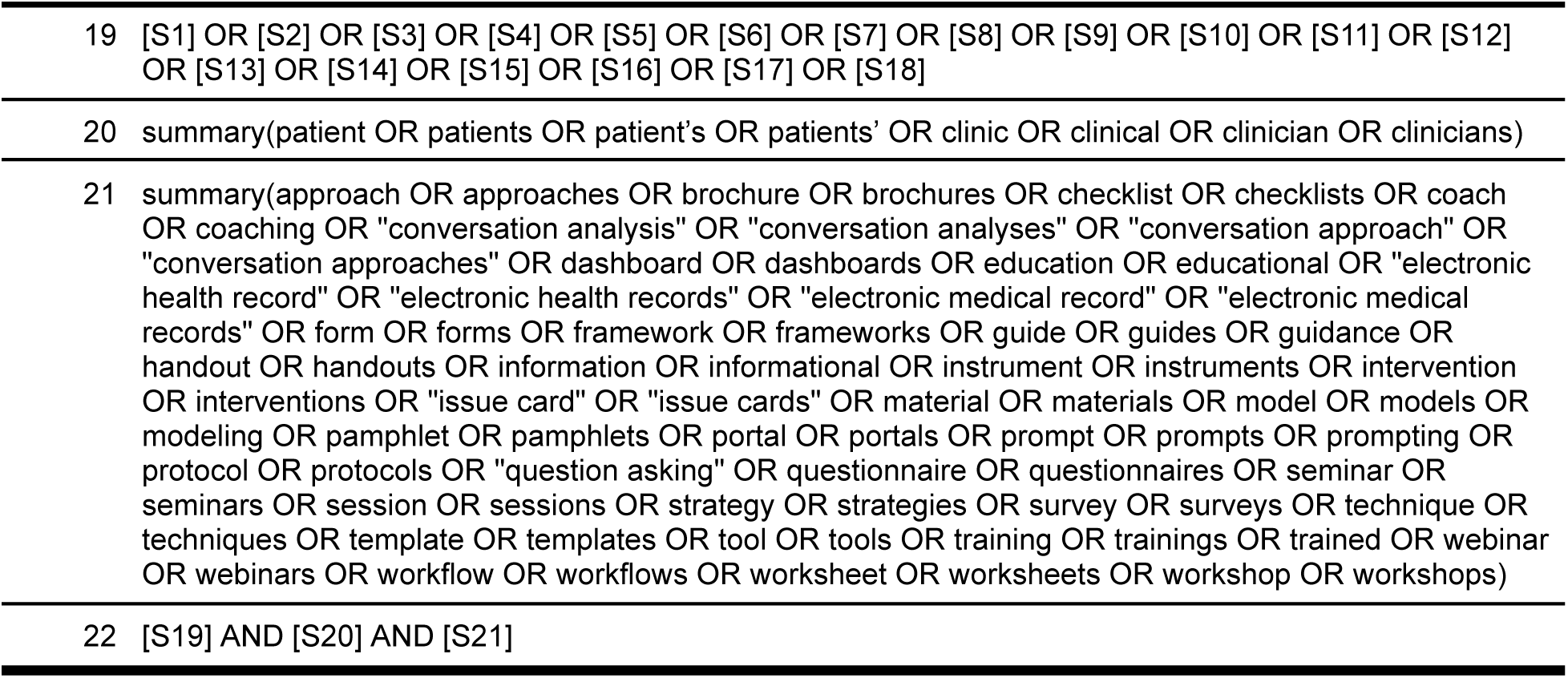
ProQuest search queries.

| Search | Query |
| --- | --- |
| 1 | summary("agenda setting" OR "set agenda" OR "set agendas" OR "setting agenda" OR "setting agendas") |
| 2 | summary("agenda mapping" OR "map agenda" OR "map agendas" OR "mapping agenda" OR "mapping agendas") |
| 3 | summary("agenda elicitation" OR "elicit agenda" OR "elicit agendas" OR "eliciting agenda" OR "eliciting agendas" OR "elicited agenda" OR "elicited agendas") |
| 4 | summary("agenda solicitation" OR "solicit agenda" OR "solicit agendas" OR "soliciting agenda" OR "soliciting agendas" OR "solicited agenda" OR "solicited agendas") |
| 5 | summary("topic setting" OR "set topic" OR "set topics" OR "setting topic" OR "setting topics") |
| 6 | summary("topic mapping" OR "map topic" OR "map topics" OR "mapping topic" OR "mapping topics") |
| 7 | summary("topic elicitation" OR "elicit topic" OR "elicit topics" OR "eliciting topic" OR "eliciting topics" OR "elicited topic" OR "elicited topics") |
| 8 | summary("topic solicitation" OR "solicit topic" OR "solicit topics" OR "soliciting topic" OR "soliciting topics" OR "solicited topic" OR "solicited topics") |
| 9 | summary("concern setting" OR "set concern" OR "set concerns" OR "setting concern" OR "setting concerns") |
| 10 | summary("concern mapping" OR "map concern" OR "map concerns" OR "mapping concern" OR "mapping concerns") |
| 11 | summary("concern elicitation" OR "elicit concern" OR "elicit concerns" OR "eliciting concern" OR "eliciting concerns" OR "elicited concern" OR "elicited concerns") |
| 12 | summary("concern solicitation" OR "solicit concern" OR "solicit concerns" OR "soliciting concern" OR "soliciting concerns" OR "solicited concern" OR "solicited concerns") |
| 13 | summary("priority elicitation" OR "elicit priority" OR "elicit priorities" OR "eliciting priority" OR "eliciting priorities" OR "elicited priority" OR "elicited priorities") |
| 14 | summary("priority solicitation" OR "solicit priority" OR "solicit priorities" OR "soliciting priority" OR "soliciting priorities" OR "solicited priority" OR "solicited priorities") |
| 15 | summary("patient agenda" OR "patient agendas" OR "patient's agenda" OR "patient's agendas" OR "patients' agenda" OR "patients' agendas") |
| 16 | summary("opening statement" OR "opening statements") |
| 17 | summary("visit opening" OR "visit openings" OR "opening visit" OR "opening visits" OR "encounter opening" OR "encounter openings" OR "opening encounter" OR "opening encounters") |
| 18 | summary("visit structure" OR "visit structures" OR "visit structuring" OR "visit structured" OR "structuring visit" OR "structuring visits" OR "structure visit" OR "structure visits" OR "encounter structure" OR "encounter structures" OR "encounter structuring" OR "encounter structured" OR "structuring encounter" OR "structuring encounters" OR "structure encounter" OR "structure encounters") |
| Search Query |  |
| 19 | [S1] OR [S2] OR [S3] OR [S4] OR [S5] OR [S6] OR [S7] OR [S8] OR [S9] OR [S10] OR [S11] OR [S12] OR [S13] OR [S14] OR [S15] OR [S16] OR [S17] OR [S18] |
| 20 | summary(patient OR patients OR patient's OR patients' OR clinic OR clinical OR clinician OR clinicians) |
| 21 | summary(approach OR approaches OR brochure OR brochures OR checklist OR checklists OR coach OR coaching OR "conversation analysis" OR "conversation analyses" OR "conversation approach" OR "conversation approaches" OR dashboard OR dashboards OR education OR educational OR "electronic health record" OR "electronic health records" OR "electronic medical record" OR "electronic medical records" OR form OR forms OR framework OR frameworks OR guide OR guides OR guidance OR handout OR handouts OR information OR informational OR instrument OR instruments OR intervention OR interventions OR "issue card" OR "issue cards" OR material OR materials OR model OR models OR modeling OR pamphlet OR pamphlets OR portal OR portals OR prompt OR prompts OR prompting OR protocol OR protocols OR "question asking" OR questionnaire OR questionnaires OR seminar OR seminars OR session OR sessions OR strategy OR strategies OR survey OR surveys OR technique OR techniques OR template OR templates OR tool OR tools OR training OR trainings OR trained OR webinar OR webinars OR workflow OR workflows OR worksheet OR worksheets OR workshop OR workshops) |
| 22 | [S19] AND [S20] AND [S21] |

**Database:** Scopus

**Dates covered:** 1788 to present

**Date last searched:** July 2025

**Limits:** None used.

**Search terms / results:** We searched titles, abstracts, and keywords in Scopus with the queries below. The search returned 1,033 results.

**Appendix 2 Table 7.**
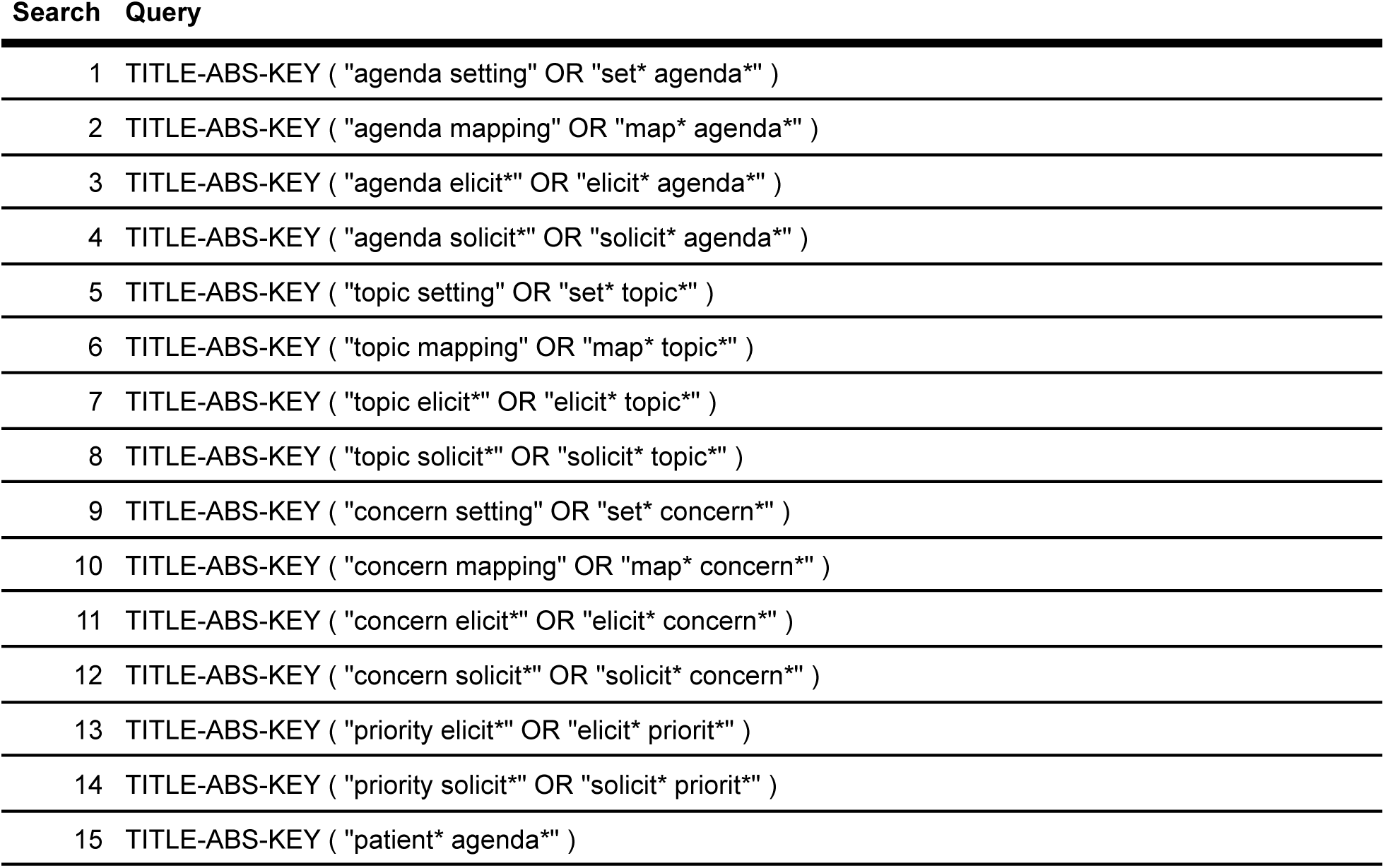
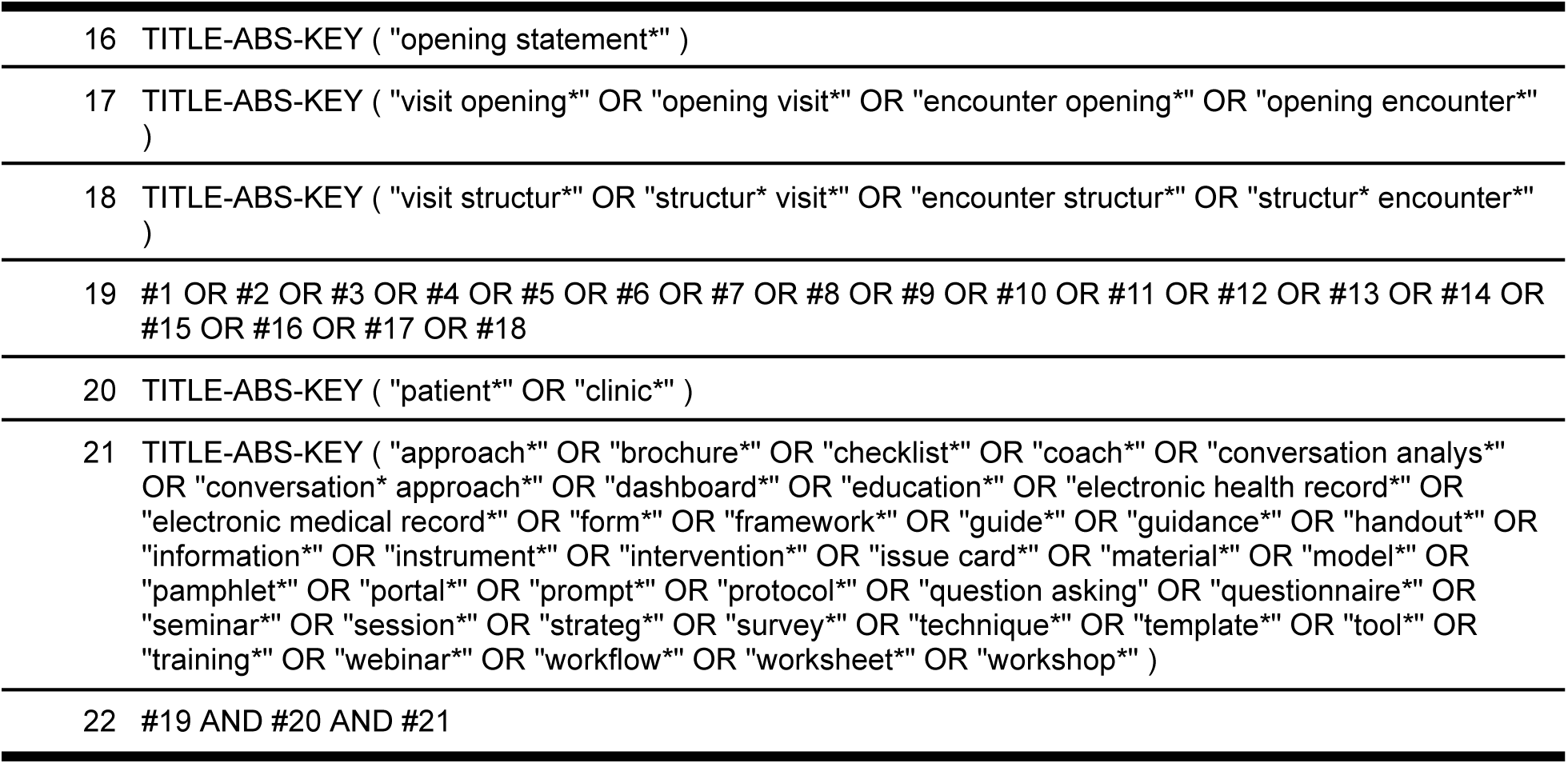
Scopus search queries.

**Database:** Web of Science

**Dates covered:** 1900 to present

**Date last searched:** July 2025

**Limits:** None used.

**Search terms / results:** We searched abstracts in Web of Science with the queries below. The search returned 1,233 results.

**Appendix 2 Table 8.**
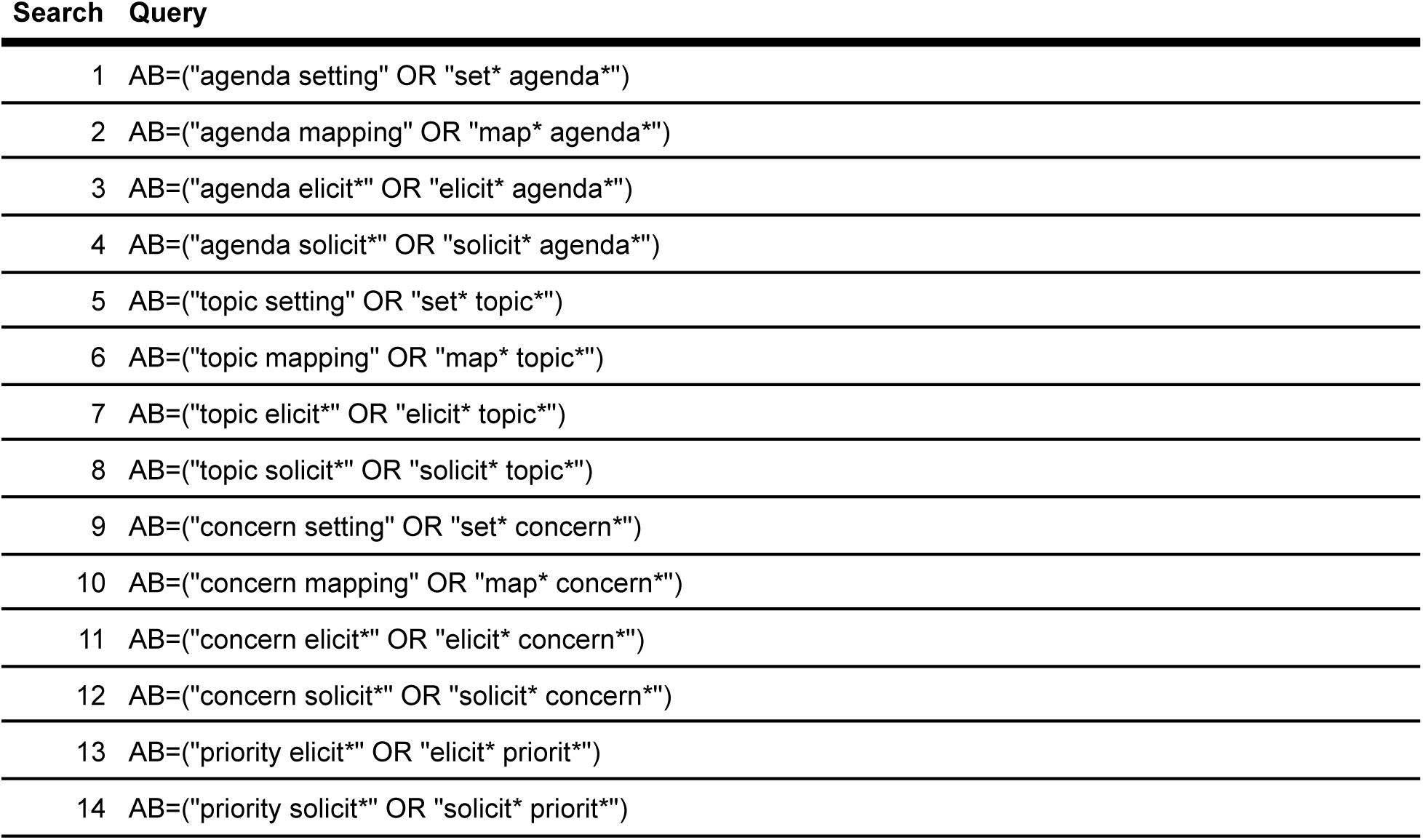
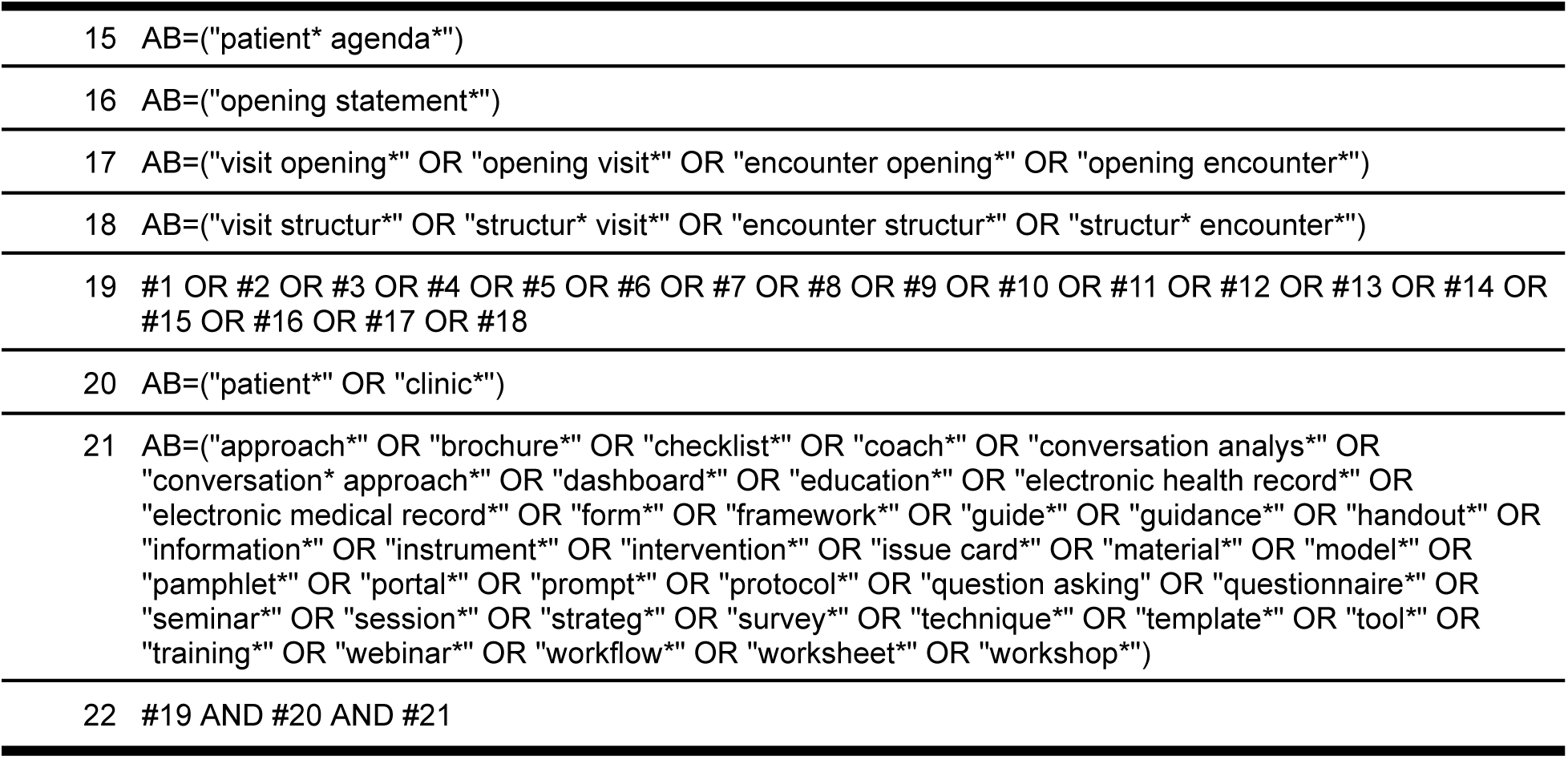
Web of Science search queries.

**Trial registry searches**

**Database:** ClinicalTrials.gov

**Dates covered:** 1997 to present

**Date last searched:** July 2025

**Limits:** None used.

**Search terms / results:** We searched ClinicalTrials.gov using the search string below entered in the ‘Other terms’ field. The search returned 20 results.

*“agenda setting” OR “agenda mapping” OR “patient agenda” OR “visit opening” OR “topic elicitation” OR “topic solicitation“*

**Other searches**

**Source:** Google Scholar

**Dates covered:** Unknown

**Date last searched:** July 2025

**Limits:** None used.

**Search terms / results:** We searched Google Scholar using the search string below, without using any modifiers. The search returned about 18,500 results. Of these, we only viewed 250 results, in accordance with our protocol.

*(“agenda setting” OR “agenda mapping” OR “patient agenda” OR “visit opening” OR “topic elicitation” OR “topic solicitation”) AND clinic* AND intervention\**

## Appendix 3. Articles excluded in full-text screening

**Appendix 3 Table 1.** Articles excluded in full-text screening with reasons for exclusion.

| Article | Reason for exclusion |
| --- | --- |
| Burggraf et al. (2019)<sup>91</sup> | No eligible comparative intervention results |
| Hamilton and Britten (2006)<sup>92</sup> | Not an original comparative empirical study |
| Jones et al. (1999)<sup>93</sup> | Not an eligible agenda-setting intervention |
| Lang and McCord (1999)<sup>94</sup> | Not an original comparative empirical study |
| Matthias et al. (2022)<sup>95</sup> | No eligible comparative intervention results |
| Morrissey et al. (2022)<sup>96</sup> | Not an eligible agenda-setting intervention |
| Rosa et al. (2022)<sup>97</sup> | Not an eligible agenda-setting intervention |
| Silber and Rosenthal (1986)<sup>98</sup> | No eligible comparator |
| Stott et al. (1995)<sup>99</sup> | Not an eligible agenda-setting intervention |
| Stott et al. (1996)<sup>100</sup> | Not an eligible agenda-setting intervention |
| Vegni et al. (2002)<sup>101</sup> | Not an eligible agenda-setting intervention |
| Wissow et al. (2011)<sup>102</sup> | Not an eligible agenda-setting intervention |
| Wolff et al. (2021)<sup>103</sup> | No eligible comparator |

## Appendix 4. Effects of agenda-setting interventions on additional outcomes

**Appendix 4 Figure 1.**
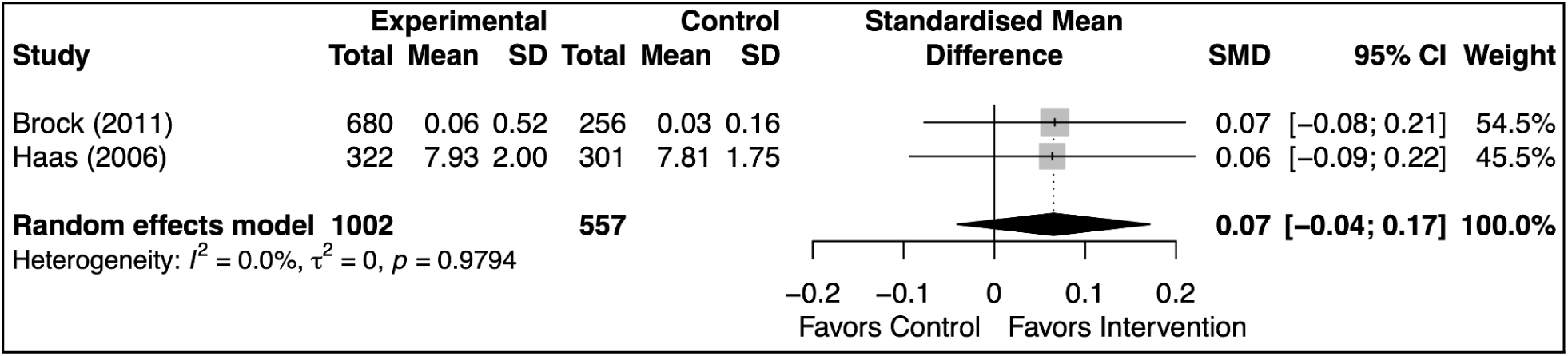
Effects of agenda-setting interventions on initial elicitation of concerns

**Appendix 4 Figure 2.**
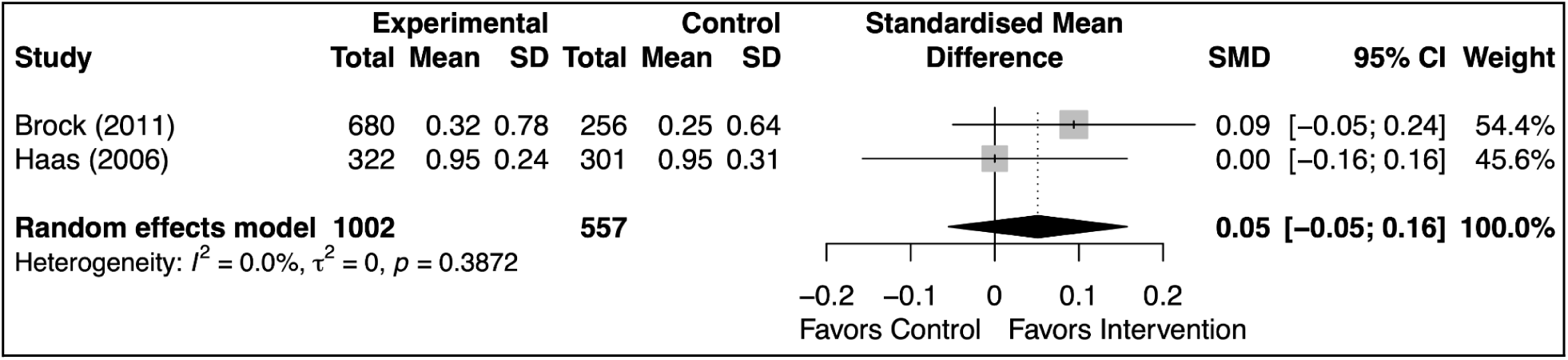
Effects of agenda-setting interventions on elicitation of additional concerns

**Appendix 4 Figure 3.**
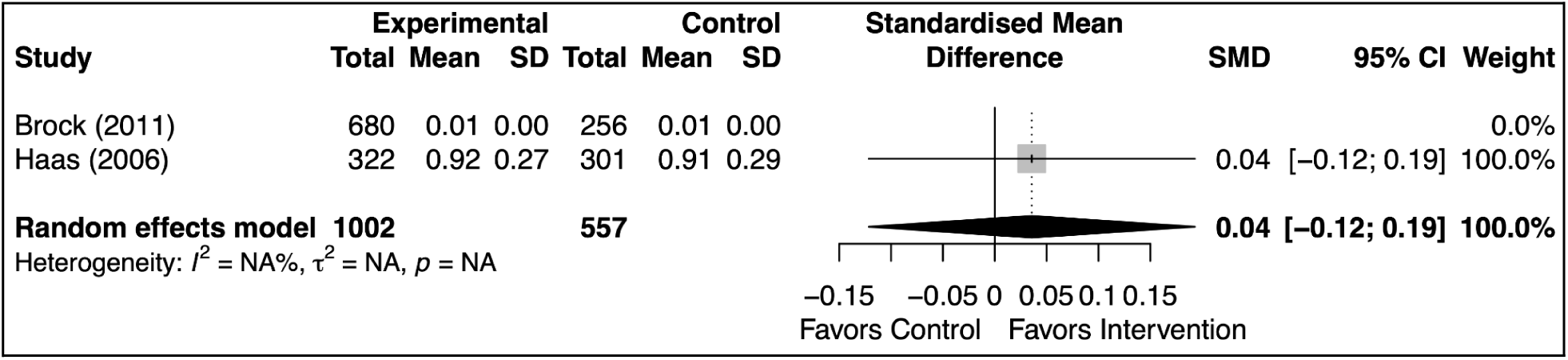
Effects of agenda-setting interventions on prioritization of concerns

**Appendix 4 Figure 4.**
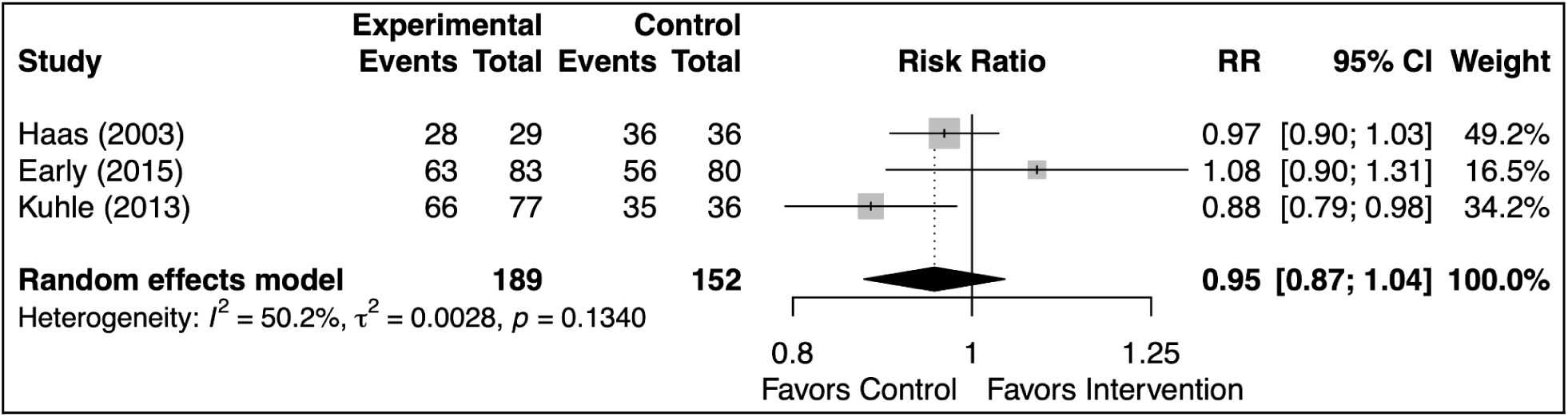
Effects of agenda-setting interventions on concerns addressed, dichotomous

**Appendix 4 Figure 5.**
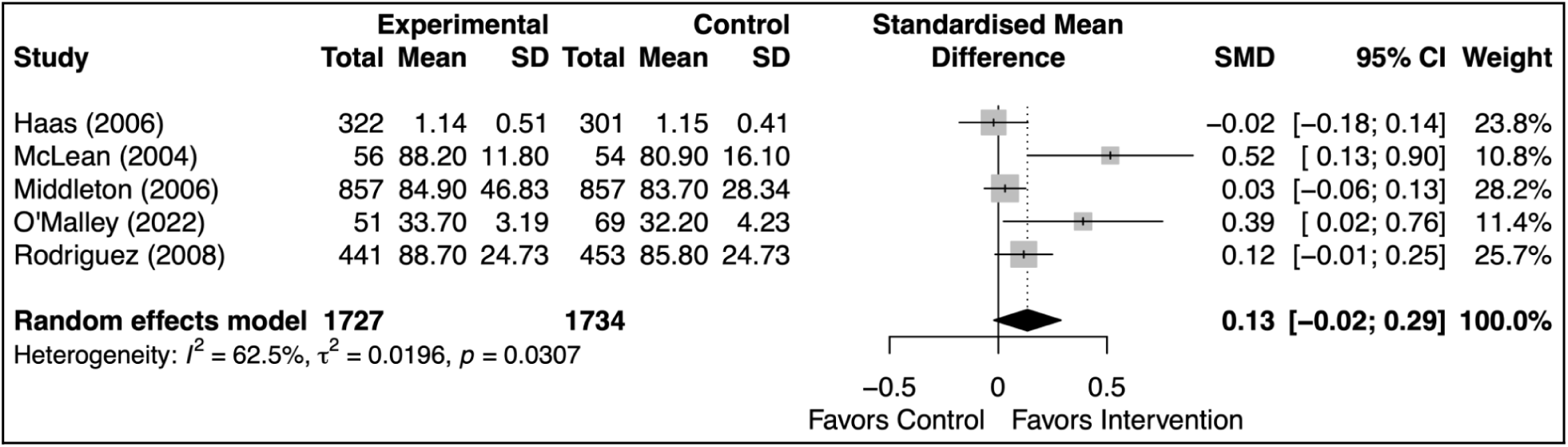
Effects of agenda-setting interventions on satisfaction with clinician

**Appendix 4 Figure 6.**
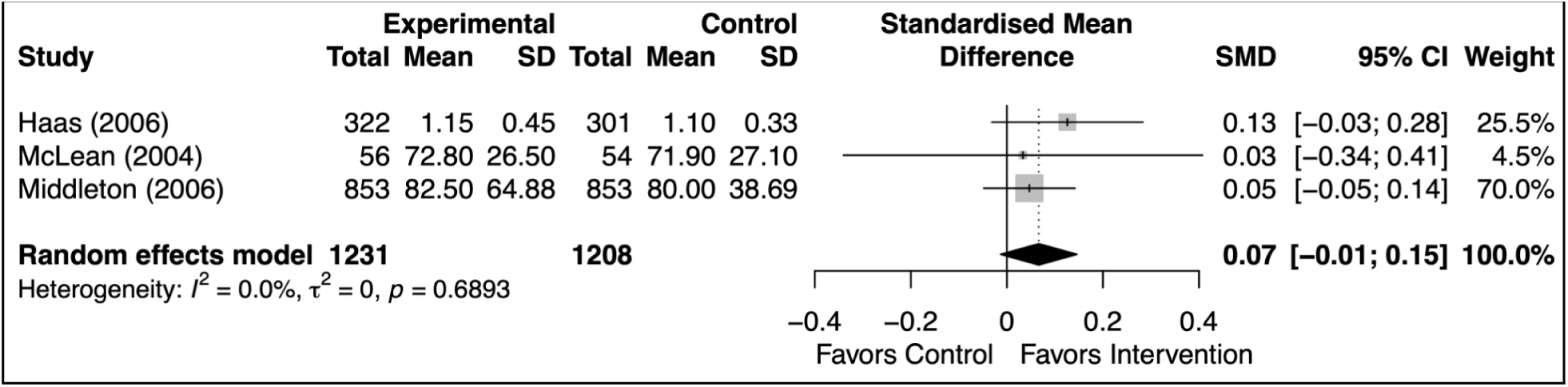
Effects of agenda-setting interventions on adequacy of visit time

**Appendix 4 Figure 7.**
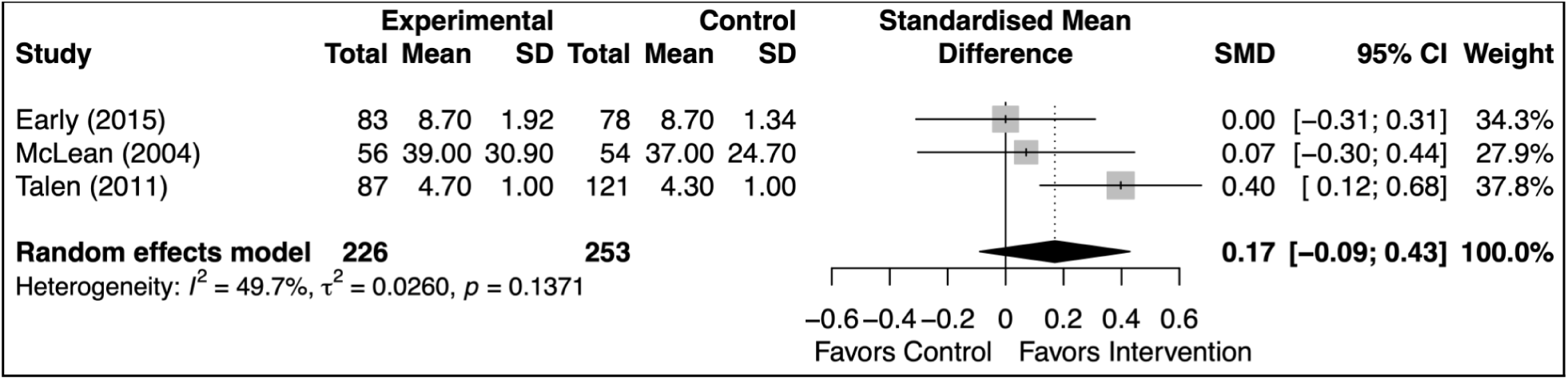
Effects of agenda-setting interventions on patient activation or enablement

## Appendix 5. Post hoc exploratory subgroup analyses

### Study design

**Appendix 5 Figure 1.**
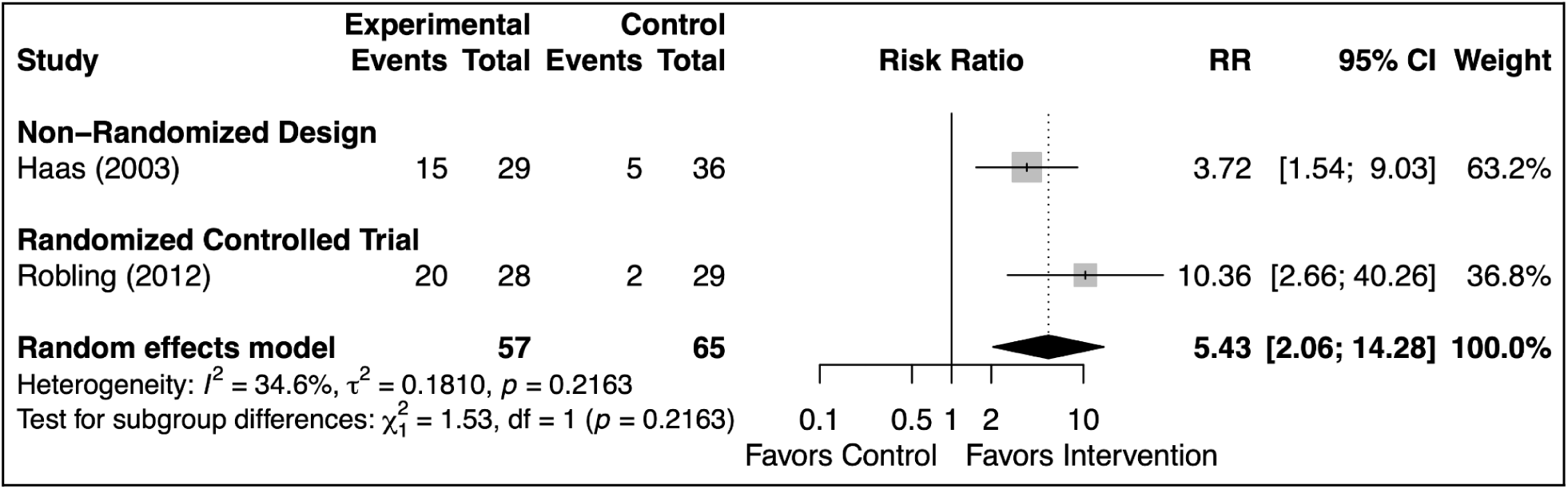
Effects of agenda-setting interventions on occurrence of agenda-setting, stratified by study design

**Appendix 5 Figure 2.**
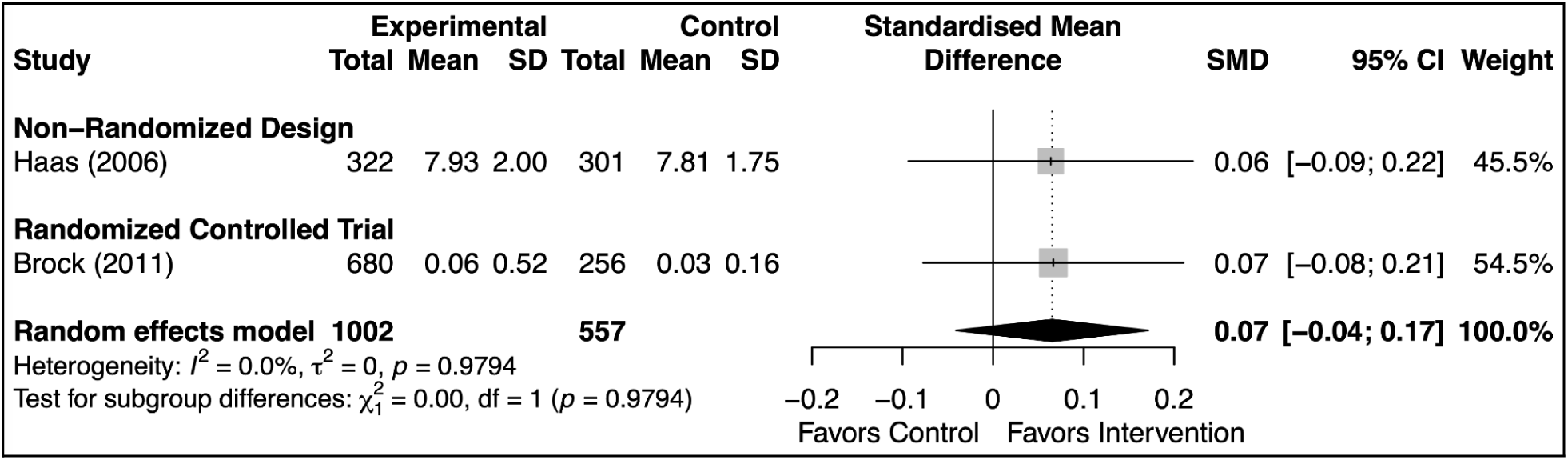
Effects of agenda-setting interventions on initial elicitation of concerns, stratified by study design

**Appendix 5 Figure 3.**
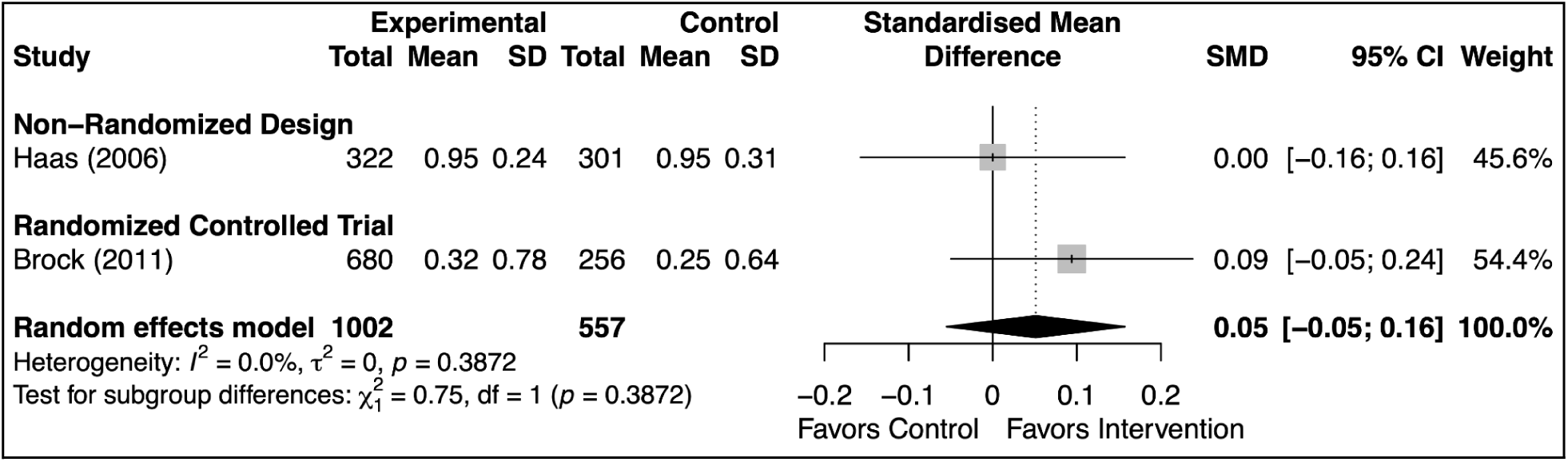
Effects of agenda-setting interventions on elicitation of additional concerns, stratified by study design

**Appendix 5 Figure 4.**
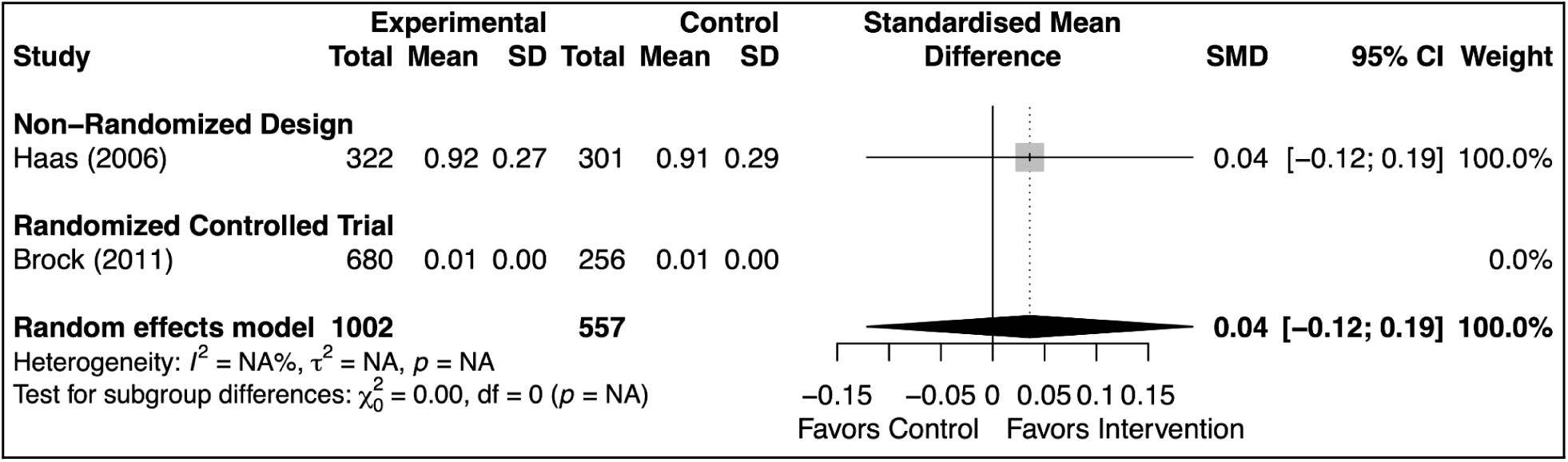
Effects of agenda-setting interventions on prioritization of concerns, stratified by study design

**Appendix 5 Figure 5.**
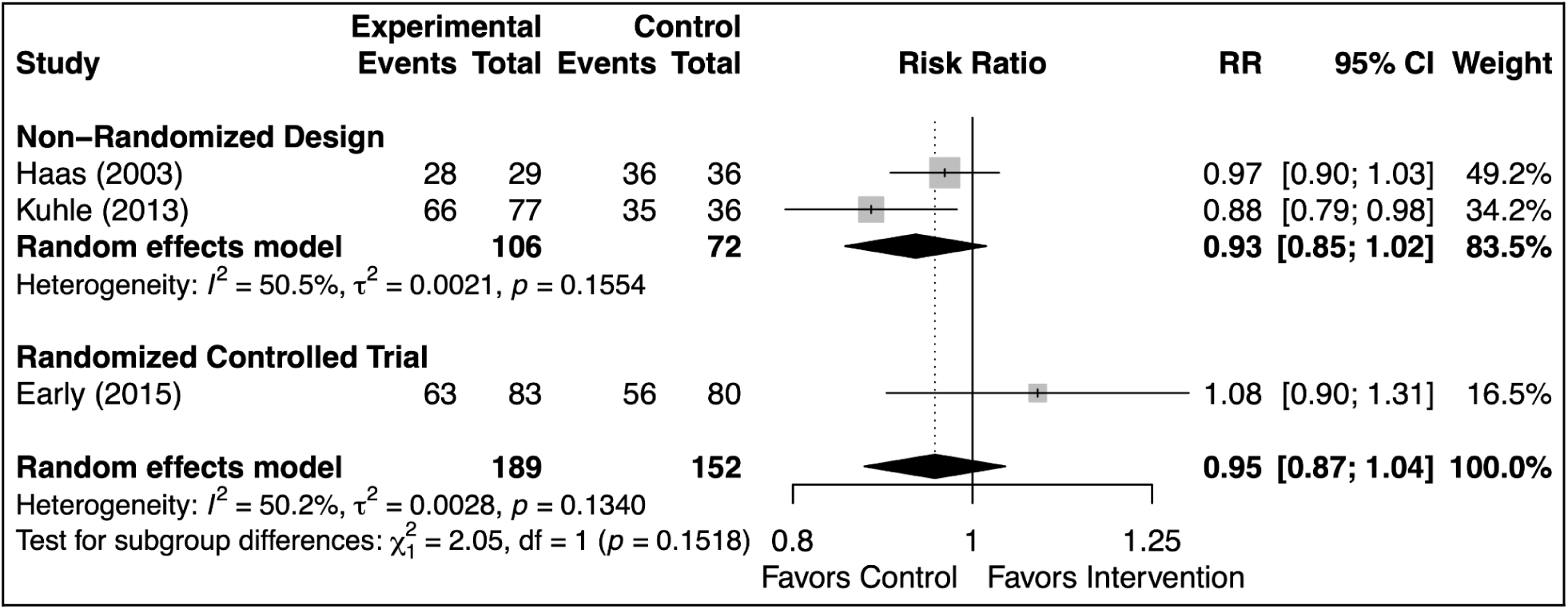
Effects of agenda-setting interventions on concerns addressed, dichotomous, stratified by study design

**Appendix 5 Figure 6.**
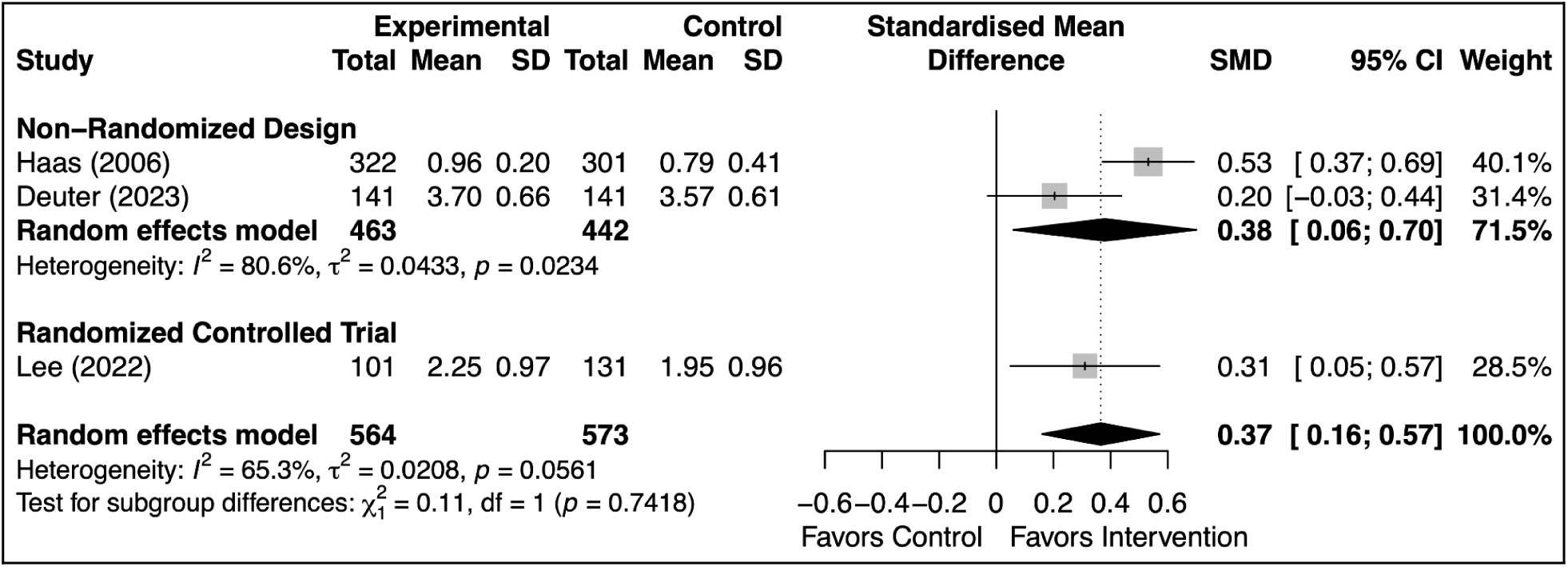
Effects of agenda-setting interventions on concerns addressed, continuous, stratified by study design

**Appendix 5 Figure 7.**
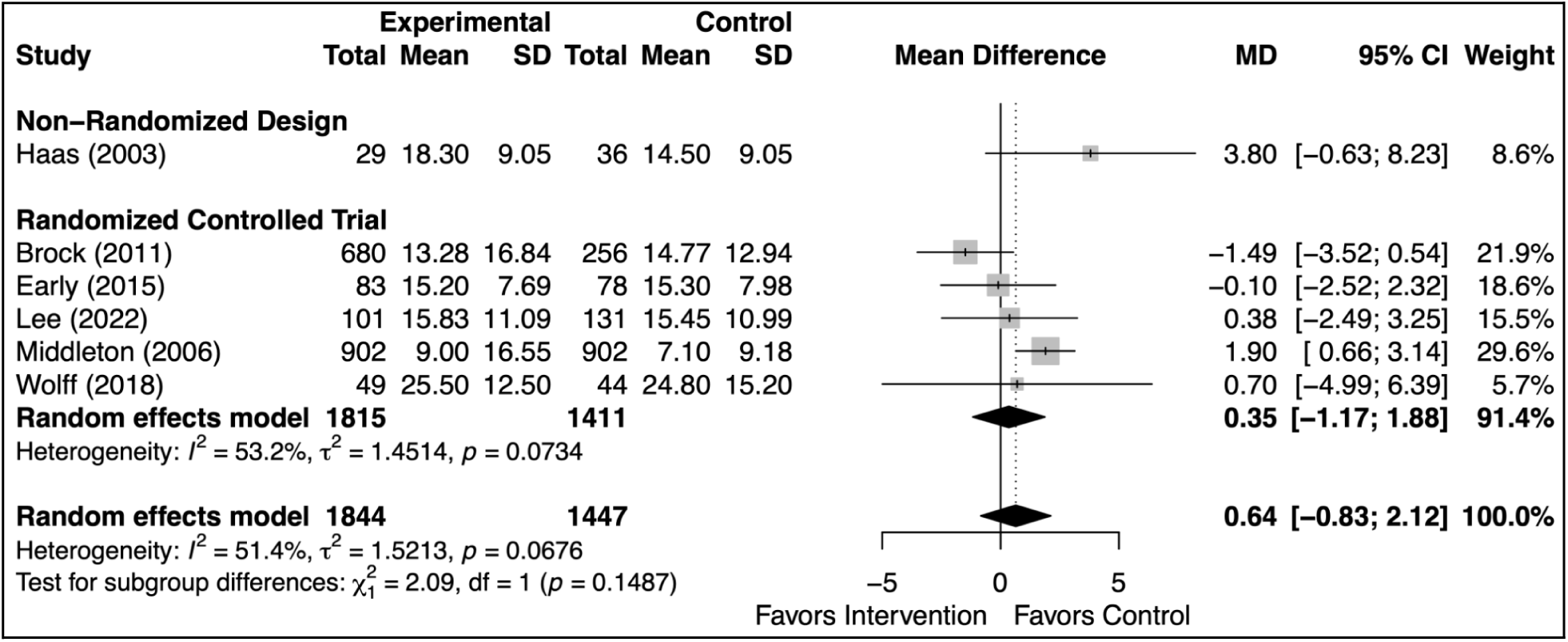
Effects of agenda-setting interventions on visit duration, stratified by study design

**Appendix 5 Figure 8.**
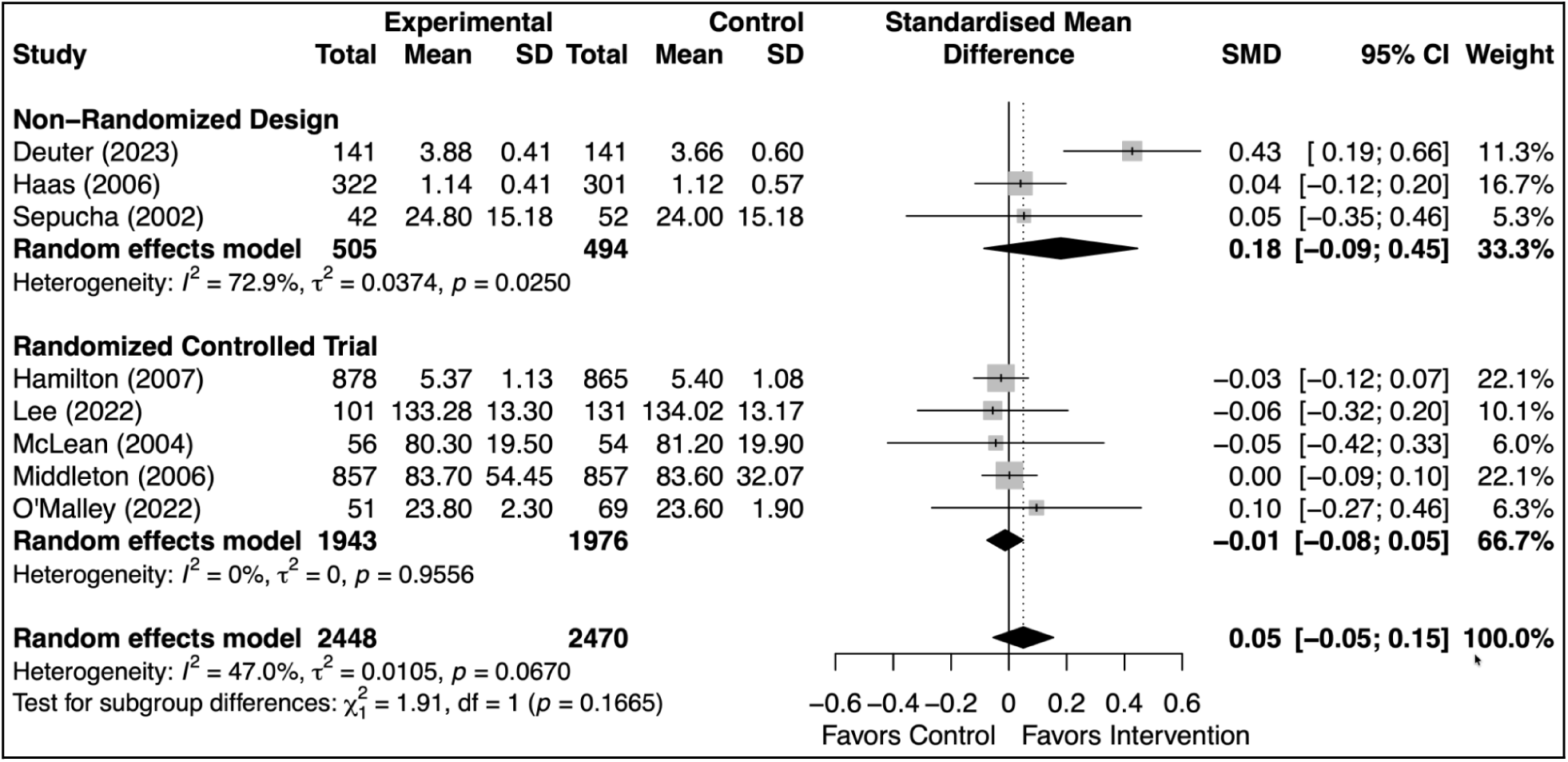
Effects of agenda-setting interventions on overall patient satisfaction, stratified by study design

**Appendix 5 Figure 9.**
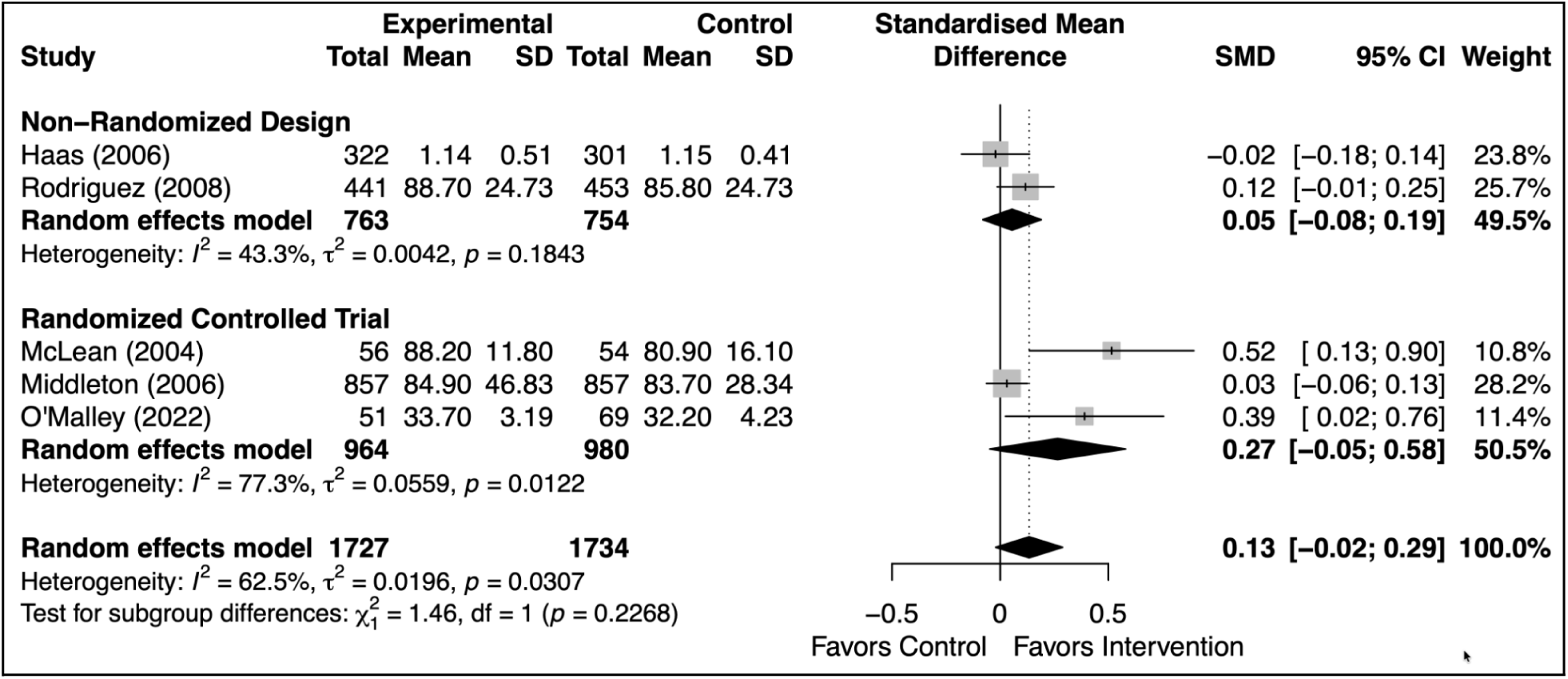
Effects of agenda-setting interventions on satisfaction with clinician, stratified by study design

**Appendix 5 Figure 10.**
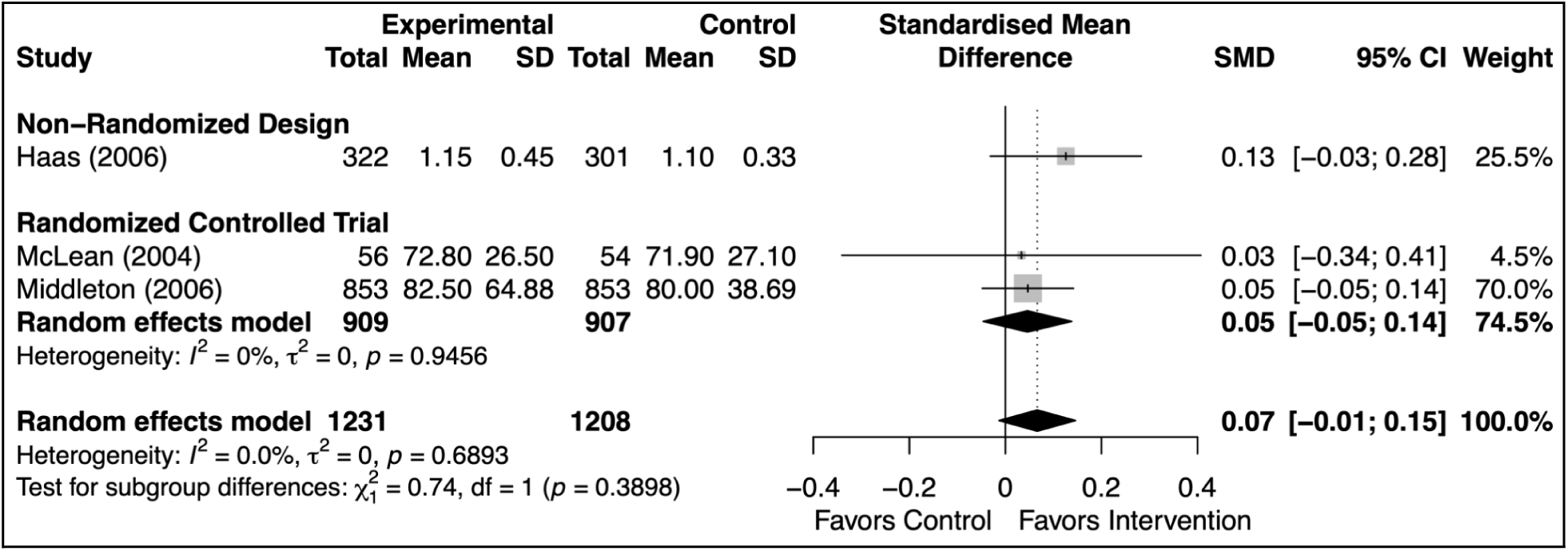
Effects of agenda-setting interventions on adequacy of visit time, stratified by study design

**Appendix 5 Figure 11.**
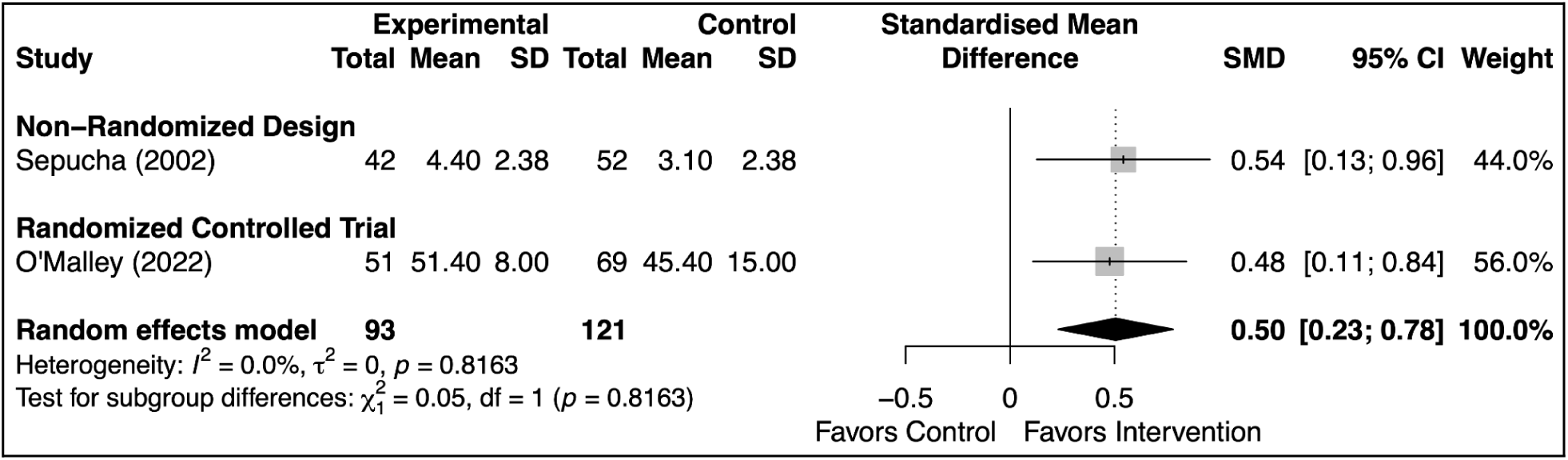
Effects of agenda-setting interventions on overall clinician satisfaction, stratified by study design

### Adjustment status

**Appendix 5 Figure 12.**
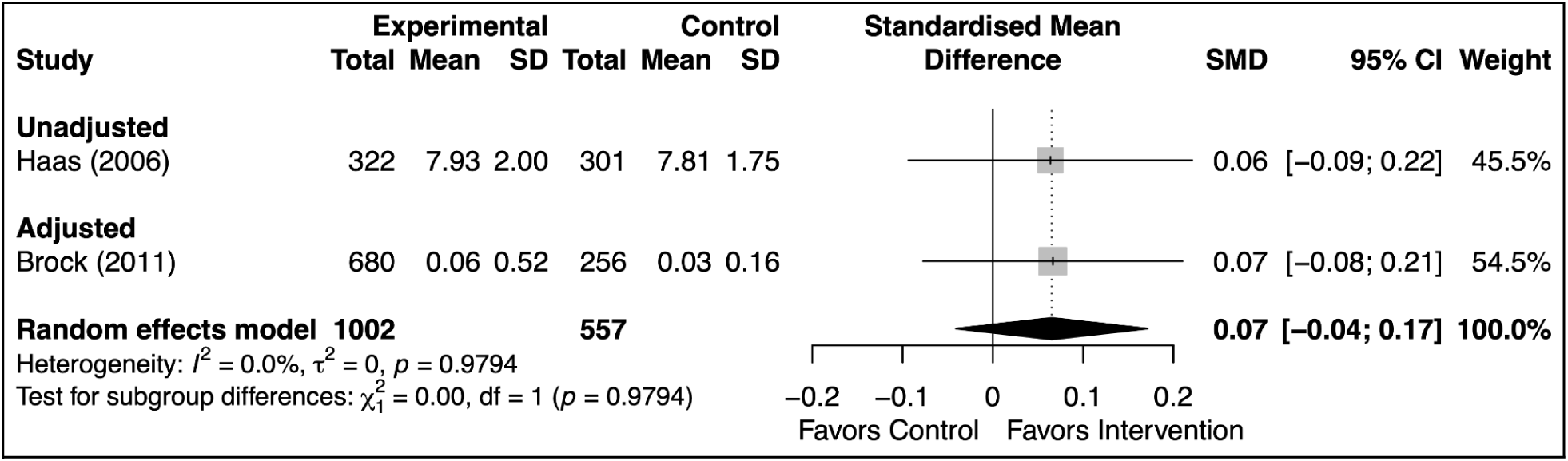
Effects of agenda-setting interventions on initial elicitation of concerns, stratified by adjustment status

**Appendix 5 Figure 13.**
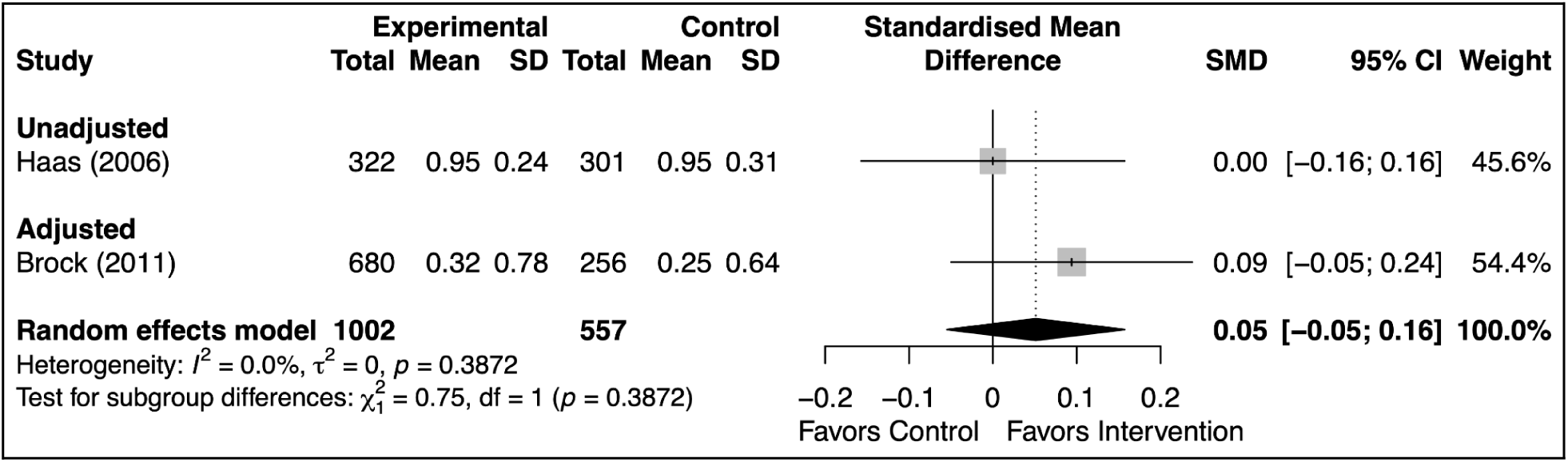
Effects of agenda-setting interventions on elicitation of additional concerns, stratified by adjustment status

**Appendix 5 Figure 14.**
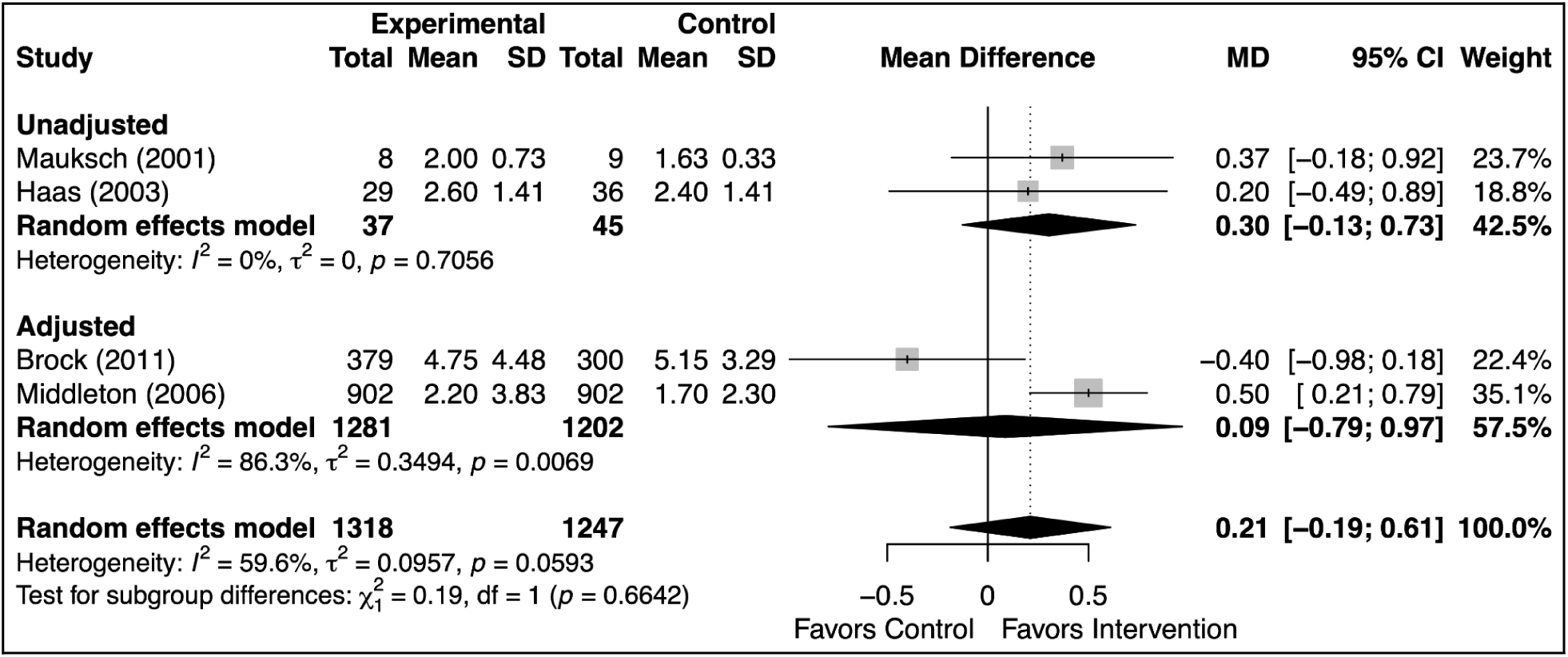
Effects of agenda-setting interventions on number of concerns raised, stratified by adjustment status

**Appendix 5 Figure 15.**
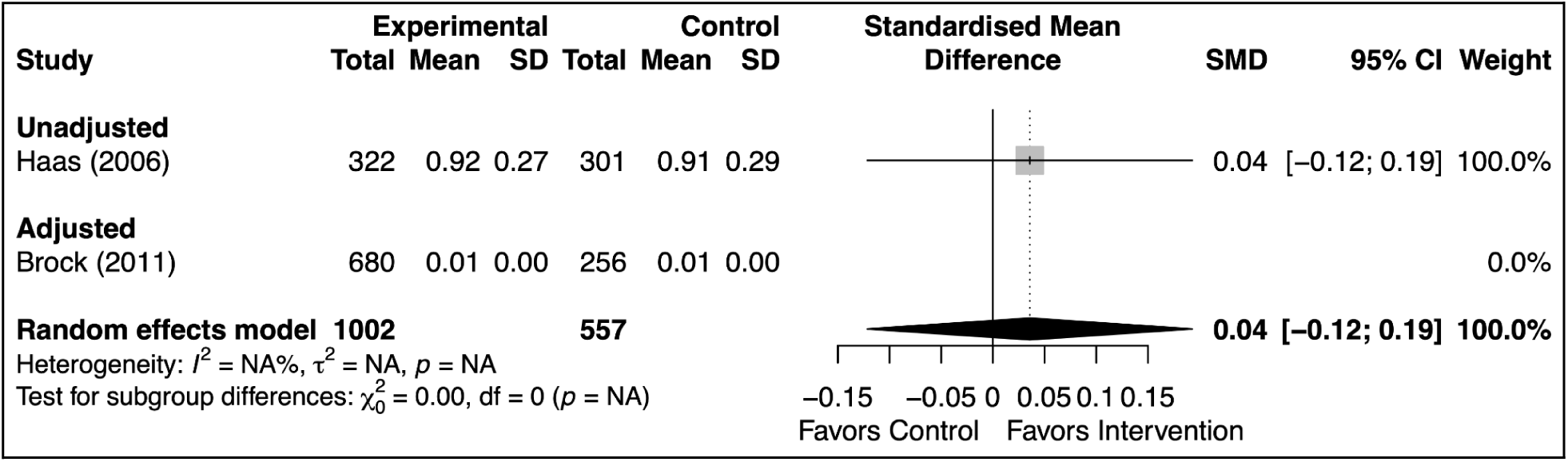
Effects of agenda-setting interventions on prioritization of concerns, stratified by adjustment status

**Appendix 5 Figure 16.**
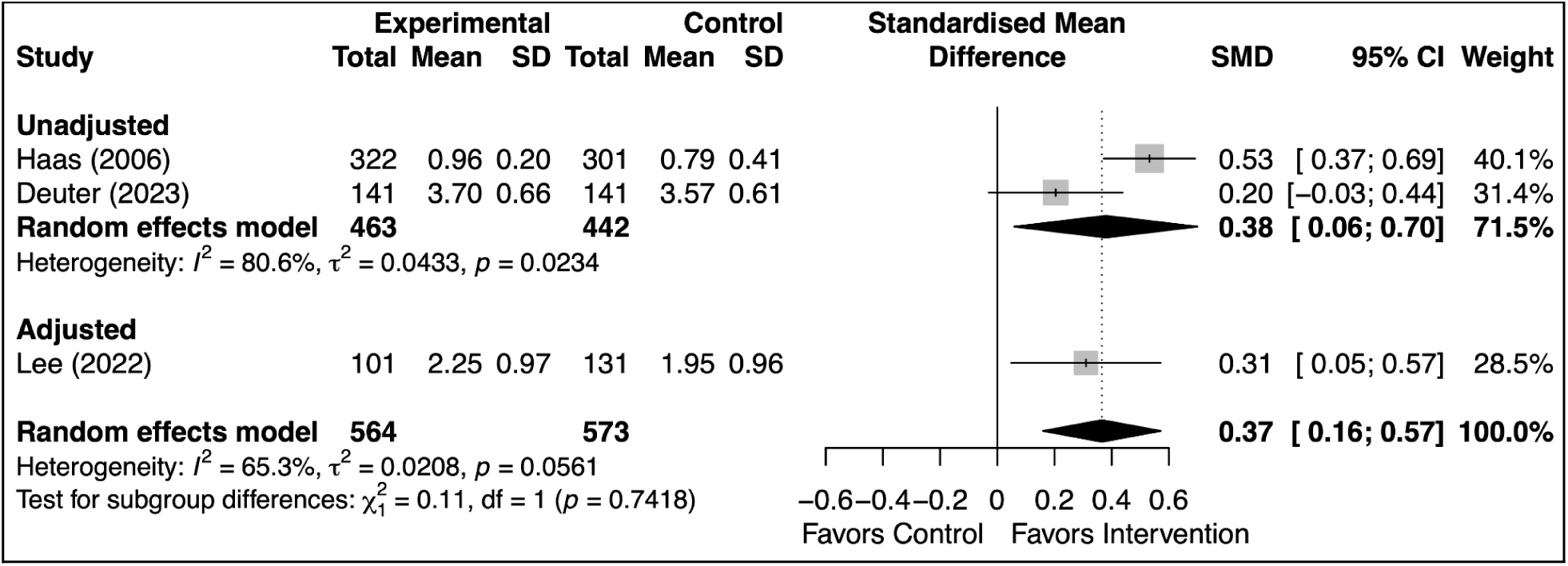
Effects of agenda-setting interventions on concerns addressed, continuous, stratified by adjustment status

**Appendix 5 Figure 17.**
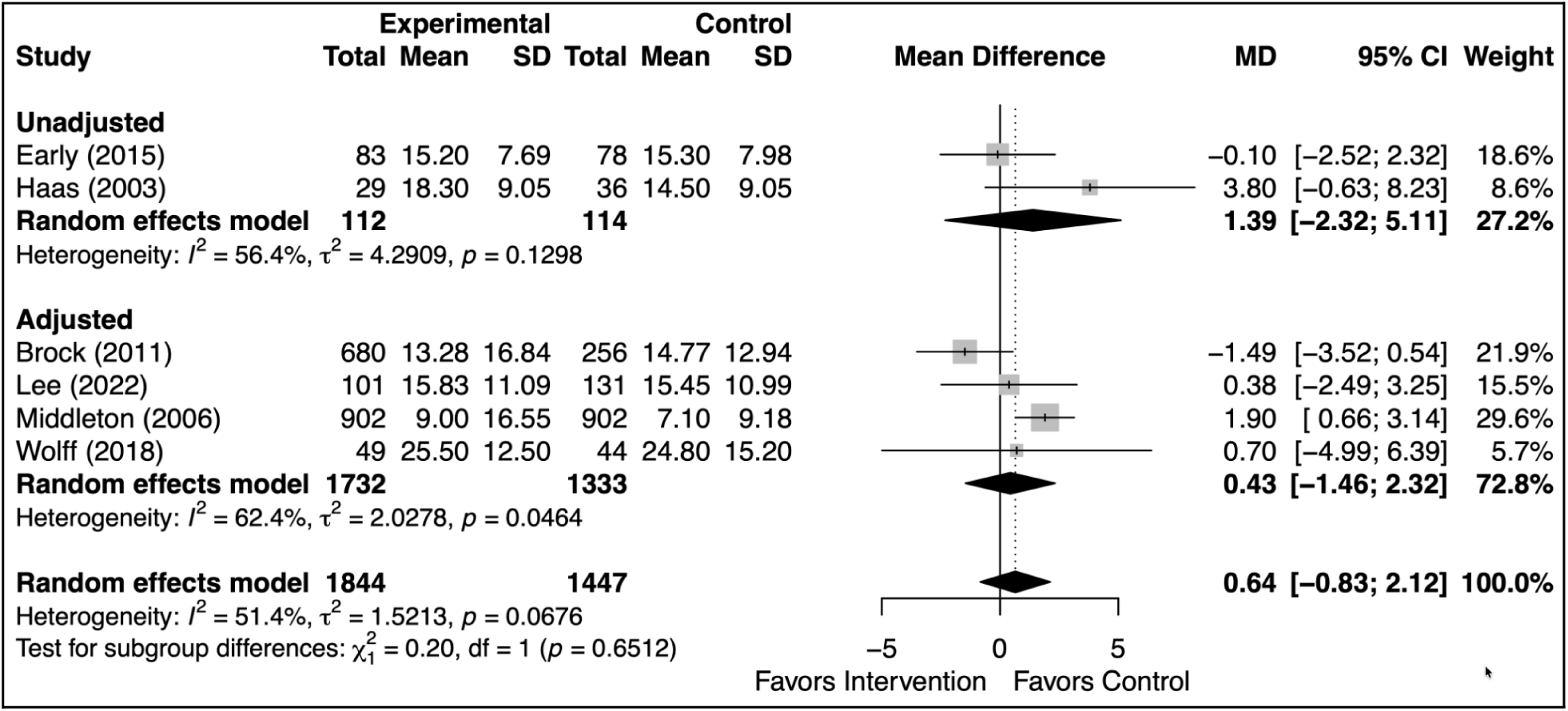
Effects of agenda-setting interventions on visit duration, stratified by adjustment status

**Appendix 5 Figure 18.**
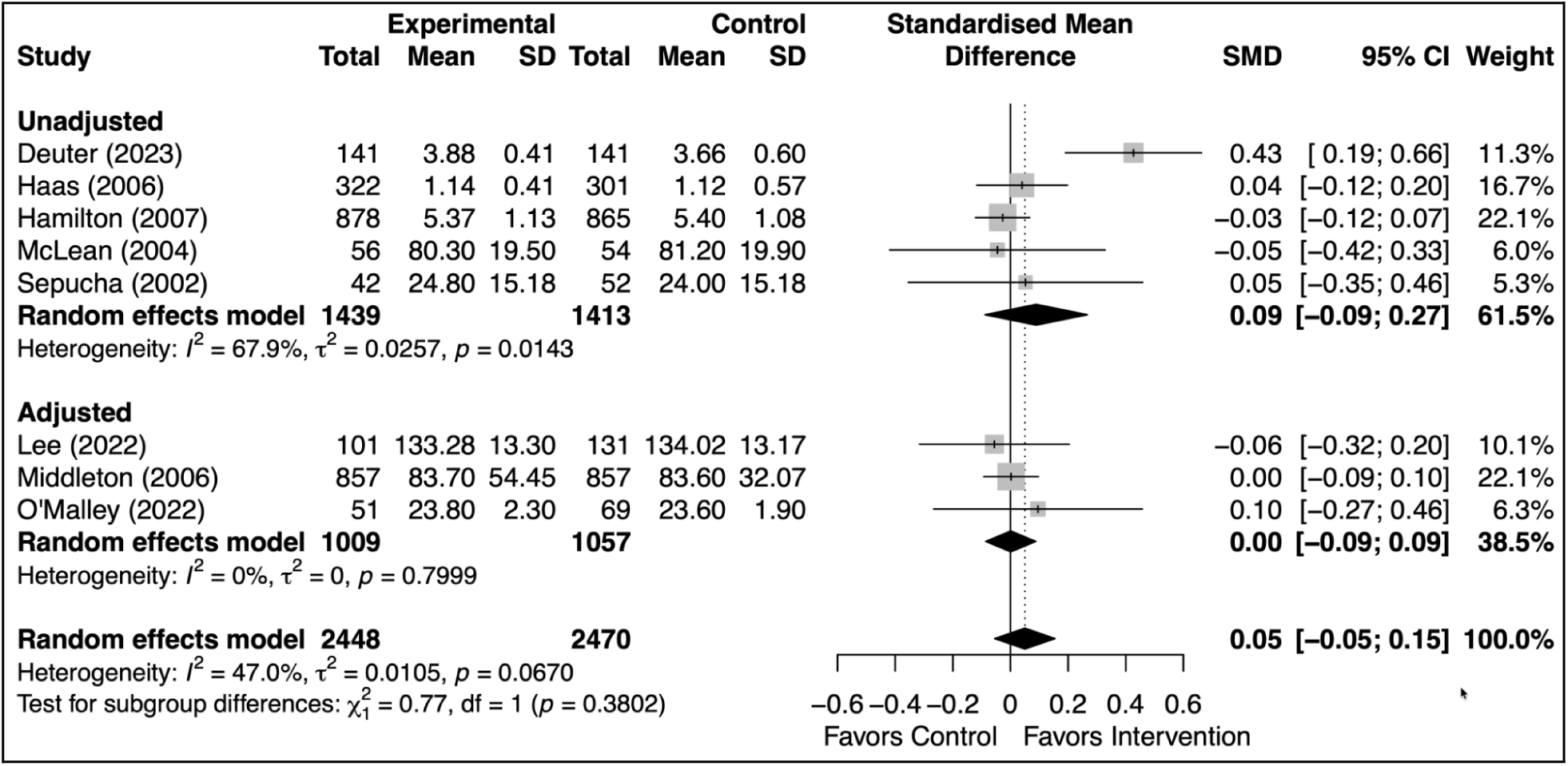
Effects of agenda-setting interventions on overall patient satisfaction, stratified by adjustment status

**Appendix 5 Figure 19.**
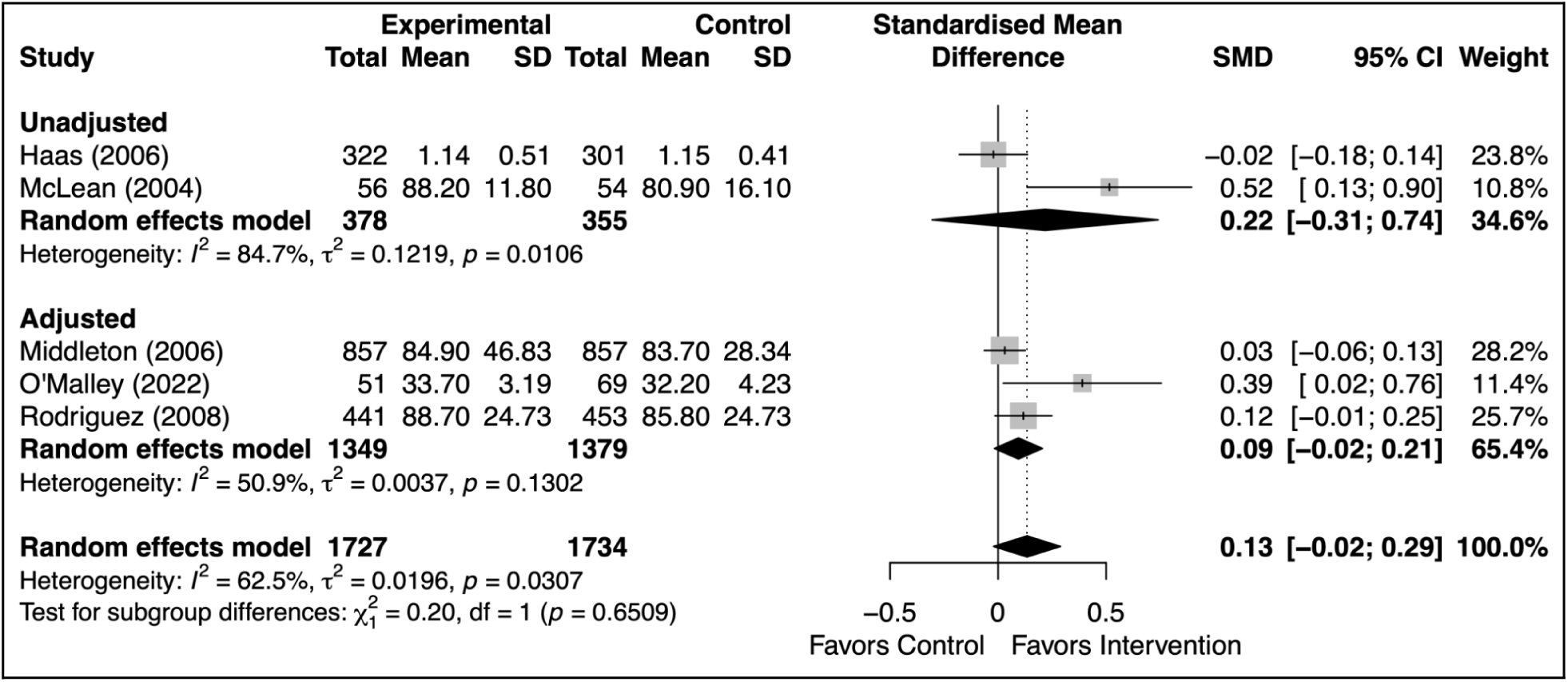
Effects of agenda-setting interventions on satisfaction with clinician, stratified by adjustment status

**Appendix 5 Figure 20.**
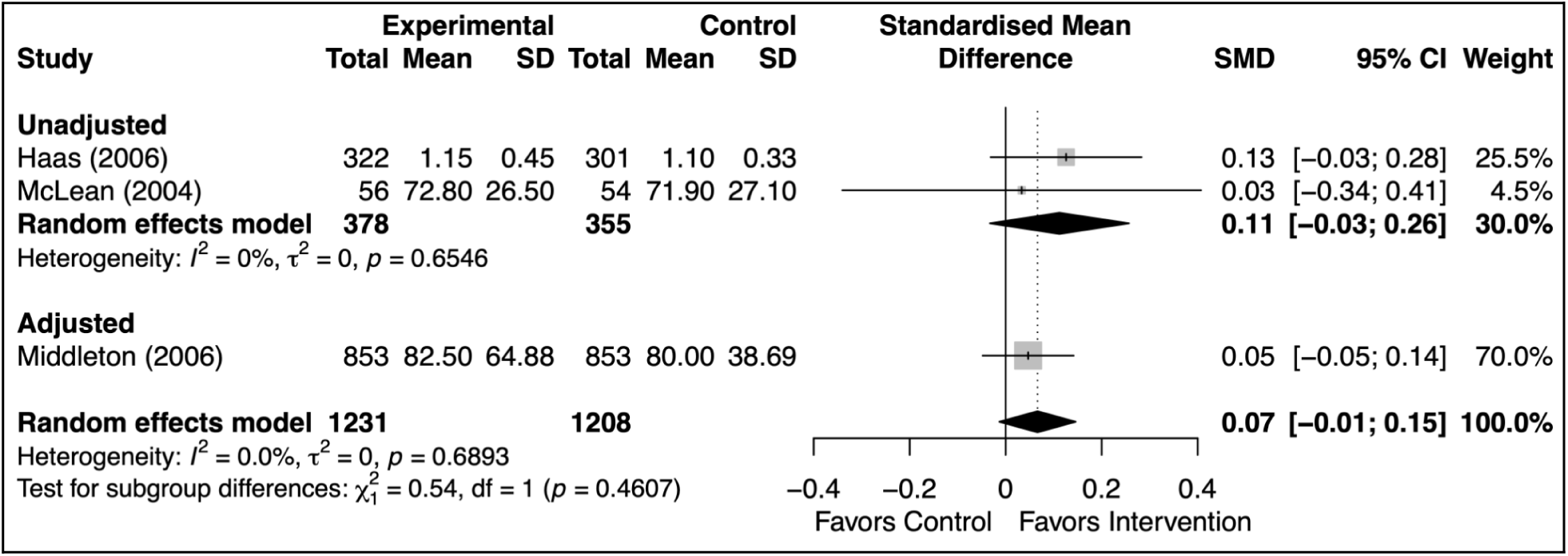
Effects of agenda-setting interventions on adequacy of visit time, stratified by adjustment status

**Appendix 5 Figure 21.**
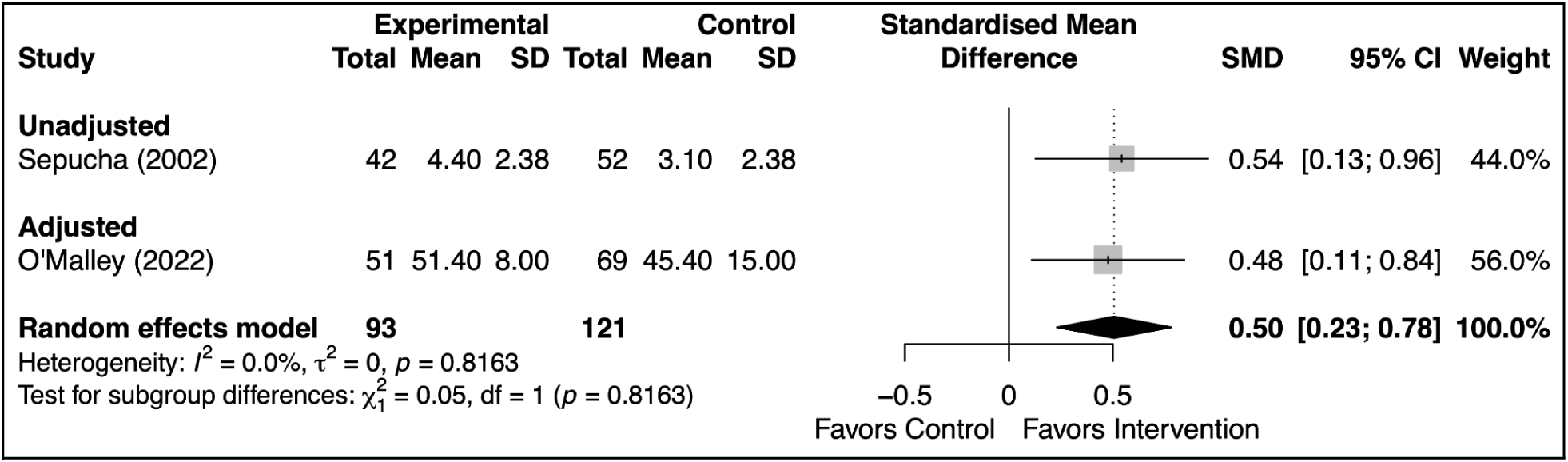
Effects of agenda-setting interventions on overall clinician satisfaction, stratified by adjustment status

### Intervention structure

**Appendix 5 Figure 22.**
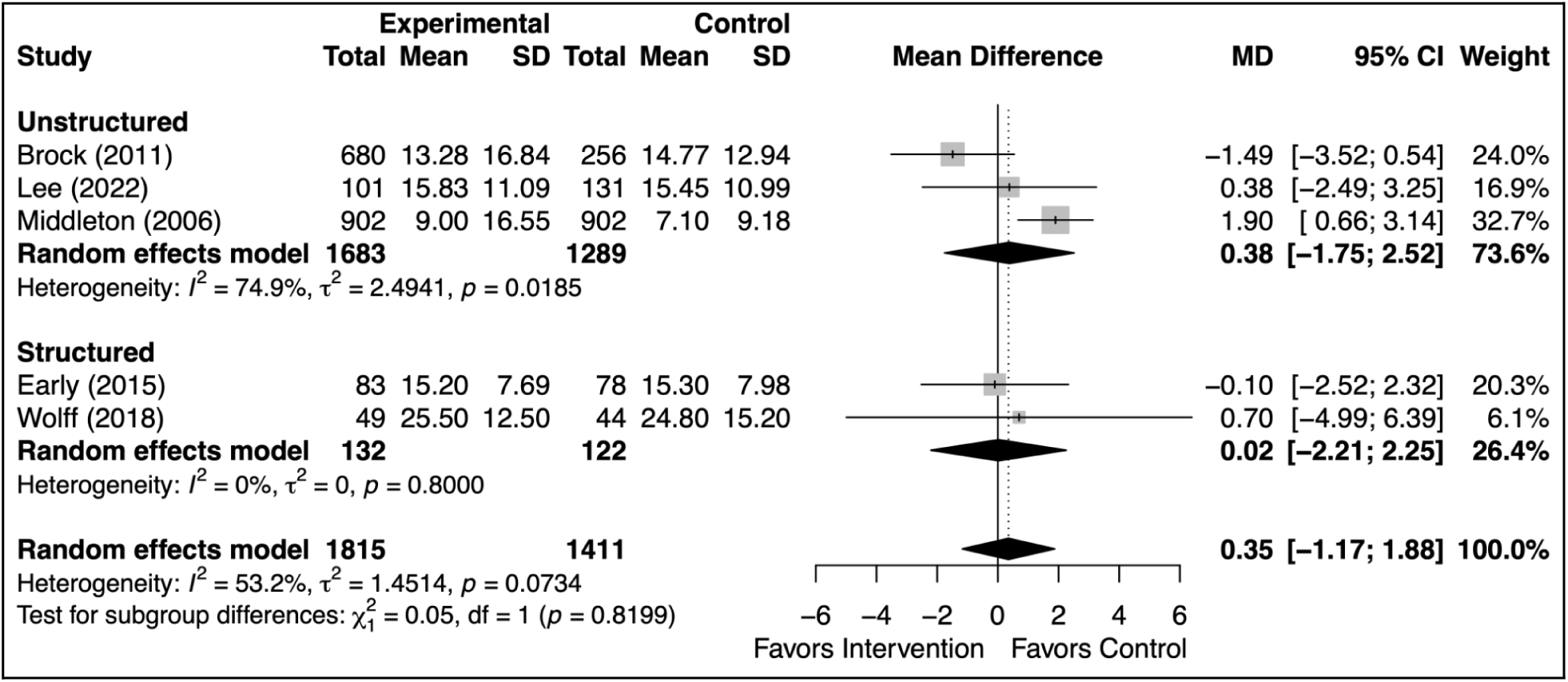
Effects of agenda-setting interventions on visit duration, stratified by intervention structure

**Appendix 5 Figure 23.**
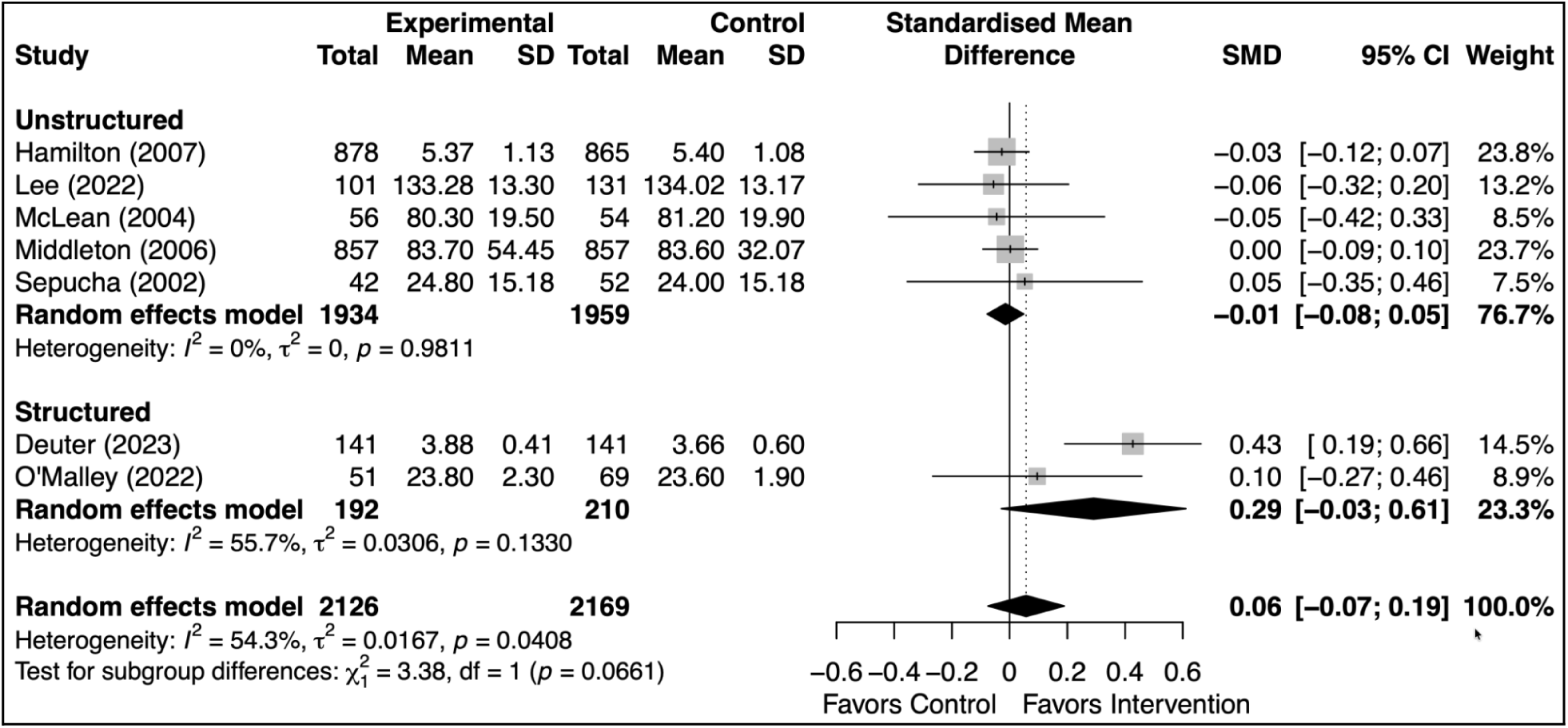
Effects of agenda-setting interventions on overall patient satisfaction, stratified by intervention structure

**Appendix 5 Figure 24.**
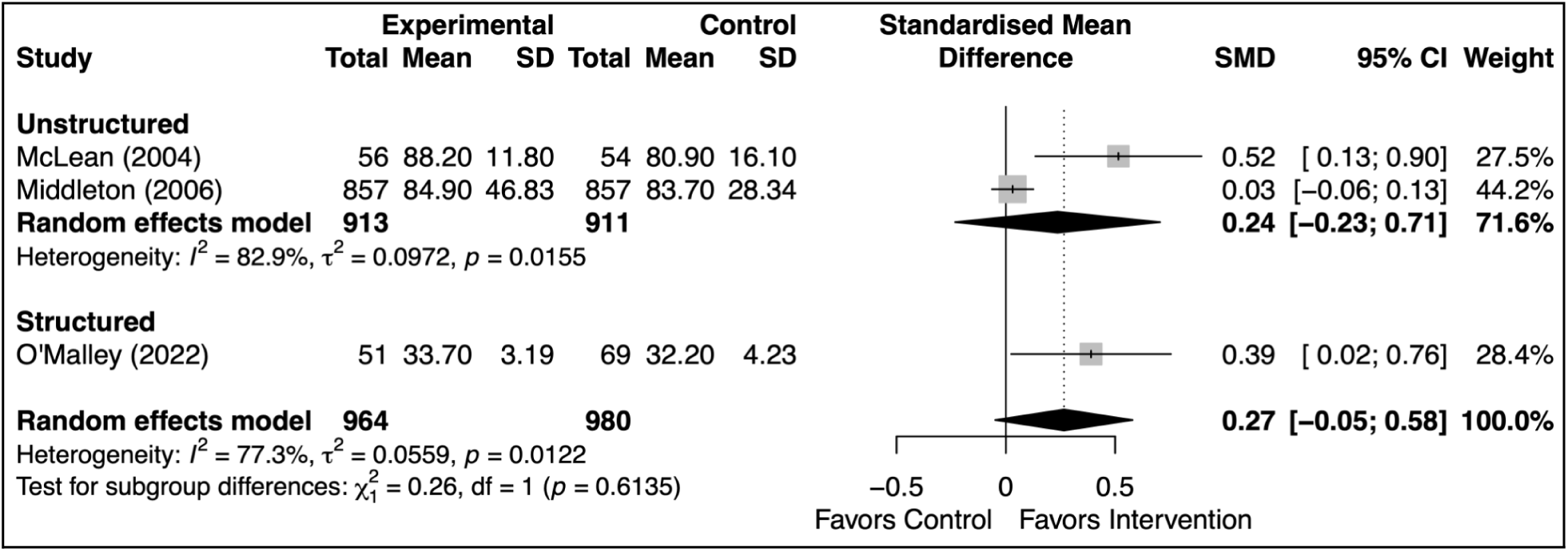
Effects of agenda-setting interventions on satisfaction with clinician, stratified by intervention structure

**Appendix 5 Figure 25.**
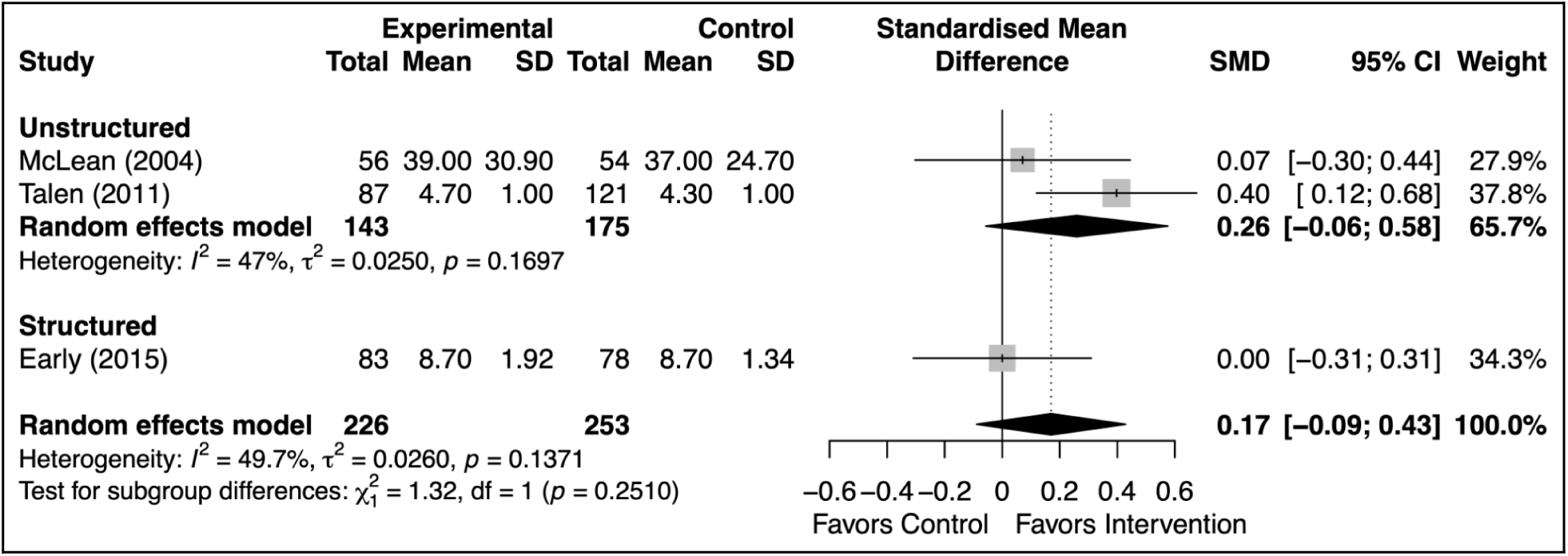
Effects of agenda-setting interventions on patient activation or enablement, stratified by intervention structure

**Appendix 5 Figure 26.**
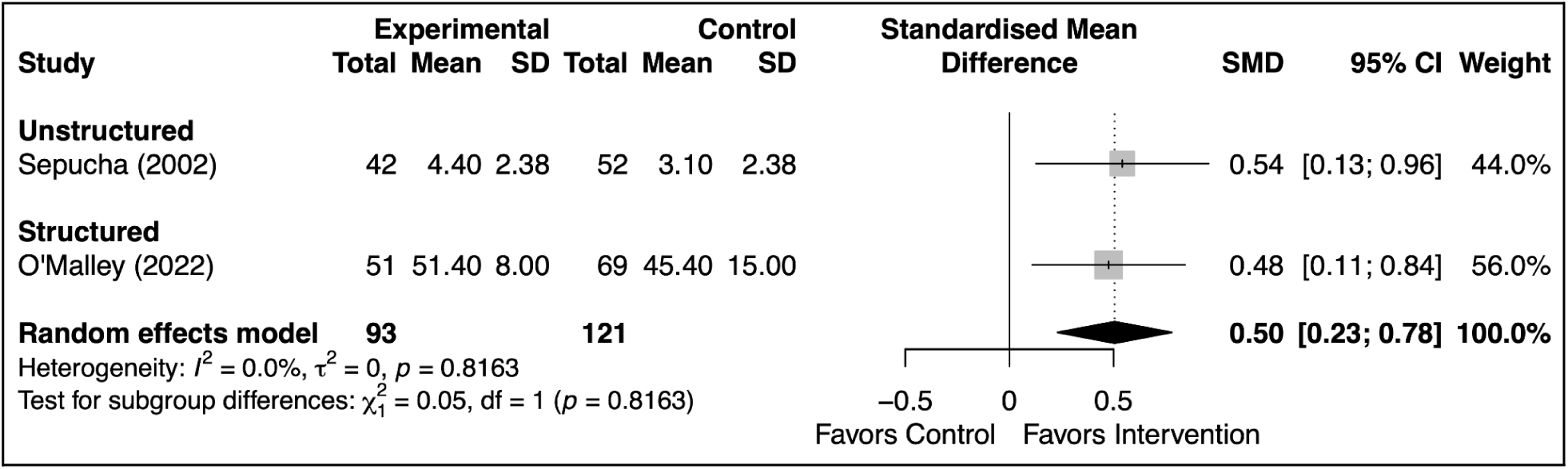
Effects of agenda-setting interventions on overall clinician satisfaction, stratified by intervention structure

## Appendix 6. Assessment of possible small-study effects

**Appendix 6 Figure 1.**
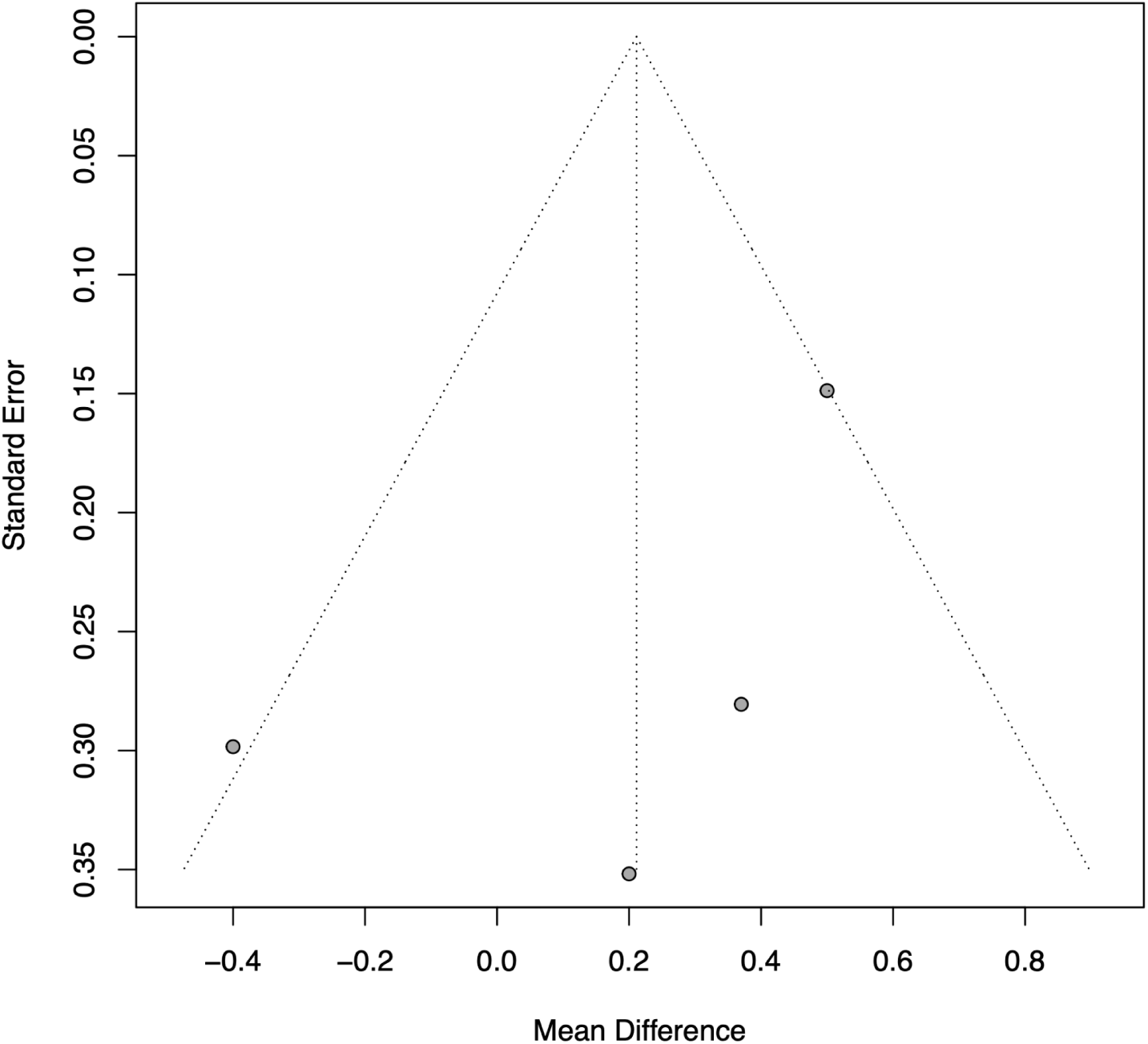
Assessment of possible small-study effects for number of concerns raised

**Appendix 6 Figure 2.**
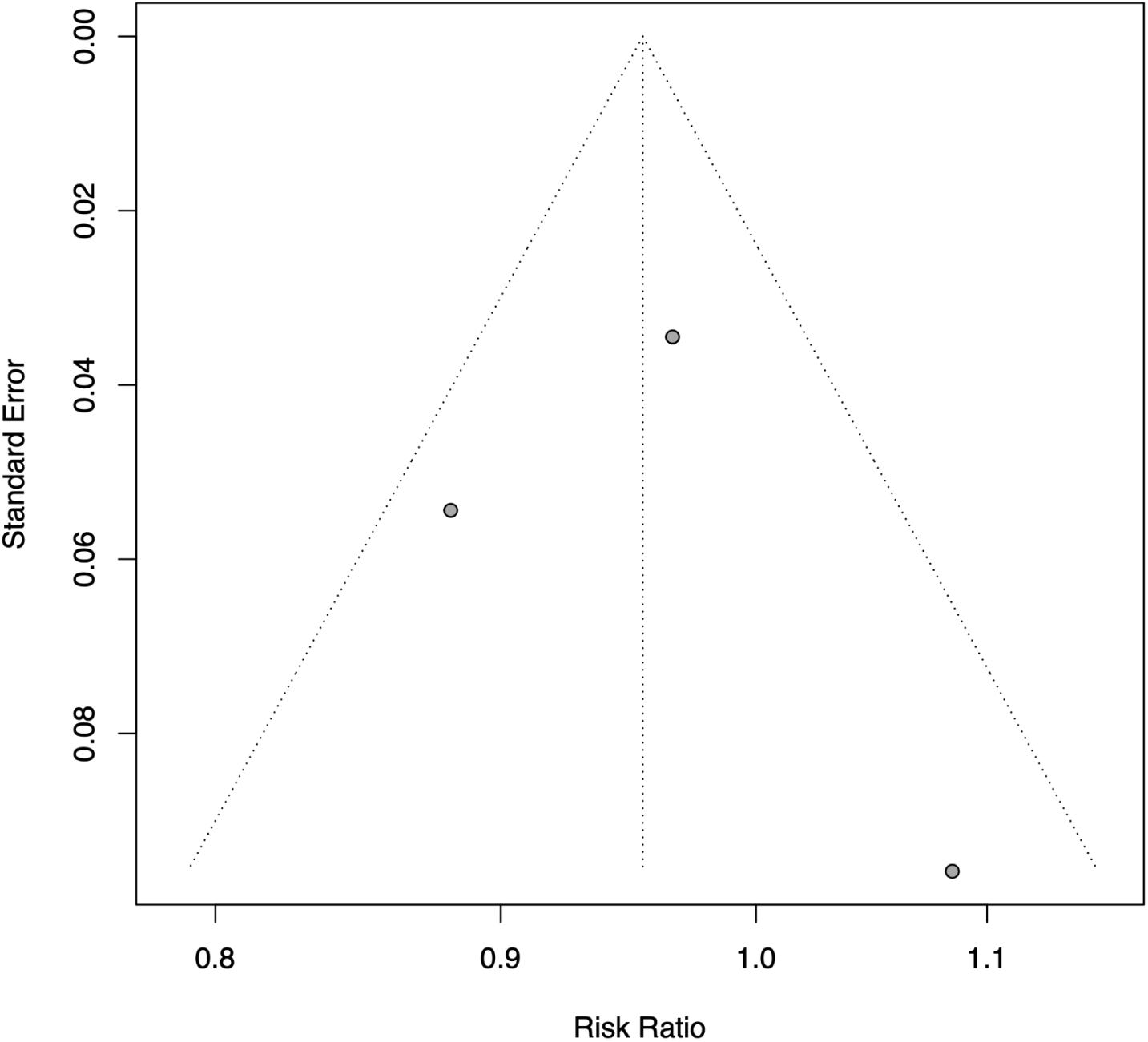
Assessment of possible small-study effects for concerns addressed, dichotomous

**Appendix 6 Figure 3.**
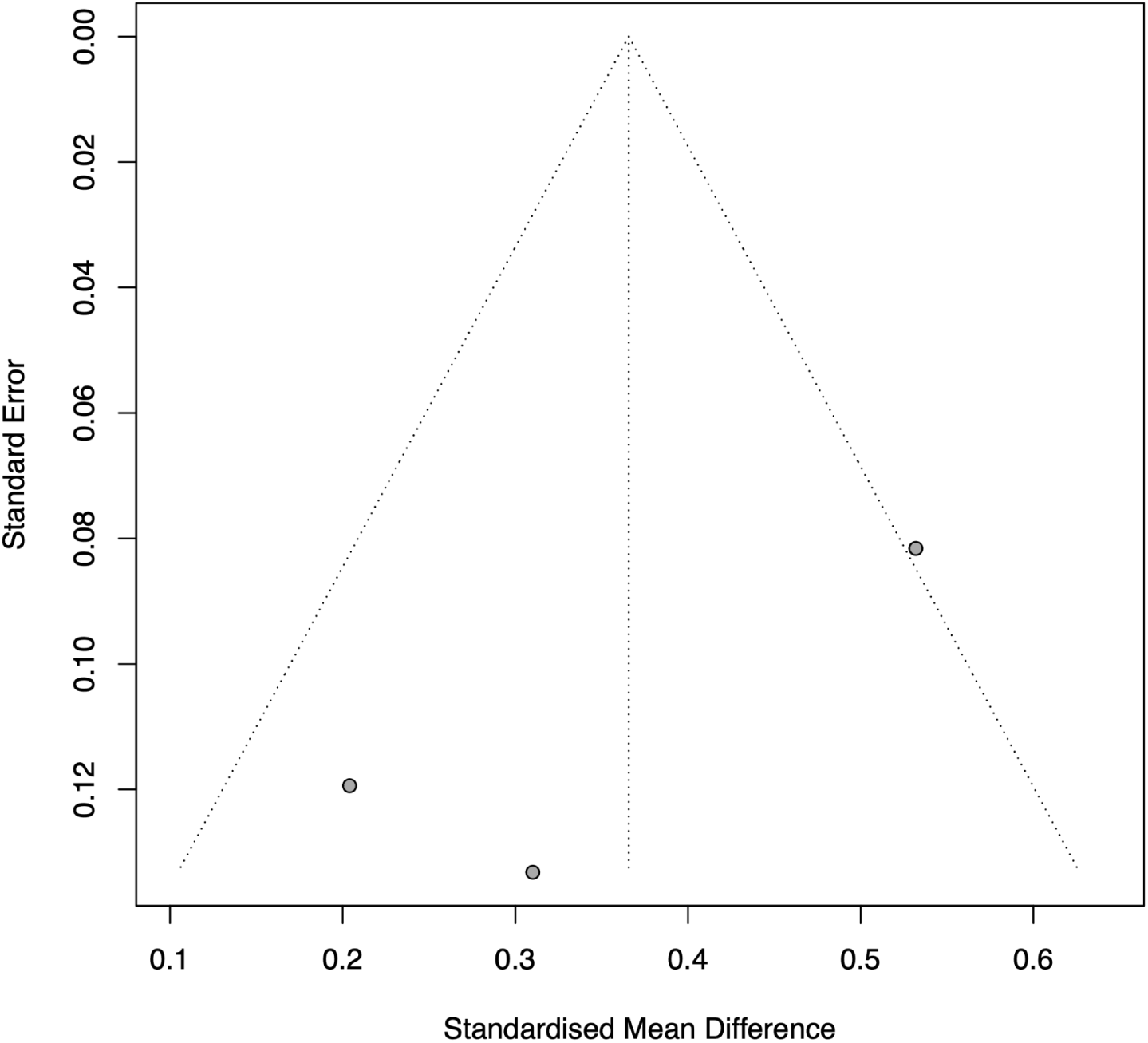
Assessment of possible small-study effects for concerns addressed, continuous

**Appendix 6 Figure 4.**
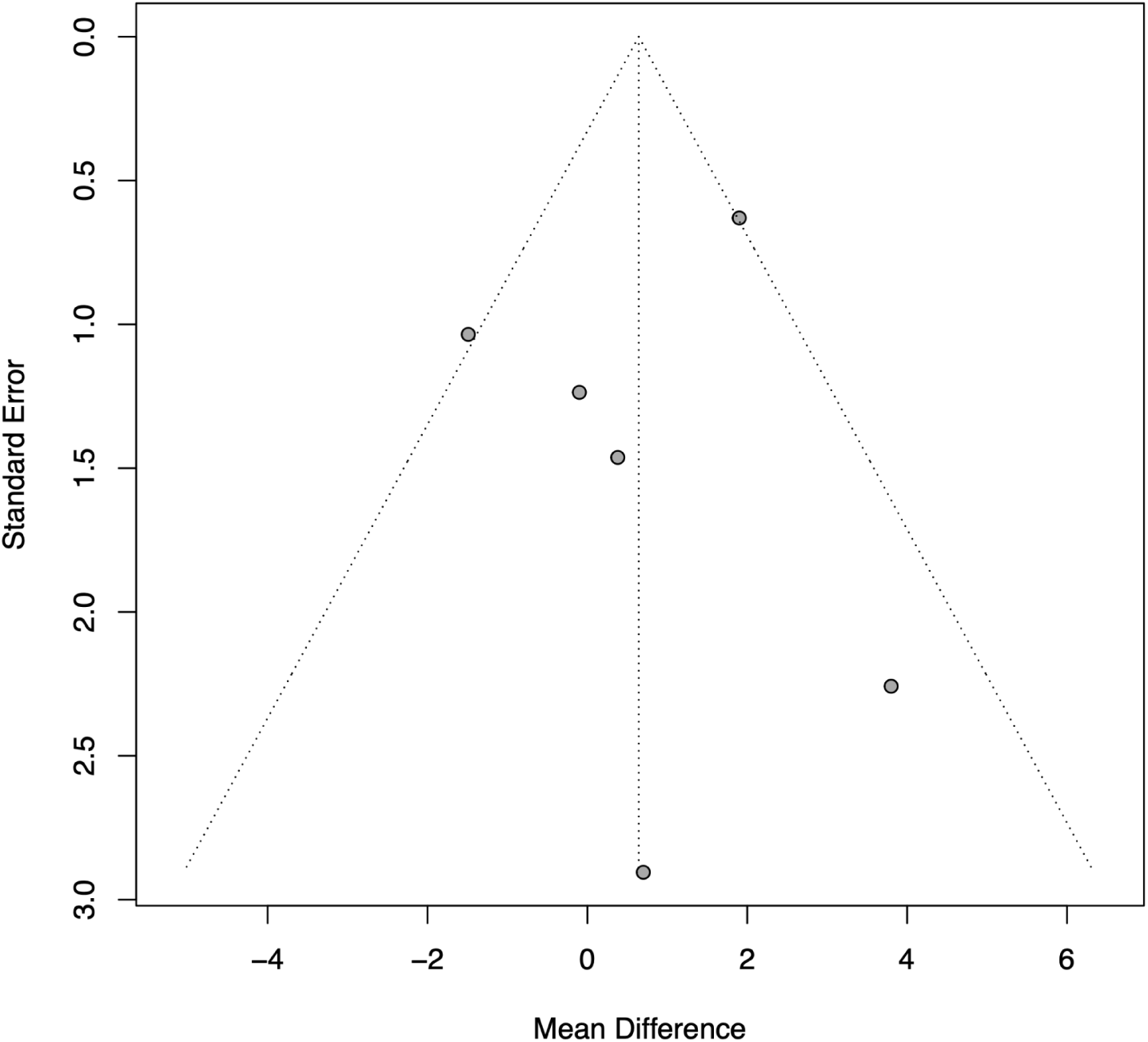
Assessment of possible small-study effects for visit duration

**Appendix 6 Figure 5.**
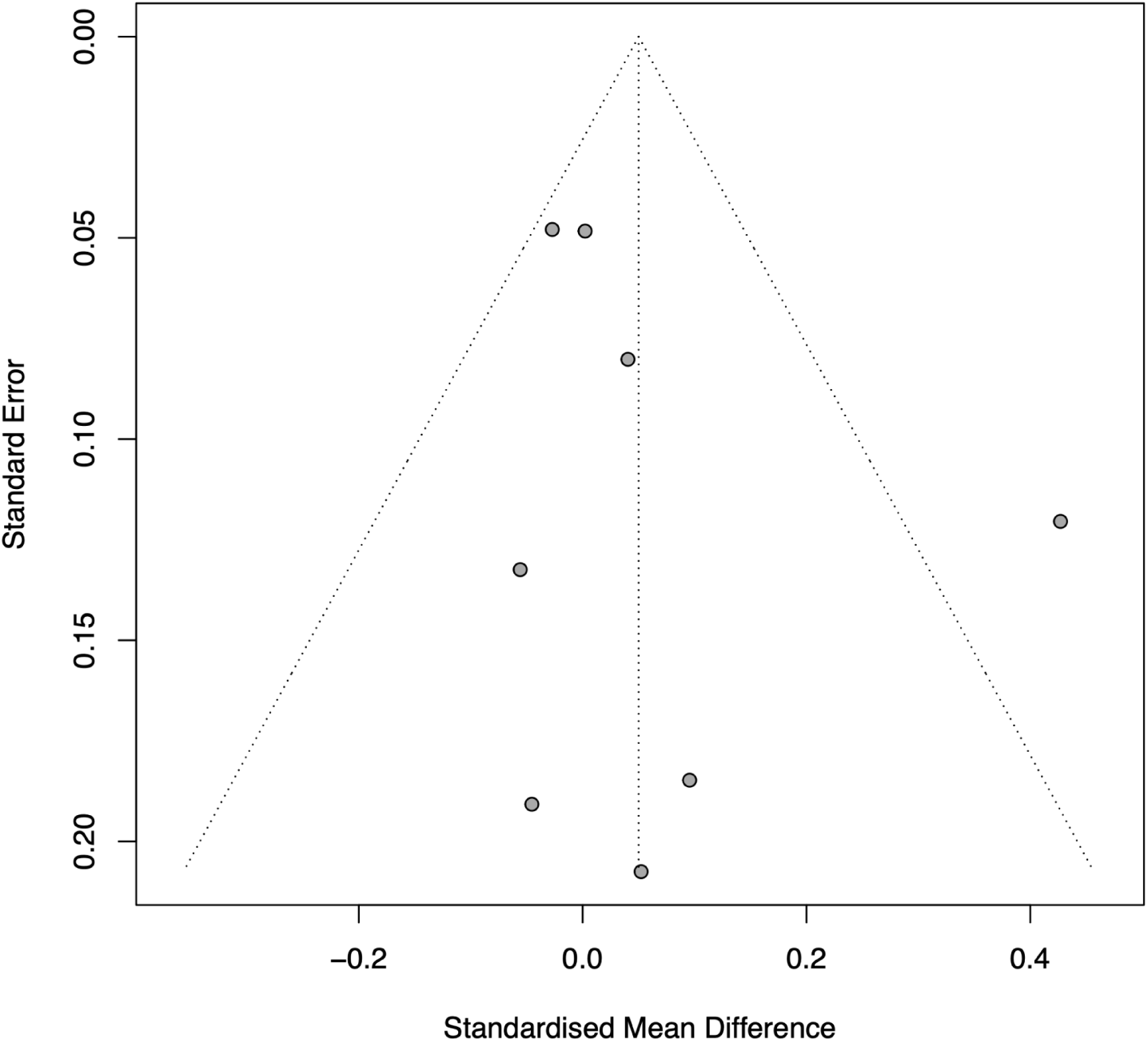
Assessment of possible small-study effects for overall patient satisfaction

**Appendix 6 Figure 6.**
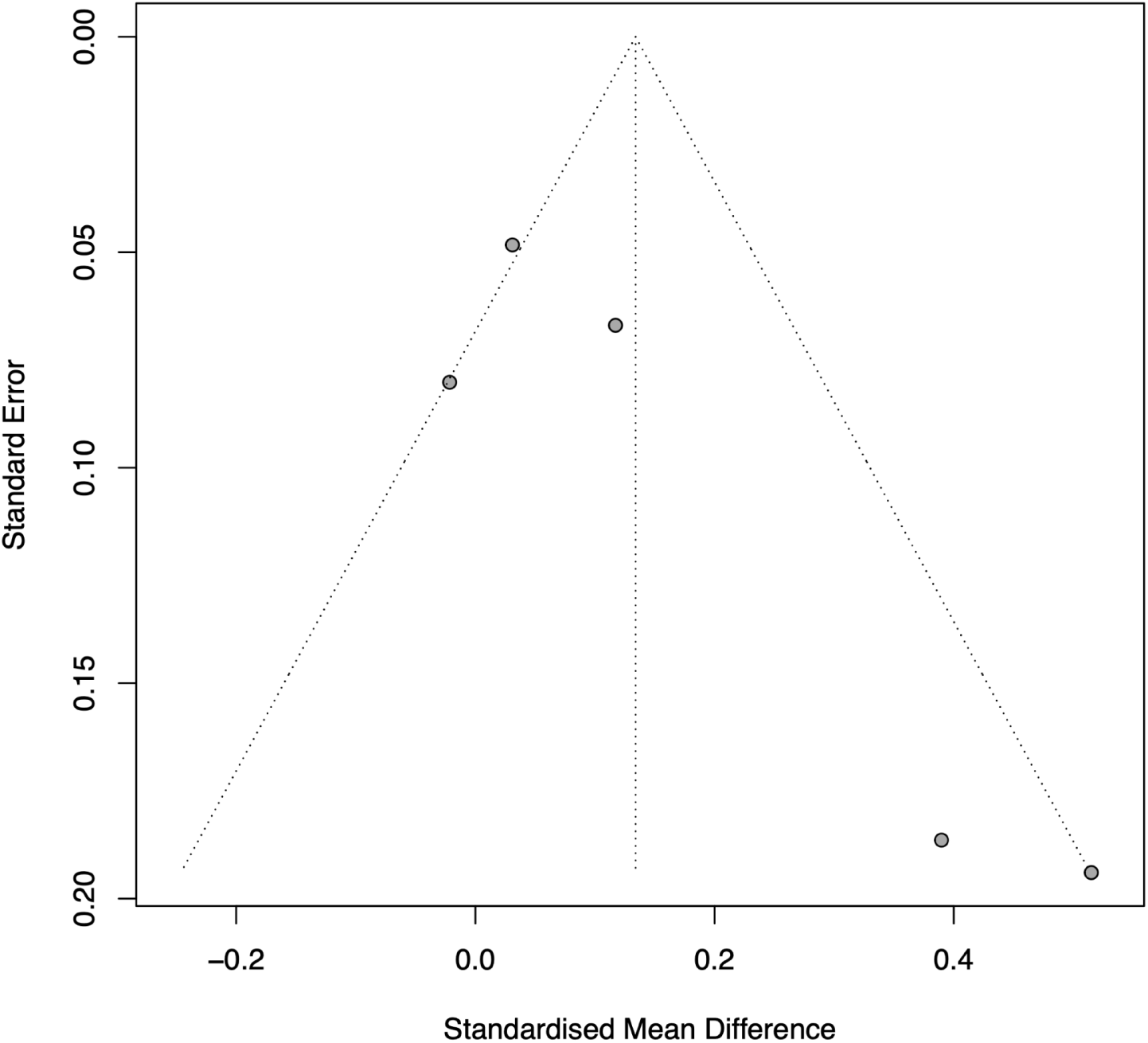
Assessment of possible small-study effects for satisfaction with clinician

**Appendix 6 Figure 7.**
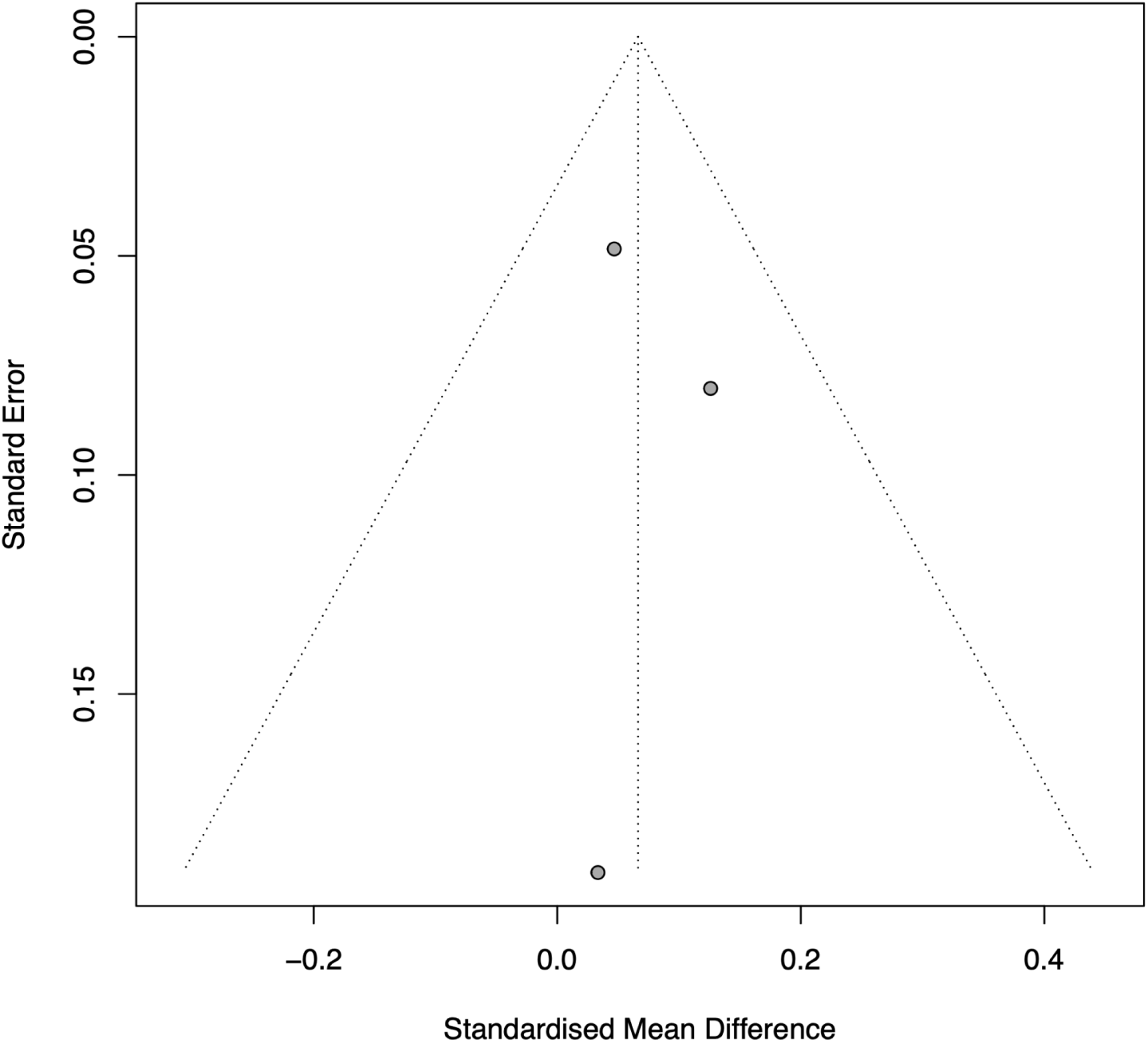
Assessment of possible small-study effects for adequacy of visit time

**Appendix 6 Figure 8.**
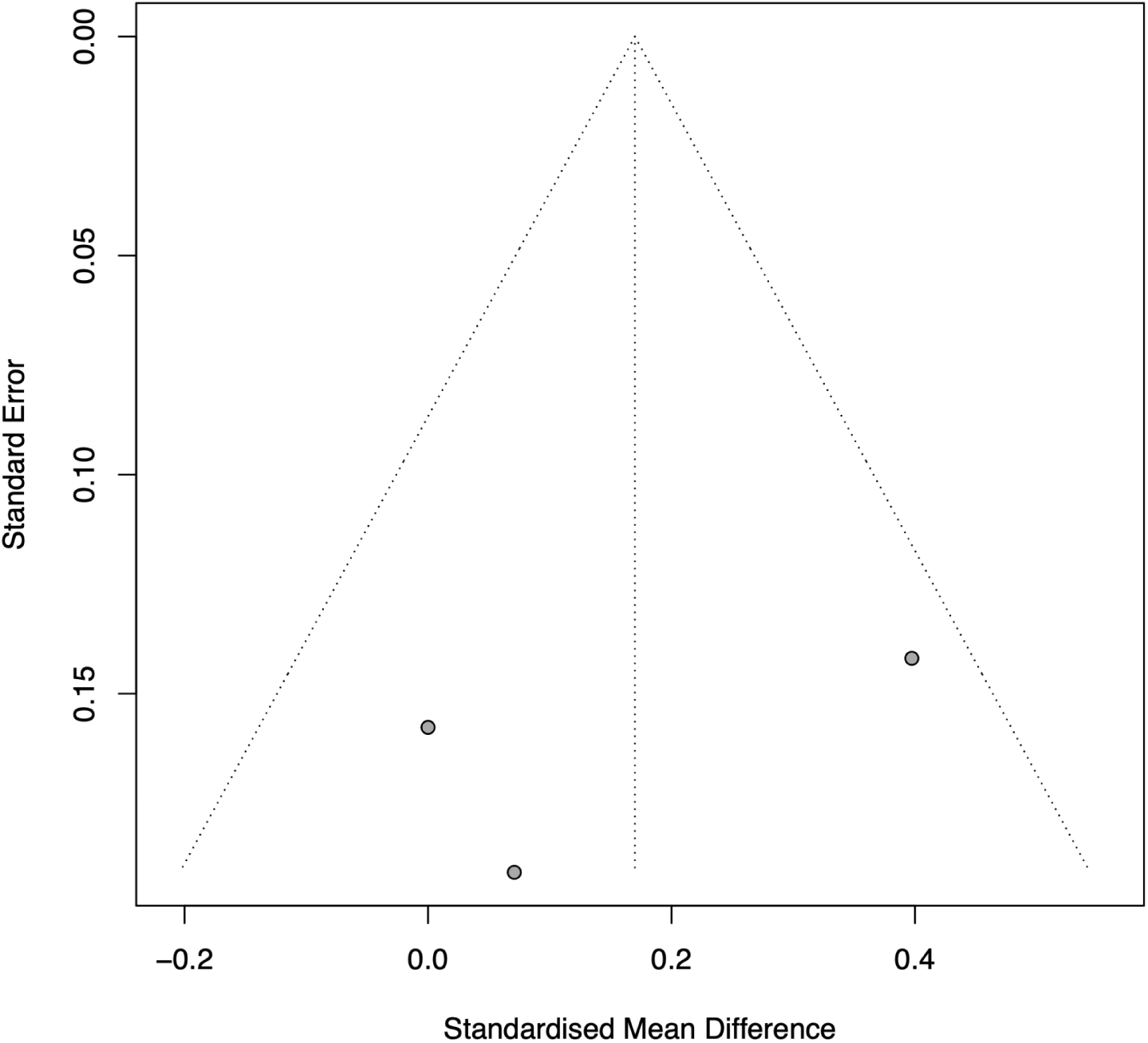
Assessment of possible small-study effects for patient activation or enablement

## Notes

### Clinical Protocols

https://pubmed.ncbi.nlm.nih.gov/39446854/

## References

1. Hood-Medland EA, White AEC, Kravitz RL, Henry SG. Agenda setting and visit openings in primary care visits involving patients taking opioids for chronic pain. BMC Fam Pract. 2021;22:4.

2. Robinson JD, Tate A, Heritage J. Agenda-setting revisited: When and how do primary-care physicians solicit patients’ additional concerns? Patient Educ Couns. 2016;99(5):718–723.

3. Frankel RM ST. Getting the most out of the clinical encounter: the Four Habits Model. Perm J. 1999;3(3):79–88.

4. Wolff JL, Roter DL, Boyd CM, et al. Patient-Family Agenda Setting for Primary Care Patients with Cognitive Impairment: the SAME Page Trial. J Gen Intern Med. 2018;33(9):1478–1486.

5. Rodriguez HP, Anastario MP, Frankel RM, et al. Can teaching agenda-setting skills to physicians improve clinical interaction quality? A controlled intervention. BMC Med Educ. 2008;8:3.

6. Gobat N, Kinnersley P, Gregory JW, Robling M. What is agenda setting in the clinical encounter? Consensus from literature review and expert consultation. Patient Educ Couns. 2015;98(7):822–829.

7. Street RL Jr, Makoul G, Arora NK, Epstein RM. How does communication heal? Pathways linking clinician-patient communication to health outcomes. Patient Educ Couns. 2009;74(3):295–301.

8. Frankel RM, Salyers MP, Bonfils KA, Oles SK, Matthias MS. Agenda setting in psychiatric consultations: an exploratory study. Psychiatr Rehabil J. 2013;36(3):195–201.

9. Baker LH, O’Connell D, Platt FW. “What else?” Setting the agenda for the clinical interview. Ann Intern Med. 2005;143(10):766–770.

10. Brock DM, Mauksch LB, Witteborn S, Hummel J, Nagasawa P, Robins LS. Effectiveness of intensive physician training in upfront agenda setting. J Gen Intern Med. 2011;26(11):1317–1323.

11. Roh H, Park KH, Jeon YJ, Park SG, Lee J. Medical students’ agenda-setting abilities during medical interviews. Korean Journal of Medical Education. 2015;27(2):77–86. doi:10.3946/kjme.2015.27.2.77

12. Coyle AC, Yen RW, Elwyn G. Interrupted opening statements in clinical encounters: A scoping review. Patient Educ Couns. 2022;105(8):2653–2663.

13. Singh Ospina N, Phillips KA, Rodriguez-Gutierrez R, et al. Eliciting the Patient’s Agenda-Secondary Analysis of Recorded Clinical Encounters. J Gen Intern Med. 2019;34(1):36–40.

14. Gobat N, Kinnersley P, Gregory JW, Pickles T, Hood K, Robling M. Measuring clinical skills in agenda-mapping (EAGL-I). Patient Educ Couns. 2015;98(10):1214–1221.

15. Gobat NH. Agenda Setting in the Clinical Encounter: What Is It, and Is It Measureable? phd. Cardiff University; 2014. Accessed December 19, 2022. https://orca.cardiff.ac.uk/id/eprint/56395

16. Early F, Everden AJ, O’Brien CM, Fagan PL, Fuld JP. Patient agenda setting in respiratory outpatients: A randomized controlled trial. Chron Respir Dis. 2015;12(4):347–356.

17. Hamilton W, Russell D, Stabb C, Seamark D, Campion-Smith C, Britten N. The effect of patient self-completion agenda forms on prescribing and adherence in general practice: a randomized controlled trial. Fam Pract. 2007;24(1):77–83.

18. Kuhle CS, Truitt F, Steffen M, et al. Improving patient-centered care: agenda-setting in occupational medicine. J Occup Environ Med. 2013;55(5):479–482.

19. Lee YK, Ng CJ, Syahirah MR, et al. Effectiveness of a web-based, electronic medical records-integrated patient agenda tool to improve doctor-patient communication in primary care consultations: A pragmatic cluster-randomized controlled trial study. Int J Med Inform. 2022;162:104761.

20. Munch L, Stensgaard S, Feinberg MB, Elwyn G, Lomborg K. Evaluating the effect of Conversation Cards on agenda-setting in annual diabetes status visits: A multi-method study. Patient Educ Couns. 2024;119(108084):108084.

21. O’Malley PG, Jackson JL, Becher D, Hanson J, Lee JK, Grace KA. Tool to improve patient-provider interactions in adult primary care: Randomized controlled pilot study. Can Fam Physician. 2022;68(2):e49–e58.

22. Robling M, McNamara R, Bennert K, et al. The effect of the Talking Diabetes consulting skills intervention on glycaemic control and quality of life in children with type 1 diabetes: cluster randomised controlled trial (DEPICTED study). BMJ. 2012;344:e2359.

23. Wolff JL, Aufill J, Echavarria D, et al. Sharing in care: engaging care partners in the care and communication of breast cancer patients. Breast Cancer Res Treat. 2019;177(1):127–136.

24. Ditton-Phare P, Sandhu H, Kelly B, Kissane D, Loughland C. Pilot evaluation of a communication skills training program for psychiatry residents using standardized patient assessment. Acad Psychiatry. 2016;40(5):768–775.

25. Haas LJ, Houchins J, Leiser JP. Changing family physicians’ visit structuring behavior: a pilot study. Fam Med. 2003;35(10):726–729.

26. Haas LJ, Glazer K, Houchins J, Terry S. Improving the effectiveness of the medical visit: a brief visit-structuring workshop changes patients’ perceptions of primary care visits. Patient Educ Couns. 2006;62(3):374–378.

27. Leydon GM, Stuart B, Summers RH, et al. Findings from a feasibility study to improve GP elicitation of patient concerns in UK general practice consultations. Patient Educ Couns. 2018;101(8):1394–1402.

28. Pritt SE. Utilizing a Patient-Centered Communication Intervention to Reduce Unmet Patient Concerns in Chronic Illness Primary Care. Doctor of Psychology. Indiana University of Pennsylvania; 2020. https://search.proquest.com/openview/e8e766d35bcc0397626318070c359cc6/1?pq-origsite=gscholar&cbl=44156

29. Middleton JF, McKinley RK, Gillies CL. Effect of patient completed agenda forms and doctors’ education about the agenda on the outcome of consultations: randomised controlled trial. BMJ. 2006;332(7552):1238–1242.

30. Everden A, Early F, Homan K, Fuld J. P94 Patient Agenda Setting And Clinic Efficiency In Outpatients: An Individual Randomised Controlled Trial. Thorax. 2014;69(Suppl 2):A118–A118.

31. Sierpe A, Yen RW, Stevens G, Van Citters AD, Elwyn G, Saunders CH. Agenda-setting in the clinical encounter: A systematic review protocol. PLoS One. 2024;19(10):e0312613.

32. PROSPERO. Accessed August 23, 2026. https://www.crd.york.ac.uk/PROSPERO/view/CRD42023468045

33. Higgins JPT, Thomas J, Chandler J, et al. Cochrane Handbook for Systematic Reviews of Interventions. John Wiley & Sons; 2019.

34. Page MJ, McKenzie JE, Bossuyt PM, et al. The PRISMA 2020 statement: an updated guideline for reporting systematic reviews. BMJ. 2021;372:n71.

35. Rey-Bellet S, Dubois J, Vannotti M, et al. Agenda Setting During Follow-Up Encounters in a University Primary Care Outpatient Clinic. Health Commun. 2017;32(6):714–720.

36. Kowalski CP, McQuillan DB, Chawla N, et al. “The Hand on the Doorknob”: Visit Agenda Setting by Complex Patients and Their Primary Care Physicians. J Am Board Fam Med. 2018;31(1):29–37.

37. Marvel MK, Epstein RM, Flowers K, Beckman HB. Soliciting the patient’s agenda: have we improved? JAMA. 1999;281(3):283–287.

38. Ouzzani M, Hammady H, Fedorowicz Z, Elmagarmid A. Rayyan—a web and mobile app for systematic reviews. Syst Rev. 2016;5(1):210.

39. Hoffmann TC, Glasziou PP, Boutron I, et al. Better reporting of interventions: template for intervention description and replication (TIDieR) checklist and guide. BMJ. 2014;348:g1687.

40. Sterne JAC, Savović J, Page MJ, et al. RoB 2: a revised tool for assessing risk of bias in randomised trials. BMJ. 2019;366:l4898.

41. RoB 2 for cluster-randomized trials. Accessed August 24, 2026. https://www.riskofbias.info/welcome/rob-2-0-tool/rob-2-for-cluster-randomized-trials

42. Sterne JA, Hernán MA, Reeves BC, et al. ROBINS-I: a tool for assessing risk of bias in non-randomised studies of interventions. BMJ. 2016;355:i4919.

43. Balduzzi S, Rücker G, Schwarzer G. How to perform a meta-analysis with R: a practical tutorial. Evid Based Ment Health. 2019;22(4):153–160.

44. Viechtbauer W. Conducting meta-analyses in R with the metafor package. J Stat Softw. 2010;36(3). doi:10.18637/jss.v036.i03

45. R Core Team. R: A Language and Environment for Statistical Computing. R Foundation for Statistical Computing. doi:10.32614/R.manuals

46. Higgins JPT, Thompson SG. Quantifying heterogeneity in a meta-analysis. Stat Med. 2002;21(11):1539–1558.

47. Chapter 10: Analysing data and undertaking meta-analyses. Accessed March 2, 2023. https://training.cochrane.org/handbook/current/chapter-10

48. Campbell M, McKenzie JE, Sowden A, et al. Synthesis without meta-analysis (SWiM) in systematic reviews: reporting guideline. BMJ. 2020;368:l6890.

49. GRADE handbook. Accessed June 1, 2023. https://gdt.gradepro.org/app/handbook/handbook.html

50. Gregory J, Robling M, DEPICTED Study Group. Evaluating the effect of a blended learning programme to improve consultation skills of paediatric diabetes clinic staff (the DEPICTED study). Pediatric Diabetes. 2010;11 (Suppl. 14):61–62.

51. Gregory J, Robling M, Bennert K, et al. Development and evaluation by a cluster randomised trial of a psychosocial intervention in children and teenagers experiencing diabetes: the DEPICTED study. Health Technol Assess. 2011;15(29):1–202.

52. Middleton J. Eliciting the patient’s agenda: A workshop for general practitioners. Education for general practice. 1998;9(2):231–234.

53. Middleton J, McKinley R. GPs value the opportunity to develop their clinical skills. Education for General Practice. 2000;11(3):307–311.

54. O’Malley PG, Becher D, Hanson J, et al. Impact of a patient activation tool to improve agenda setting in chronic disease encounters: A randomized controlled trial. Journal of General Internal Medicine. Published online 2012.

55. Stuart B, Leydon G, Woods C, et al. The elicitation and management of multiple health concerns in GP consultations. Patient Educ Couns. 2019;102(4):687–693.

56. Sepucha KR, Belkora JK, Mutchnick S, Esserman LJ. Consultation planning to help breast cancer patients prepare for medical consultations: effect on communication and satisfaction for patients and physicians. J Clin Oncol. 2002;20(11):2695–2700.

57. Talen MR, Muller-Held CF, Eshleman KG, Stephens L. Patients’ communication with doctors: a randomized control study of a brief patient communication intervention. Fam Syst Health. 2011;29(3):171–183.

58. Mauksch LB, Hillenburg L, Robins L. The Establishing Focus protocol: Training for collaborative agenda setting and time management in the medical interview. Fam Syst Health. 2001;19(2):147–157.

59. McLean M, Armstrong D. Eliciting patients’ concerns: a randomised controlled trial of different approaches by the doctor. Br J Gen Pract. 2004;54(506):663–666.

60. Deuter M, Martinez C, Preikschat B, et al. Effectiveness and Clinical Usefulness of Electronic Agenda-setting in Psychiatric Practices: A South Texas Psychiatric PBRN Study. J Psychiatr Pract. 2023;29(1):31–37.

61. Mauksch LB, Dugdale DC, Dodson S, Epstein R. Relationship, communication, and efficiency in the medical encounter: creating a clinical model from a literature review. Arch Intern Med. 2008;168(13):1387–1395.

62. Allgood S, Park J, Soleiman K, et al. Taxonomy and effectiveness of clinician agenda-setting questions in routine ambulatory encounters: A mixed method study. Patient Educ Couns. 2023;115:107889.

63. Wollney EN, Vasquez TS, Fisher CL, et al. A systematic scoping review of patient and caregiver self-report measures of satisfaction with clinicians’ communication. Patient Educ Couns. 2023;117(107976):107976.

64. Zill JM, Christalle E, Müller E, Härter M, Dirmaier J, Scholl I. Measurement of physician-patient communication--a systematic review. PLoS One. 2014;9(12):e112637.

65. Wang SJ, Hu WY, Chang YC. Question prompt list intervention for patients with advanced cancer: a systematic review and meta-analysis. Support Care Cancer. 2024;32(4):231.

66. Ting YY, Ey JD, Treloar EC, Reid JL, Bradshaw EL, Maddern GJ. Patient prompts in surgical consultations: A systematic review. Surgery. 2022;172(6):1759–1767.

67. Licqurish SM, Cook OY, Pattuwage LP, et al. Tools to facilitate communication during physician-patient consultations in cancer care: An overview of systematic reviews. CA Cancer J Clin. 2019;69(6):497–520.

68. Brandes K, Linn AJ, Butow PN, van Weert JCM. The characteristics and effectiveness of Question Prompt List interventions in oncology: a systematic review of the literature: QPL interventions in oncology. Psychooncology. 2015;24(3):245–252.

69. Kinnersley P, Edwards A, Hood K, et al. Interventions before consultations for helping patients address their information needs. Cochrane Database Syst Rev. 2007;2010(3):CD004565.

70. Sansoni JE, Grootemaat P, Duncan C. Question Prompt Lists in health consultations: A review. Patient Educ Couns. 2015;98(12):S0738-3991(15)00258-X.

71. Brown RF, Butow PN, Dunn SM, Tattersall MH. Promoting patient participation and shortening cancer consultations: a randomised trial. Br J Cancer. 2001;85(9):1273–1279.

72. Ramlakhan JU, Dhanani S, Berta WB, Gagliardi AR. Optimizing the design and implementation of question prompt lists to support person-centred care: A scoping review. Health Expect. 2023;26(4):1404–1417.

73. Stacey D, Lewis KB, Smith M, et al. Decision aids for people facing health treatment or screening decisions. Cochrane Database Syst Rev. 2024;1(1):CD001431.

74. Clayman ML, Gulbrandsen P, Morris MA. A patient in the clinic; a person in the world. Why shared decision making needs to center on the person rather than the medical encounter. Patient Educ Couns. 2017;100(3):600–604.

75. Hargraves I, LeBlanc A, Shah ND, Montori VM. Shared decision making: The need for patient-clinician conversation, not just information. Health Aff (Millwood). 2016;35(4):627–629.

76. Elwyn G, Frosch D, Thomson R, et al. Shared decision making: a model for clinical practice. J Gen Intern Med. 2012;27(10):1361–1367.

77. Edelen MO, Rodriguez A, Huang W, Gramling R, Ahluwalia SC. A novel scale to assess palliative care patients’ experience of Feeling Heard and understood. J Pain Symptom Manage. 2022;63(5):689–697.e1.

78. Saunders CH, Durand MA, Scalia P, et al. User-Centered Design of the consideRATE Questions, a Measure of People’s Experiences When They Are Seriously Ill. J Pain Symptom Manage. 2021;61(3):555–565.e5.

79. Gramling R, Stanek S, Ladwig S, et al. Feeling Heard and Understood: A Patient-Reported Quality Measure for the Inpatient Palliative Care Setting. J Pain Symptom Manage. 2016;51(2):150–154.

80. Nano JP, Stevens G, Elwyn G, et al. Using consideRATE to evaluate patient experience in a cancer center: Psychometric and healthcare assessments. Psychooncology. 2025;34(10):e70292.

81. Sun X, Briel M, Walter SD, Guyatt GH. Is a subgroup effect believable? Updating criteria to evaluate the credibility of subgroup analyses. BMJ. 2010;340(mar30 3):c117.

82. Schandelmaier S, Briel M, Varadhan R, et al. Development of the Instrument to assess the Credibility of Effect Modification Analyses (ICEMAN) in randomized controlled trials and meta-analyses. CMAJ. 2020;192(32):E901–E906.

83. Proctor E, Silmere H, Raghavan R, et al. Outcomes for implementation research: conceptual distinctions, measurement challenges, and research agenda. Adm Policy Ment Health. 2011;38(2):65–76.

84. Durlak JA, DuPre EP. Implementation matters: a review of research on the influence of implementation on program outcomes and the factors affecting implementation. Am J Community Psychol. 2008;41(3-4):327–350.

85. Saunders CH, Durand MA, Scalia P, et al. “It helps us say what’s important…” Developing Serious Illness Topics: A clinical visit agenda-setting tool. Patient Educ Couns. 2023;113:107764.

86. Jain N, Bernacki RE. Goals of care conversations in serious illness: A practical guide. Med Clin North Am. 2020;104(3):375–389.

87. Bernacki RE, Block SD, American College of Physicians High Value Care Task Force. Communication about serious illness care goals: a review and synthesis of best practices: A review and synthesis of best practices. JAMA Intern Med. 2014;174(12):1994–2003.

88. Pozzar RA, Tulsky JA, Berry DL, et al. Developing a Collaborative Agenda-Setting Intervention (CASI) to promote patient-centered communication in ovarian cancer care: A design thinking approach. Patient Educ Couns. 2024;120(108099):108099.

89. Beckman HB, Frankel RM. The effect of physician behavior on the collection of data. Ann Intern Med. 1984;101(5):692–696.

90. Dyche L, Swiderski D. The effect of physician solicitation approaches on ability to identify patient concerns. J Gen Intern Med. 2005;20(3):267–270.

91. Burggraf L, Stark S, Schedlbauer A, Kühlein T, Roos M. 10 Ideas, concerns and expectations (ICE) in general practice consultations – report of a mixed methods study. In: Oral Presentations. BMJ Publishing Group Ltd; 2019. doi:10.1136/bmjebm-2019-pod.24

92. Hamilton W BN. Patient agendas in primary care. BMJ. 2006;332(7552):1225–1226.

93. Jones VF, Sisson B, Franco S. The effect of a structured encounter form on patient and resident education. J At Mol Phys.

94. Lang F, McCord RS. Agenda setting in the patient-physician relationship. JAMA. 1999;282(10):942–943.

95. Matthias MS, Adams J, Burgess DJ, et al. Communication and Activation in Pain to Enhance Relationships and Treat Pain with Equity (COOPERATE): Rationale, study design, methods, and sample characteristics. Contemp Clin Trials. 2022;118:106790.

96. Morrissey EC, Byrne M, Casey B, et al. Improving outcomes among young adults with type 1 diabetes: the D1 Now pilot cluster randomised controlled trial. Pilot Feasibility Stud. 2022;8(1):56.

97. Rosa WE, Cannity K, Moreno A, et al. Geriatrics communication skills training program for oncology healthcare providers to improve the management of care for older adults with cancer. PEC Innov. 2022;1(100066):100066.

98. Silber TJ, Rosenthal JL. Usefulness of a review of systems questionnaire in the assessment of the hospitalized adolescent. J Adolesc Health Care. 1986;7(1):49–52.

99. Stott NC, Rollnick S, Rees MR, Pill RM. Innovation in clinical method: diabetes care and negotiating skills. Fam Pract. 1995;12(4):413–418.

100. Stott NC, Rees M, Rollnick S, Pill RM, Hackett P. Professional responses to innovation in clinical method: diabetes care and negotiating skills. Patient Educ Couns. 1996;29(1):67–73.

101. Vegni E, Martinoli M, Moja EA. Improving patient-centred medicine: a preliminary experience for teaching communication skills to Italian general practitioners. Educ Health (Abingdon). 2002;15(1):51–57.

102. Wissow L, Gadomski A, Roter D, Larson S, Lewis B, Brown J. Aspects of mental health communication skills training that predict parent and child outcomes in pediatric primary care. Patient Educ Couns. 2011;82(2):226–232.

103. Wolff JL, Scerpella D, Cockey K, et al. SHARING Choices: A pilot study to engage family in advance care planning of older adults with and without cognitive impairment in the primary care context. Am J Hosp Palliat Care. 2021;38(11):1314–1321.

